# Novel Genetic Variants Associated with Gout Identified Through a Genome-Wide Study in the UK Biobank (N = 150,542)

**DOI:** 10.1101/2025.02.07.25321834

**Authors:** Yiwen Tao, Tengda Cai, Qi Pan, Luning Yang, Sen Lin, Mainul Haque, Tania Dottorini, Abhishek Abhishek, Weihua Meng

**Affiliations:** Nottingham Ningbo China Beacons of Excellence Research and Innovation Institute, University of Nottingham Ningbo China, Ningbo, China, 315100; School of Mathematical Sciences, University of Nottingham Ningbo China, Ningbo, China, 315100; School of Veterinary Medicine and Science, University of Nottingham, Nottingham, UK, LE12 5RD; Division of Rheumatology, Orthopaedics and Dermatology, School of Medicine, University of Nottingham, Nottingham, UK, NG5 1PB; Division of Population Health and Genomics, Ninewells Hospital and Medical School, University of Dundee, Dundee, UK, DD2 4BF; Center for Public Health, Faculty of Medicine, Health and Life Sciences, School of Medicine, Dentistry and Biomedical Sciences, Queen’s University Belfast, Belfast, UK, BT12 6BA

**Keywords:** Gout, hyperuricemia, UK Biobank, genome-wide association, tissue-specific gene expression, transcriptome-wide association studies

## Abstract

Gout is a prevalent and painful form of inflammatory arthritis associated with hyperuricemia, which leads to monosodium urate crystal deposition in joints and surrounding tissues, triggering acute inflammatory responses. This disease is also closely linked to serious comorbidities, including cardiovascular diseases, chronic kidney diseases, diabetes, and increased mortality risk, significantly impacting global health. In this study, we conducted a comprehensive genome-wide association study (GWAS) based on the UK Biobank pain questionnaire 2019, comprising 10,474 gout cases and 140,068 controls, identifying 13 loci associated with gout, including four novel loci. These findings were further explored in the FinnGen cohort, with 12 loci being replicated significantly. Sex-stratified analyses revealed notable differences, with 16 loci identified in males and two loci identified in females, reflecting both shared and sex-specific genetic influences on gout susceptibility. In addition, genetic correlation analyses demonstrated strong associations between gout and traits related to urate levels, specific medication use, and metabolic functions. Transcriptome-wide association studies highlighted several genes, such as *SLC16A9* and *ASAH2B*, which showed significant expression patterns across various tissues, implicating metabolic and immune pathways in gout. Phenome-wide association studies of significant SNPs revealed links to metabolic, immunological, and skeletal traits, underscoring the multi-faceted nature of gout. These results contribute valuable insights into the genetic architecture and biological mechanisms underlying gout, suggesting potential avenues for tailored interventions.

## Introduction

Gout is a common and painful inflammatory arthritis that affects millions of people worldwide. It is commonly related to hyperuricemia, leading to the deposition of monosodium urate crystals in joints and tissues, triggering acute inflammation ^1,2^. Most instances of gout are characterized by the rapid development of severe acute monarticular arthritis in a peripheral joint of the leg ^3^. Over recent decades, the incidence of gout has steadily increased, largely due to lifestyle factors such as diet, obesity, and metabolic conditions ^4^. Gout is also significantly linked to cardiovascular diseases, kidney diseases, diabetes, and early mortality, further compounding its impact on public health ^5,6^.

Epidemiological evidence from multiple countries indicates that gout is becoming increasingly prevalent, with men being more susceptible than women. Globally, the prevalence of gout is estimated at 1-4%, with 3-6% of men and 1-2% of women affected in Western countries ^7^. In the UK, the standardized prevalence of gout grew by 63.9% between 1997 (1.52%) and 2012 (2.49%) ^8^. Similarly, an analysis of a health-care database from Canada, showed an increase in gout prevalence from 3% in 2003 to 3.8% in 2012 ^9^. A study conducted in Mainland China reported an increase in gout prevalence from 0.9% in 2000 to 1.4% in 2014 ^10^.

Genetic factors play a crucial role in the development of gout, with several studies highlighting its strong hereditary component. Twin studies have demonstrated a strong genetic component in gout, with heritability estimates reaching 60% for uric acid kidney clearance, 87% for uric acid-to-creatinine ratios, and 28% to 31% for gout itself ^9,11^. Additionally, a population-based study confirmed that gout runs in families, with higher risk for individuals with a family history, influenced by both genetic and environmental factors that vary by sex ^12^. A study from the UK Biobank further estimated that the heritability of elevated serum urate and gout, driven by common genetic variants, is similar among European individuals ^11^. Genome-wide association studies (GWAS) have greatly advanced our understanding of the genetic architecture of gout. Several genetic loci have been identified to be associated with gout risk, including *SLC2A9*, *ABCG2*, *SLC17A3, ABCG2*, *SLC2A9*, *SLC22A11*, *GCKR*, *MEPE*, *PPM1K-DT*, *LOC105377323* and *ADH1B* ^13,14^. During the preparation of this paper, a new genome-wide association analysis revealed 377 loci and 410 genetically independent signals associated with gout, including 149 previously unreported loci ^15^.

The purpose of this study was to identify novel genetic variants associated with gout using data from the UK Biobank, specifically leveraging the 2019 pain questionnaire to refine the gout phenotype definition. This updated definition aimed for greater precision in classifying cases and controls for pain related phenotypes, resulting in more accurate definitions. Sex-specific GWAS were conducted to investigate potential genetic variants with differential effects between males and females. Furthermore, we sought to replicate our findings using the FinnGen Biobank cohort ^16^ and performed post-GWAS analyses, including pathway analysis and gene prioritization, to gain deeper insights into the biological mechanisms driving gout.

## Materials and Methods

### Data information

This study used UK Biobank data, a large-scale health resource comprising over 500,000 participants aged 40-69 from across the UK. For this analysis, we used the pain questionnaire conducted in 2019 (https://biobank.ctsu.ox.ac.uk/crystal/ukb/docs/pain_questionnaire.pdf), with responses from 166,733 individuals. Participants provided informed consent to complete health-related questionnaires and donate biological samples. To minimize population stratification, the analysis in this study was conducted exclusively on white British participants (Field ID: 21000). Ethical approval for the UK Biobank was granted by the National Research Ethics Service (reference 11/NW/0382).

### Case and control information

Based on responses to the second UK Biobank questionnaire question, “Have you ever been told by a doctor that you have had any of the following conditions?” from the UK Biobank 2019 pain questionnaire. Participants who answered “yes” for gout were categorized as cases, while controls were those who answered “no”. This allowed us to capture self-reported gout cases and define controls with no history of gout. To ensure data quality, we excluded participants who chose “Do not know” or “Prefer not to answer,” as well as those with missing responses.

### GWAS design and statistical analysis

One primary GWAS was conducted to identify genetic variants associated with gout and two sex-stratified GWAS was performed to explore sex impact on gouts. In the replication phase, we used publicly accessible M13 gout statistics from the FinnGen dataset. This dataset includes 12,723 gout cases and 507,487 controls. For readers’ interest, we have also included six additional GWAS on urate values, details of which were described in the Supplementary Files. Figure 1 outlines the workflow of the GWAS in this study, including the three GWAS presented in the main text and six additional analyses provided in the Supplementary Files.

**Fig. 1.**
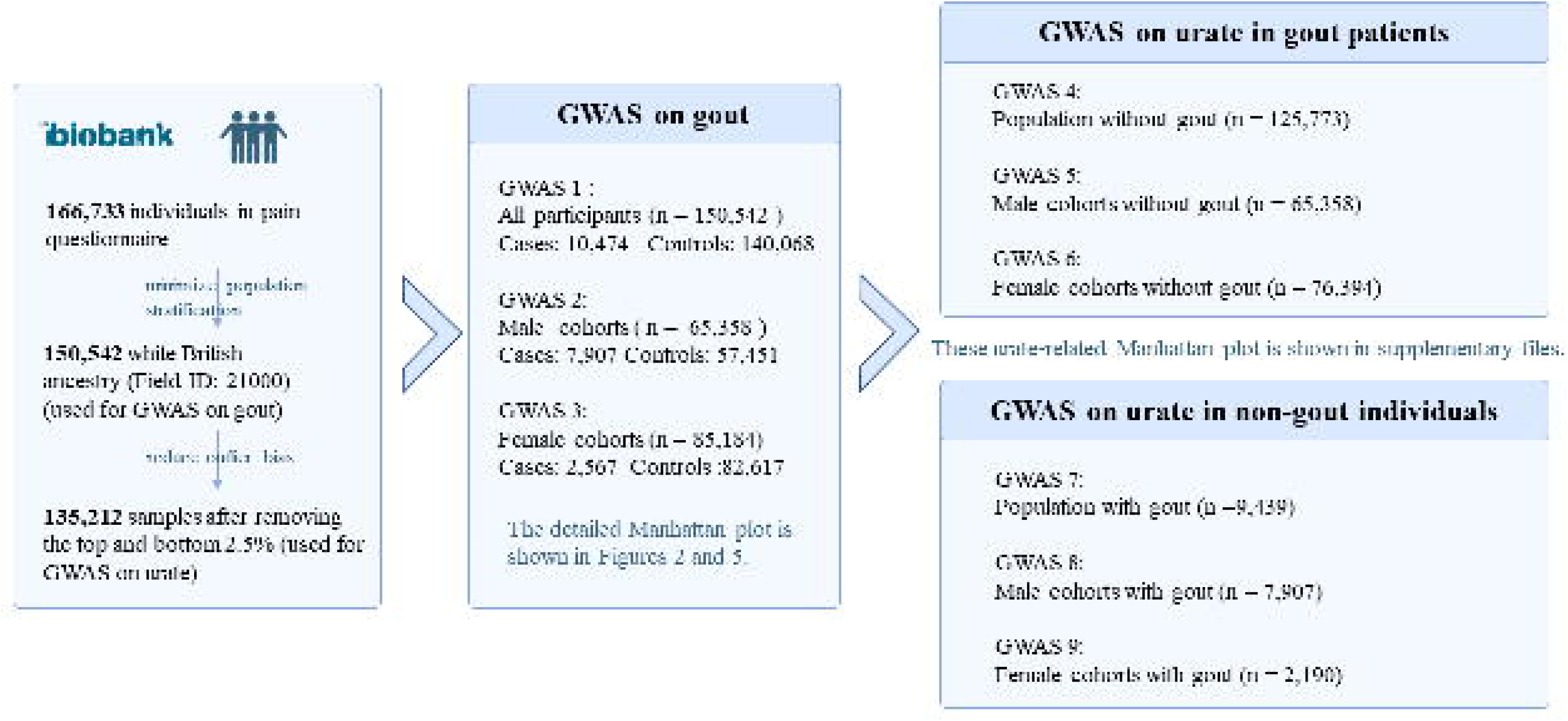
Overview of GWAS analyses on gout and urate levels. This figure summarizes the GWAS design, including 166,733 UK Biobank participants, with 150,542 White British individuals selected to minimize population stratification. GWAS on gout included all participants (n = 150,542), male cohorts (n = 65,358), and female cohorts (n = 85,184). To reduce outlier bias, 135,212 samples were analyzed for urate GWAS after excluding the top and bottom 2.5% of urate levels, stratified into populations without gout (n = 125,773) and with gout (n = 9,439). Detailed Manhattan plots are shown in Figures 2 and 5, with supplementary plots in Supplementary Files.

We used the fastGWA tool in GCTA (v1.94.1) to perform a genome-wide association analysis, applying a generalized mixed linear model ^17^. Quality control measures included removing SNPs with minor allele frequencies below 0.5%, low INFO scores (< 0.3), and those violating Hardy-Weinberg equilibrium (*p* < 1 × 10^−6^). Variants on mitochondrial DNA and sex chromosomes were excluded. The analysis was adjusted for age, sex, and the first ten principal components to account for population structure. A genome-wide significance threshold of *p* < 5 × 10^−8^ was used, and narrow-sense heritability was estimated using GCTA.

### Functional annotation and genetic correlation analysis

We used Functional Mapping and Annotation (FUMA) to annotate the GWAS results ^18^. FUMA integrates several analytical tools, including gene-based association analysis, gene-set analysis, and tissue expression analysis, using default parameters. Gene-based and gene-set analyses were conducted with MAGMA (v1.06) integrated in FUMA. In gene-based association analysis, genes were mapped to 19,023 protein-coding regions, with significance defined at *p* = 0.05/19,203 = 2.60 × 10^−6^. In gene-set analysis, FUMA examines gene collections with shared biological functions, using a significance threshold of *p* = 0.05/15,485 = 3.23 × 10^−6^. Tissue expression analysis was carried out using GTEx v8 datasets, which includes 54 tissue types, to identify genes with tissue-specific expression patterns. We also performed genetic correlation analysis using linkage disequilibrium score regression (LDSC) to estimate the genetic correlations between gout and other traits in the Complex-Traits Genetics Virtual Lab ^19^.

### Integration of eQTL, chromatin architecture, and positional mapping

To explore the regulatory mechanisms linked to the variants identified in the GWAS, we integrated expression quantitative trait loci (eQTL) analysis, chromatin interaction data, and positional mapping. Cis-eQTLs, which impact gene expression by interacting with nearby variants within a 1 Mb range, were the primary focus. We applied positional mapping with a 10 kb distance threshold to pinpoint regulatory elements associated with identified SNPs.

### Transcriptome- and Phenome-Wide Association Studies (TWAS and PheWAS)

Transcriptome-wide association studies (TWAS) were performed to assess the impact of genetic variants on gene expression in gout-relevant tissues, utilizing GTEx v7 data ^20^. Phenome-wide association studies (PheWAS) were conducted using GWAS summary statistics from 4,756 GWAS results available on the ATLAS platform ^21^, which aimed to uncover associations between gout-linked variants and other traits, with a focus on metabolic and inflammatory phenotypes. Multiple comparisons were corrected using the Bonferroni method.

### Mendelian Randomization Analysis

We employed a two-sample Mendelian Randomization (MR) approach to investigate the potential causal links between other traits and gout. Based on the results from our genetic correlation and PheWAS analyses, we examined whether specific genetic variants had a direct effect on gout. Data for the MR analysis were sourced from the UK Biobank and the IEU Open GWAS platform. The Inverse Variance Weighted (IVW) method was used for the primary analysis, with additional sensitivity analyses conducted using MR Egger, Weighted Median and Simple mode approaches. Heterogeneity was assessed using Cochran’s Q test, and horizontal pleiotropy was evaluated with the MR Egger intercept test. We also performed bidirectional MR to examine the possibility of reverse causation.

### Protein-protein interaction analysis and evaluation of druggability

To investigate the therapeutic potential of significant loci identified in the primary GWAS, we conducted a protein-protein interaction (PPI) analysis to explore interactions between these loci. The PPI analysis was performed using the STRING database (https://string-db.org). We further assessed the druggability of the candidate genes by referencing the Druggable Genome database (https://dgidb.org/) and the ChEMBL database (https://www.ebi.ac.uk/chembl). Druggable genes were categorized into three tiers: tier 1 included targets of approved drugs and clinical-phase drug candidates, tier 2 consisted of genes closely related to approved drug targets or drug-like compounds, and tier 3 comprised genes with more distant similarities to approved drug targets. Information about associated compounds, drug approval status, and therapeutic indications was retrieved from the Finan’s study on 4479 druggable genes^22^ and ChEMBL database.

## Results

### Sample Description

The study utilized data from the UK Biobank’s 2019 pain questionnaire, which provided updated information on gout. A total of 166,733 participants completed this second questionnaire, leading to the identification of a more precisely defined gout phenotype. The data were refined to include only White-British genetic ancestry who met the QC criteria. For the primary GWAS analysis, we included 150,542 individuals and 7,402,791 SNPs, with 10,474 classified as cases (individuals reporting gout) and 140,068 as controls (those reporting no gout). The cohort consisted of 65,358 males (of which 7,907 were cases and 57,451 were controls) and 85,184 females (with 2,567 cases and 82,617 controls). Table 1 provides an overview of the key demographic and clinical characteristics of both the case and control groups. Statistically significant differences were observed between the groups in variables including age, sex and urate with *p* values less than 0.001.

**Table 1.** Clinical characteristics of gout cases and controls in the UK Biobank.

| GWAS Analysis | Covariates | Cases | Controls | *p*-value |
| --- | --- | --- | --- | --- |
| Primary GWAS | Sex (female: male) | 2,567: 7,907 | 82,617: 57,451 | <0.001 |
|  | Age (years) | 58.2 (7.08) | 55.8 (7.64) | <0.001 |
|  | Urate (umol/L) | 394.8 (95.15) | 296.4 (72.23) | <0.001 |
| Female-specific GWAS | Age (years) | 58.2 (6.97) | 55.4 (7.56) | <0.001 |
|  | Urate (umol/L) | 320.1 (94.65) | 263.6 (60.281) | <0.001 |
| Male-specific GWAS | Age (years) | 58.2 (7.12) | 56.3 (7.72) | <0.001 |
|  | Urate (umol/L) | 418.8 (81.94) | 343.3 (61.17) | <0.001 |

Total number of females: 85,184; Total number of males: 65,358.
A $\chi^2$ test was used to test the difference of gender frequency between cases and controls and an independent t test was used for other covariates.
Continuous covariates were presented as mean (standard deviation)
BMI represents body mass index.

### GWAS Results

In the primary GWAS, we identified 13 loci that reached genome-wide significance (*p* < 5 × 10^−8^), as summarized in Table 2 and showed in Figure 2a. Of these, nine loci had been previously reported, while four represented novel discoveries. The most significant association was observed at rs2199936 in the *ABCG2* gene on chromosome 4, with a *p* value of 1.75 × 10^−97^. The second strongest association was at rs58656183 in *SLC2A9*, also on chromosome 4, with a *p* value of 5.52 × 10^−90^ (regional plot is provided in Figure 3). Additionally, we identified four novel loci, including rs149865899 in the *CD160* gene (*p* = 3.85 × 10^−11^), rs28607641 in *UBE2Q2* (*p* = 5.88 × 10^−11^), rs3041216 in *DAP3* (*p* = 1.18 × 10^−10^) and rs644740 in *OVOL1* (*p* = 6.22 × 10^−9^) (regional plots are provided in Figure 4). Figure 2b displays SNP density per 0.1 Mb window across chromosomes, with the highest density on chromosomes 1 and 2. The Q–Q plot for the primary GWAS is shown in Figure 2c, with a genomic control lambda value of 1.118, indicating a close fit to expected values with slight deviations at extreme p values. The comprehensive list of all significant SNPs is available in Supplementary Table 1, and detailed regional plots for all significant loci are provided in Supplementary Figure 1. The SNP-based heritability for gout was estimated at 0.019, with a standard error of 0.3953.

**Fig. 2.**
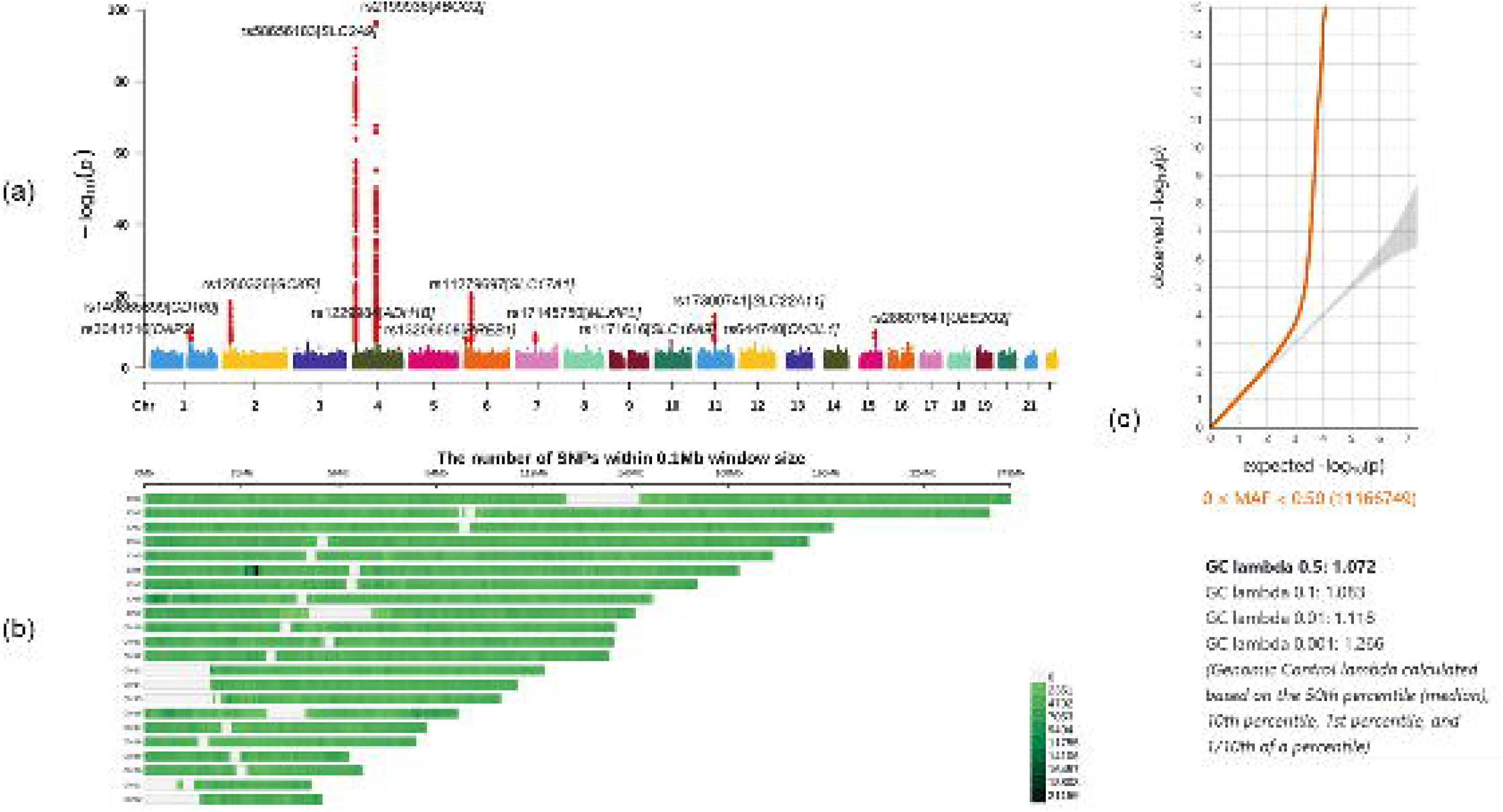
Primary GWAS analysis on gout. **a)** The Manhattan plot of the primary GWAS analysis on gout (N = 150,542). Each dot represents an SNP, plotted by chromosomal position along the x-axis and significance (– log10 *p* value) on the y-axis. The dashed red line indicates the cut-off *p* value of 5□×□10^−8^. **b)** The number of SNPs within 0.1Mb window size on each chromosome, indicating the SNP density and highlighting regions with higher genetic variation. Chromosomes 1 and 2 exhibit the highest SNP density. **c)** The Q-Q plot of the primary GWAS, illustrating the observed versus expected p-value distribution, with GC lambda values calculated across the 50th, 10th, 1st, and 0.1 percentiles (1.072, 1.083, 1.118, and 1.266, respectively), suggesting a close fit to expectations with minor deviations at extreme values.

**Fig. 3.**
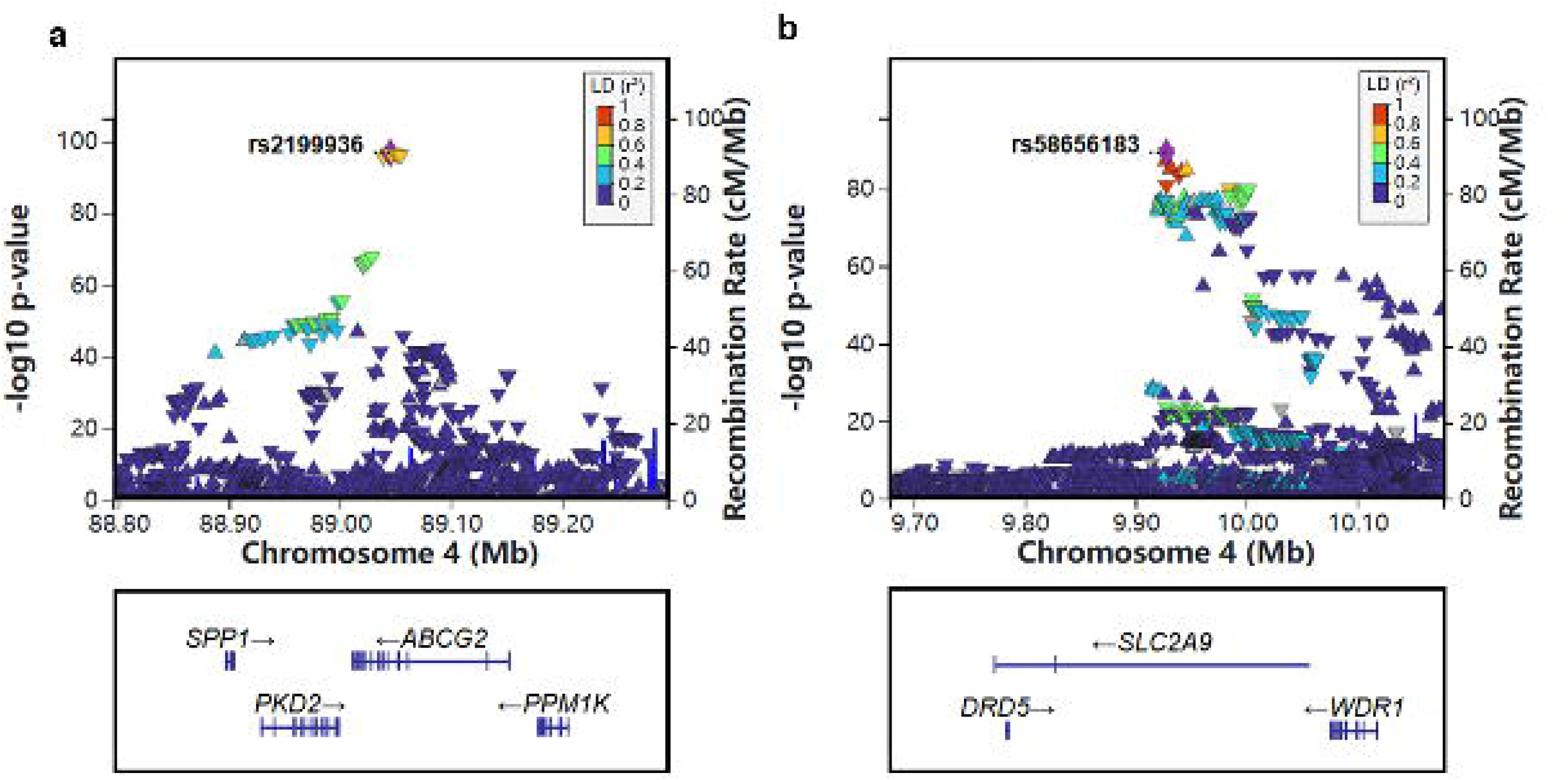
Regional plots of *ABCG2* and *SLC2A9* loci. The regional plot of loci in the *ABCG2* (a) and *SLC2A9* (b) region based on the primary GWAS association analysis. Each plot shows local SNP associations, with the lead SNP highlighted in purple. The x-axis represents the genomic position in megabases (Mb), while the y-axis displays the association strength of each SNP as –log10 *p* value.

**Fig. 4.**
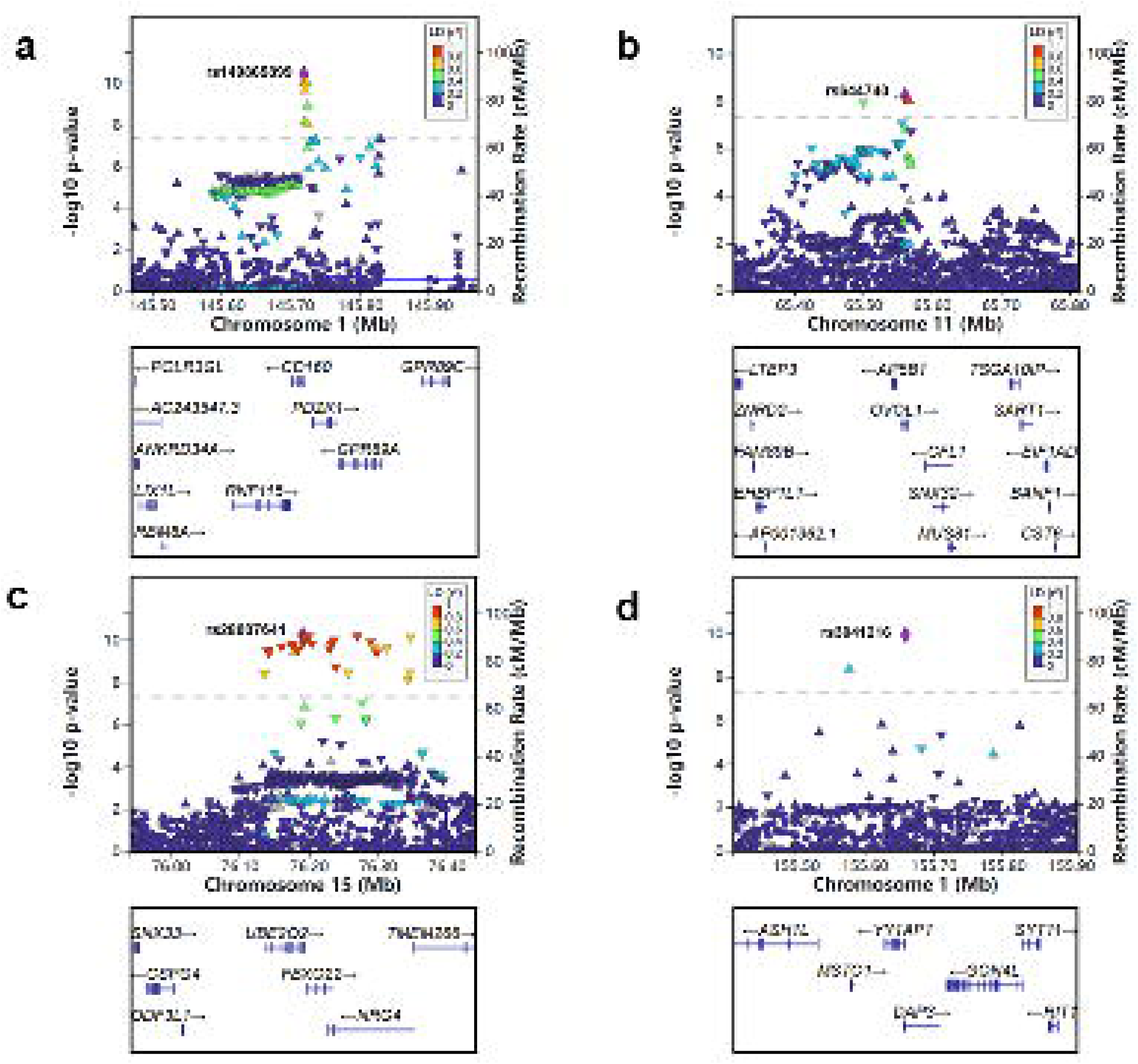
Regional plots of additional loci. The regional plots of loci in the *CD160* (a)*, OVOL1* (b)*, UBE2Q2* (c) and *DAP3* (d) regions based on the primary GWAS association analysis. Each plot shows local SNP associations, with the lead SNP highlighted in purple. The x-axis represents the genomic position in megabases (Mb), while the y-axis displays the association strength of each SNP as –log10 *p* value.

**Table 2.**
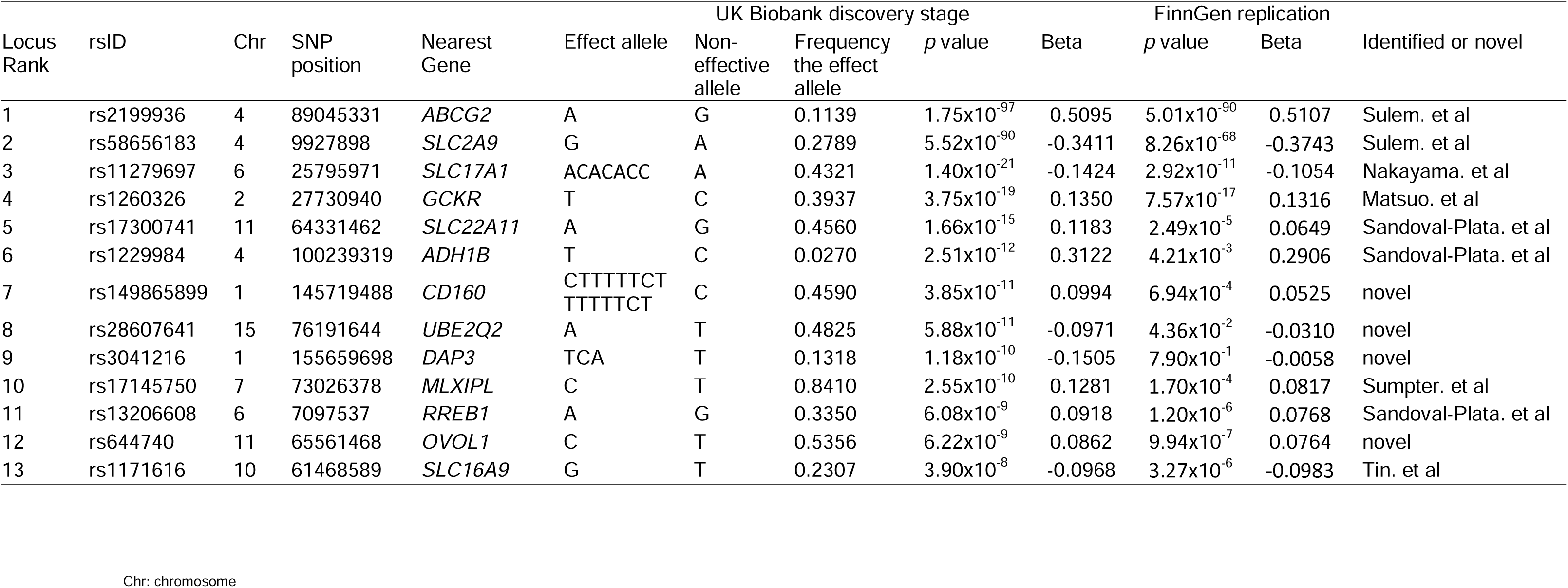
The top SNPs within 13 loci identified by the GWAS on gout.

**Table 3.** The top SNPs within two loci for the female-specific GWAS and 16 loci for the male-specific GWAS on gout.

| Secondary GWAS | Locus Rank | rsID | Chr | SNP position | Nearest Gene | Effect allele | Non-effective allele | Frequency the effect allele | p value | Beta |
| --- | --- | --- | --- | --- | --- | --- | --- | --- | --- | --- |
| Female-specific GWAS | 1 | rs3775948 | 4 | 9995182 | *SLC2A9* | G | C | 0.2452 | $1.1 \times 10^{-12}$ | -0.2416 |
| | 2 | rs2199936 | 4 | 89045331 | *ABCG2* | A | G | 0.1132 | $6.14 \times 10^{-9}$ | 0.2684 |
| Male-specific GWAS | 1 | rs74904971 | 4 | 89050026 | *ABCG2* | C | A | 0.8852 | $8.52 \times 10^{-98}$ | -0.5930 |
| | 2 | rs58656183 | 4 | 9927898 | *SLC2A9* | G | A | 0.2781 | $1.06 \times 10^{-82}$ | -0.3805 |
| | 3 | rs11279697 | 6 | 25795971 | *SLC17A1* | ACACACC | A | 0.4319 | $2.09 \times 10^{-22}$ | -0.1691 |
| | 4 | rs60821450 | 11 | 64327150 | *SLC22A11* | C | CT | 0.4379 | $8.04 \times 10^{-18}$ | 0.1506 |
| | 5 | rs1260326 | 2 | 27730940 | *GCKR* | T | C | 0.3963 | $1.27 \times 10^{-15}$ | 0.1402 |
| | 6 | rs1229984 | 4 | 100239319 | *ADH1B* | T | C | 0.0287 | $2.86 \times 10^{-14}$ | 0.3924 |
| | 7 | rs149865899 | 1 | 145719488 | *CD160* | CTTTTCT<br>TTTTTCT | C | 0.4608 | $1.80 \times 10^{-13}$ | 0.1289 |
| | 8 | rs6957745 | 7 | 73056750 | *MLXIPL* | T | C | 0.7985 | $4.54 \times 10^{-12}$ | 0.1494 |
| | 9 | rs7964492 | 12 | 57823585 | *R3HDM2* | A | C | 0.7617 | $4.31 \times 10^{-11}$ | 0.1333 |
| | 10 | rs1171616 | 10 | 61468589 | *SLC16A9* | G | T | 0.2302 | $1.66 \times 10^{-9}$ | -0.1236 |
| | 11 | rs199875356 | 15 | 76347375 | *NRG4* | CT | C | 0.4506 | $2.18 \times 10^{-9}$ | -0.1041 |
| | 12 | rs12203597 | 6 | 7085512 | *RREB1* | G | A | 0.3302 | $2.44 \times 10^{-9}$ | 0.1093 |
| | 13 | rs13082026 | 3 | 52962681 | *SFMBT1* | C | T | 0.5656 | $9.33 \times 10^{-9}$ | -0.1034 |
| | 14 | rs644740 | 11 | 65561468 | *OVOL1* | C | T | 0.5359 | $9.37 \times 10^{-9}$ | 0.0992 |
| | 15 | rs3041216 | 1 | 155659698 | *DAP3* | TCA | T | 0.1318 | $1.93 \times 10^{-8}$ | -0.1527 |
| | 16 | rs12933062 | 16 | 69812097 | *WWP2* | T | A | 0.6265 | $3.54 \times 10^{-8}$ | 0.1006 |

Chr: chromosome

To assess the replication of the genetic variants associated with gout identified in our primary GWAS, we examined the *p* values and effect sizes of these variants in the FinnGen M13 gout dataset. This dataset includes 12,723 gout cases and 507,487 controls. Out of the tested loci, 12 demonstrated significant replication after Bonferroni correction (*p* < 0.05/12 = 0.0042), including rs2199936 in *ABCG2* on chromosome 4 (*p* = 5.01 × 10^−90^), rs58656183 in *SLC2A9* on chromosome 4 (*p* = 8.26 × 10^−68^), rs11279697 in *SLC17A1* on chromosome 6, (*p* = 2.92 × 10^−11^) and rs1260326 in *GCKR* on chromosome 2 (*p* = 7.57 × 10^−17^), rs17300741 in *SLC22A11* on chromosome 11 (*p* = 2.49 × 10^−5^), rs1229984 in *ADH1B* on chromosome 4 (*p* = 4.21 × 10^−3^), rs149865899 in *CD160* on chromosome 1 (*p* = 6.94 × 10^−4^), rs28607641 in *UBE2Q2* on chromosome 15 (*p* = 4.36 × 10^−2^), rs17145750 in *MLXIPL* on chromosome 7 (*p* = 1.70 × 10^−4^), rs13206608 in *RREB1* on chromosome 6 (*p* = 1.20 × 10^−6^), rs644740 in *OVOL1* on chromosome 11 (*p* = 9.94 × 10^−7^), rs1171616 in *SLC16A9* on chromosome 10 (*p* = 3.27 × 10^−6^) (Table 2).

In sex-stratified analyses, we performed separate GWAS for males and females to examine sex-specific genetic contribution, showed in Figure 5. Among females, we identified two significant loci, including rs3775948 in *SLC2A9* (*p* = 1.1 × 10^−12^) and rs2199936 in *ABCG2* (*p* = 6.14 × 10^−9^), consistent with the primary analysis results. In males, we identified 16 significant loci, with the strongest association again seen at rs74904971 in *ABCG2* (*p* = 8.52 × 10^−96^), followed by rs58656183 in *SLC2A9* (*p* = 1.06 × 10^−82^). The male-specific loci independent of the primary GWAS were those in *R3HDM2* (rs7964492, *p* = 4.31 × 10^−11^), *NRG4* (rs199875356, *p* = 2.18 × 10^−9^), *SFMBT1* (rs13082026, *p* = 9.33 ×10^−9^) and *WWP2* (rs12933062, *p* = 3.54 × 10^−8^). The six additional GWAS on urate were provided in the Supplementary Files for reader’ interest.

**Fig. 5.**
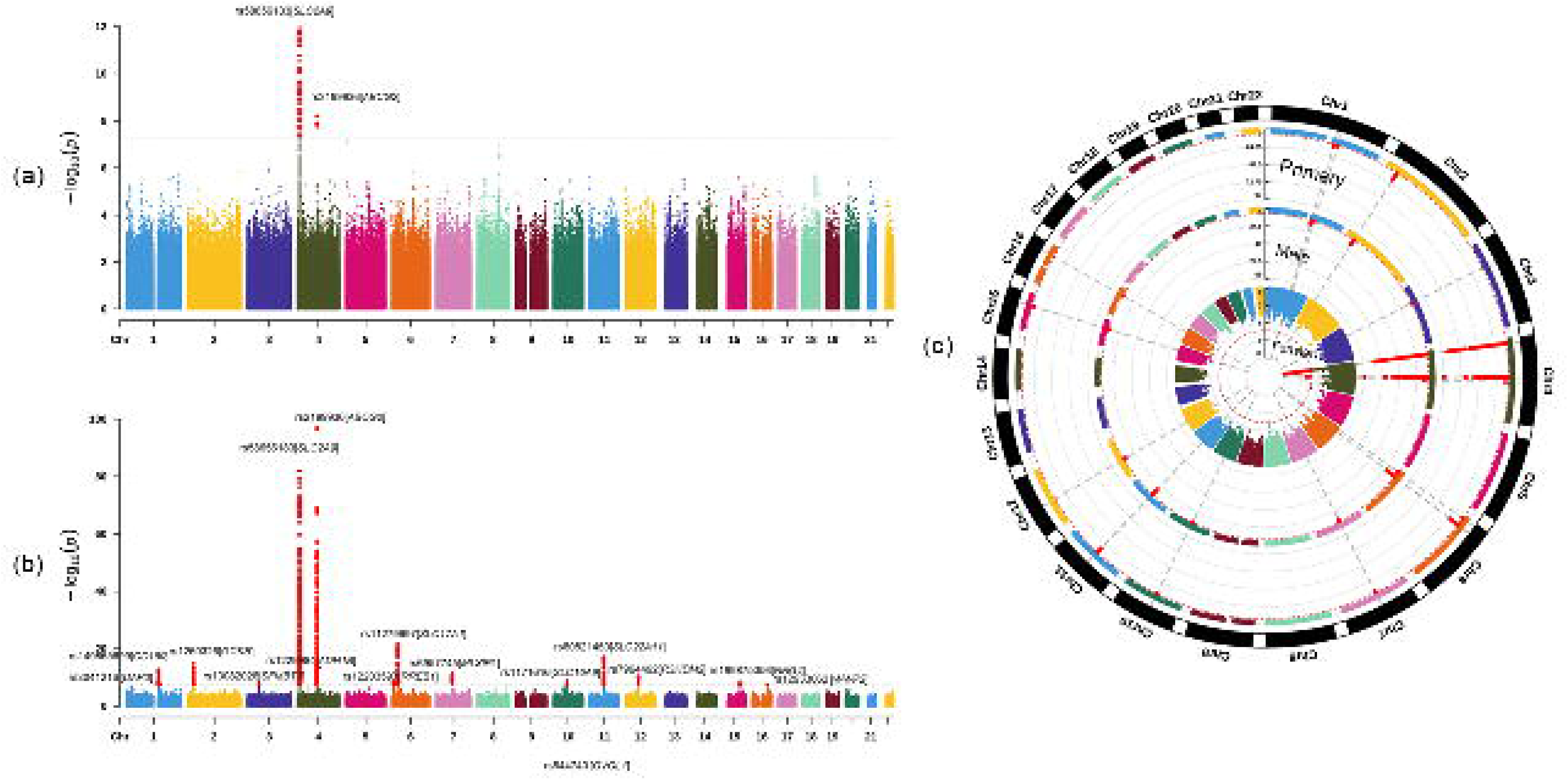
Sex-specific GWAS analyses. **a)** The Manhattan plot of the female-specific GWAS analysis (N = 85,184); **b)** The Manhattan plot of the male-specific GWAS analysis (N= 65,358). Each dot represents an SNP, plotted by chromosomal position along the x-axis and significance (– log10 *p* value) on the y-axis. The dashed red line indicates the cut-off *p* value of 5□×□10^−8^. **c)** A circular Manhattan plot summarizing results from the three GWAS (primary, female-specific, and male-specific), providing a comprehensive comparison of significant loci across cohorts.

### Gene, Gene-Set, and Tissue-Specific Expression Analysis Using FUMA

The gene-based analysis identified *SLC2A9* as the most significant gene linked to gout. In addition, many other genes, such as *ABCG2*, *SLC22A11*, and *SLC17A1*, also showed strong associations with gout, all surpassing the genome-wide significance threshold after the Bonferroni correction test (*p* = 0.05/19,203 = 2.60 × 10^−6^). A detailed Manhattan plot is presented in Supplementary Figure 2.

The gene-set analysis revealed the “GOBP_URATE_METABOLIC_PROCESS” as the most significantly associated set (*p* = 3.28 × 10^−16^), followed by “GOMF_URATE_TRANSMEMBRANE_TRANSPORTER_ACTIVITY” (*p* = 4.17 × 10^−9^) and “GOBP_URATE_TRANSPORT” (*p* = 2.88 × 10 ^−8^). The top ten gene sets identified are listed in Supplementary Table 2.

The tissue-specific expression analysis did not yield any significant results, A summary of tissue expression levels is provided in Supplementary Figure 3 and gene expression heatmaps are shown in Supplementary Figure 4.

### Genetic Correlation Analysis Using LDSC

The analysis revealed four correlations had *r_g_* values greater than 0.7, indicating a strong genetic relationship. We found a strong positive correlation with urate (*r_g_* = 0.89, *p* = 2.25 × 10^−119^). Additionally, gout showed correlations with medications including allopurinol (*r_g_* = 0.92, *p* = 4.04× 10^−82^) and colchicine (*r_g_* = 0.80, *p* = 5.43× 10^−5^). Full details are available in Supplementary Table 3. The genetic correlation between male and female participants for gout was also calculated (*r_g_* = 0.64, *p* = 5.50 × 10^−9^).

### Integration of eQTL, chromatin structure, and positional mapping

The integration of eQTL analysis, chromatin interactions, and positional mapping revealed several novel insights into the genetic architecture of gout. The cis-eQTL analysis identified that the most significant association was *SNX17* expression in muscle tissue (*p* = 1.48 × 10^−88^, FDR = 2.15 × 10^−75^) on chromosome 2. The tissues most strongly associated with *ABCG2* were islets (*p* = 4.60 × 10^−12^, FDR = 3.71× 10^−10^), whole blood (*p* = 2.90 × 10^−10^, FDR = 1.46 × 10^−6^) and thyroid (*p* = 9.98 × 10^−10^, FDR = 3.34 × 10^−7^). Detailed results after Bonferroni correction are provided in Supplementary Table 4.

Chromatin architecture analysis showed that on chromosome 2, *GCKR* was found to interact with genes such as *ABHD1*, *SLC30A3*, and *MRPL33*. Chromatin interactions on chromosome 4 revealed distinct regulatory regions associated with two key genes: *ABCG2* and *SLC2A9*. *ABCG2* was linked to genes like *PPM1K*, *HERC6*, and *DMP1*, while *SLC2A9* was associated with *WDR1*, *DRD5*, and *CLNK*, but no chromatin interactions were detected between *ABCG2* and *SLC2A9*. The mapped genes by chromatin interaction on the chromosomes 2 and 4 are provided in Figure 6 and the others are provided in Supplementary Figure 5.

**Fig. 6.**
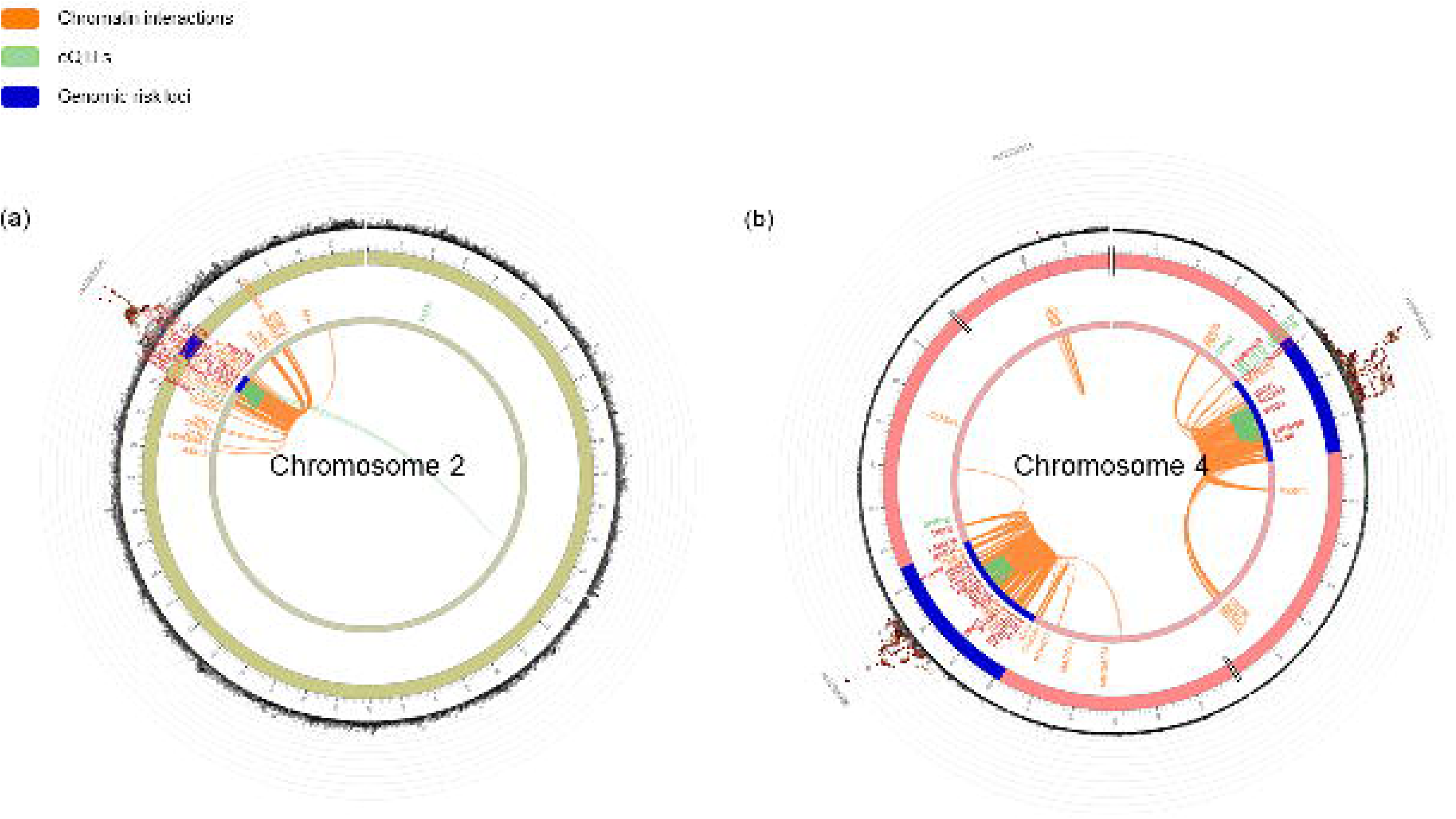
Circos plots of chromatin interactions and eQTL connections. Circos plots illustrating chromatin interactions and eQTL connections relevant to the identified gout-associated loci. The outermost layer shows a Manhattan plot with loci meeting the significance threshold of *p* < 0.05. Each SNP in a genomic risk locus is color-coded according to its maximum LD, measured by r^2^ with an independent significant SNP in the locus: red (r^2^ > 0.8), orange (r^2^ > 0.6), green (r^2^ > 0.4), and blue (r^2^ > 0.2). The middle layer marks genomic risk loci (with lead SNP *p* < 5 × 10^−8^) in blue. The innermost layer highlights eQTLs (green) and/or chromatin interactions (orange). Plots a-b show specific top SNPs on chromosomes 2 and 4 and supplementary figure 5 shows the other SNPs on chromosomes 1, 6, 7, 10, 11, 15 respectively.

### TWAS

Several genes demonstrated statistically significant associations across various tissues. *SLC16A9* showed significant negative associations in multiple tissues, including subcutaneous adipose (Z = −5.45, *p* = 4.97 × 10^−8^) and artery aorta (Z = −5.23, *p* = 1.65 × 10^−7^). Except for *SLC16A9*, no other significant genes from our GWAS showed tissue-specific expression associations. A detailed list of genes and their tissue-specific expression associations after Bonferroni correction can be found in Supplementary Table 5.

### PheWAS

Several SNPs showed strong associations with metabolic, immunological, and skeletal traits. The SNP rs2199936 was notably associated with uric acid levels (*p* = 1.97 × 10^−131^). rs1260326 showed a remarkable association with triglyceride and cholesterol levels (*p* = 2.29 × 10^−239^), while rs17300741 was linked to serum urate levels (*p* = 4.00 × 10^−35^), both of which are directly connected to gout pathology. SNP rs1229984 demonstrated a significant correlation with psychiatric traits, such as the number of drinks per day (*p* = 1.60 × 10^−203^). Additionally, rs644740 was significantly associated with both skeletal and metabolic traits, showing strong links to heel bone mineral density (*p* = 1.30 × 10^−23^) and uric acid levels (*p* = 2.77 × 10^−20^).

Gene-level associations also revealed critical insights. The *ABCG2* gene was highly associated with uric acid levels (*p* = 2.24 × 10^−92^), confirming its role in gout susceptibility. Similarly, *SLC2A9* was significantly linked to serum urate (*p* = 4.77 × 10^−80^) and nucleotide metabolism (*p* = 2.23 × 10^−43^). These findings reinforce the central role of urate transport and purine metabolism in gout development. The *SLC17A1* gene also displayed associations with immunological traits, such as mean corpuscular hemoglobin (*p* = 5.78 × 10^−103^). A detailed list of all traits that passed the Bonferroni correction can be found in Supplementary Table 6, with visual representations of significant phenotypes associated with these SNPs and genes provided in Supplementary Figures 6 and 7.

### MR results

MR analyses used various phenotypes including heel bone mineral density with 426,824 participants (53,184 cases and 373,640 controls), BMI with 461,460 participants, and alcohol consumption with 112,117 participants ^23,24^ from the IEU database (Table 4).

**Table 4.** Comprehensive Details of IEU GWAS Datasets for MR Analyses.

| GWAS dataset | ID | Sample size | Cases | Controls |
| --- | --- | --- | --- | --- |
| Heel bone mineral density | ebi-a-GCST006979 | 426,824 | 53,184 | 373,640 |
| Body mass index | ukb-b-19953 | 461,460 | NA | NA |
| Alcohol consumption | ieu-a-1283 | 112,117 | NA | NA |

In the forward causality analysis, gout was examined as the exposure to evaluate its potential impact on heel bone mineral density, BMI, and alcohol consumption. The Inverse Variance Weighted (IVW) method indicated significant associations between gout and BMI, with an odds ratio (OR) of 0.98 (95% CI: 0.95 to 1.00, *p* = 4.45 × 10^−2^). There is no significant positive causal relationship between gout and heel bone mineral density or alcohol consumption.

Conversely, reverse causality analyses explored whether genetic predictions of heel bone mineral density, BMI, and alcohol consumption had a causal effect on the risk of gout. The IVW method provided compelling evidence for a significant reverse causal relationship between BMI and gout (OR = 1.52, 95% CI: 1.32 to 1.74, *p* = 3.28 ×10^−9^). Additionally, the Weighted Median method further supported this association (*p* = 1.44 × 10^−8^). Other MR methods, including the MR Egger and Simple Mode, demonstrated similar directional trends, though they did not always reach statistical significance, indicating some robustness in these findings across methods. For heel bone mineral density and alcohol consumption, none of the MR methods indicated a significant causal relationship with gout. Detailed results and figures are presented in Supplementary Figure 8 and Supplementary Table 7.

### PPI and association of potential drug targets

13 significant loci identified in the primary GWAS were used to explore their interactions. The resulting network, shown in Supplementary Figure 9, revealed key connections between the proteins encoded by the identified loci. We further analyzed the associations between these significant genes and approved drugs, finding that *ABCG2*, *SLC22A11*, *ADH1B*, and *GCKR* are all classified as Tier-1 druggable genes. These genes interact with various approved medications, including antineoplastic agents, antihypertensive agents, uricosuric agents, and lipid-lowering agents, as detailed in Supplementary Table 8.

## Discussion

In this study, we identified 13 significant genetic loci associated with gout through the primary GWAS using the UK Biobank pain questionnaire. Among these, four loci were novel, while others were previously known. Sex-stratified GWAS revealed 16 loci specific to males and two loci unique to females.

The most significant association was identified at the *ABCG2* locus on chromosome 4 (rs2199936, *p* = 1.75 × 10^−97^). This gene has been extensively studied in relation to uric acid metabolism, and its role in gout has been well established ^25^. The strong signal at this locus reinforces the importance of *ABCG2* in the pathophysiology of gout again, as it encodes a transporter protein that plays a key role in the uric acid efflux transport ^26^. Dysfunctional variants of *ABCG2* can lead to reduced uric acid excretion, thereby contributing to hyperuricemia, a major risk factor for gout ^27^. Another well-established locus identified in our study was *SLC2A9* on chromosome 4 (rs58656183, p = 5.52 × 10^−90^). *SLC2A9* encodes a glucose transporter that also facilitates urate reabsorption in the kidney ^28^. Variants in this gene have been consistently associated with serum urate levels and gout risk ^5,29^. In recent years, both *ABCG2* and *SLC2A9* have been attempted to use as drug targets to treat hyperuricemia and gout ^30,31^

Among the other genes significantly associated with gout, seven loci have also been previously identified in GWAS, including those in the genes *SLC17A1*, *GCKR*, *SLC22A11*, *ADH1B*, *MLXIPL, RREB1* and *SLC16A9* ^15^. *SLC17A1* encodes a sodium-phosphate cotransporter that plays a role in urate transport ^32^. Research has shown that SNPs in the *SLC17A1* gene decrease uric acid excretion and elevate blood UA levels, contributing to hyperuricemia ^33^. Variants in this gene have been associated with altered uric acid levels, influencing gout susceptibility. *GCKR* regulates glucose and lipid metabolism, helping to maintain metabolic balance and protect the liver from damage associated with excess substrates ^34^. *SLC22A11 functions in the reabsorption of uric acid at the renal level, directly influencing urate homeostasis* ^35^. *ADH1B* is primarily known for encoding alcohol dehydrogenase enzyme, which is involved in alcohol metabolism ^36^. Alcohol consumption is a well-known risk factor for gout, and *ADH1B* plays a critical role in modulating this risk. *MLXIPL* (also known as ChREBP) is associated with elevated plasma triglyceride levels, increased concentrations of liver enzymes, and a higher risk of coronary artery disease ^37^. *RREB1* is a transcription factor that regulates genes involved in cellular growth and differentiation ^38^. *SLC16A9* encodes a monocarboxylate transporter involved in uric acid excretion and has been shown to affect serum uric acid levels ^35^. These genes contribute to gout either by directly influencing blood urate levels or by altering metabolic pathways that indirectly impact gout risk.

Among the novel loci identified, *CD160* (rs149865899, *p* = 3.85 × 10^−11^) on chromosome 1 was particularly intriguing. *CD160* is a transmembrane glycoprotein anchored by glycosylphosphatidylinositol, which is primarily known for its role in immune regulation ^39^. *CD160* serves as a negative regulator of CD4+ T cell activation by interacting with herpesvirus entry mediator, thereby inhibiting T cell proliferation and cytokine production ^40^. Inhibiting unnecessary immune responses while maintaining general immunity can prevent excessive immune reactions and help preserve the balance of the immune system ^41^. Its association with gout suggests a potential link between immune response and the development of gout, which could offer new avenues for research into the inflammatory processes underlying the disease. As gout is characterized by acute inflammatory responses to urate crystal deposition, genetic variants in immune-related genes may play a role in determining an individual’s susceptibility to these inflammatory episodes ^42,43^.

Another novel finding was the association of *UBE2Q2* (rs28607641, *p* = 5.88 × 10^−11^) on chromosome 15. This gene encodes a ubiquitin-conjugating enzyme (E2), which is involved in protein degradation pathways ^44^. The ubiquitination process is essential for proteome regulation, with E2 mediating the transfer of ubiquitin-like proteins from ubiquitin-activating enzyme to target proteins ^45^. Ubiquitination plays a critical role in regulating the NF-κB family of transcription factors, which are central to inflammation and carcinogenesis ^46^. In the context of gout, it is possible that variants in *UBE2Q2* could affect inflammatory signaling or the degradation of proteins involved in urate metabolism. Urate crystals deposited in joints can trigger a strong inflammatory response, and altered protein degradation might exacerbate or modulate this process ^47^. Future research could focus on understanding how *UBE2Q2* variants influence the balance between pro-inflammatory and anti-inflammatory signals in gout flares.

The *DAP3* gene (rs3041216, *p* = 1.18 × 10^−10^) on chromosome 1 encodes a protein that is part of the small subunit of the mitochondrial ribosome, playing a crucial role in intramitochondrial protein synthesis and apoptotic signaling ^48,49^. Previous studies indicate that mitochondria play a crucial role in triggering gout attacks, and further research could clarify the effect of this gene on mitochondria and cause gout ^50^. Moreover, The dominant negative form of *DAP3* inhibits apoptosis of FAS/APO-1 and TNF-α, the latter of which binds to TNF-r1 (p55R), activates NF-κB and AP-1, and promotes inflammatory gene expression ^51^. The identification of *DAP3* as a gout-associated gene suggests that mitochondrial health and apoptotic regulation are important factors in gout susceptibility and may represent potential targets for therapeutic intervention.

The forth novel locus identified in our study was *OVOL1* (rs644740, *p* = 6.22 × 10^−9^) on chromosome 11. *OVOL1* is an essential transcription factors for epithelial lineages in vertebrate embryonic development, playing key roles in maintaining the epithelial state and facilitating terminal differentiation during tissue homeostasis ^52^. *OVOL1* has been previously associated with other complex traits, such as atopic dermatitis and metabolic conditions ^53^.

Our sex-stratified GWAS of gout revealed two distinct genetic loci associated with gout risk in females and 16 loci in males, suggesting potential sex-specific genetic mechanisms contributing to the disease. In the female-specific GWAS, the two loci (*SLC2A9* and *ABCG2*) are shared with the primary GWAS results. They do not represent novel findings in the female-specific analysis but reaffirm the importance of these genes in regulating urate levels and gout risk across both sexes. In the male-specific GWAS, the most significant loci that overlapped with the primary GWAS findings include *ABCG2* on chromosome 4 (rs74904971, *p* = 8.52 ×10^−98^), *SLC2A9* on chromosome 4 (rs58656183, *p* = 8.52 ×10^−98^) and *SLC17A1* on chromosome 6 (rs11279697, *p* = 1.06 ×10^−82^). Several loci were uniquely associated with gout in males. These include *R3HDM2* (rs7964492, *p* = 4.31 ×10^−11^), *NRG4* (rs199875356, *p* = 2.18 ×10^−9^), *SFMBT1* (rs13082026, *p* = 9.33 ×10^−9^) and *WWP2* (rs12933062, *p* = 3.54 ×10^−8^). However, these genes have not been directly associated with gout, and further research is needed to explore their potential roles in gout development. One major factor of the difference in the loci number of sex-stratified GWAS is that men (7,907) have a much larger number of cases than women (2,567). With fewer cases in the female cohort, the study’s ability to detect such associations is reduced, likely leading to an underestimation of gout-related loci in women simply due to limited statistical power. Another important factor is that men (average 418.8 umol/L in this study) have naturally higher serum urate levels, a key risk factor for gout, than women (average 320.1 umol/L in this study) (Table 1). This may be because estrogen promotes the excretion of uric acid in the urine of young women ^54^. Gout also differs between sexes in terms of disease presentation. Men are more likely to develop gout earlier and experience more severe symptoms, which may reflect a higher genetic burden detectable in male-specific GWAS ^55^. In contrast, women tend to develop gout later in life, and other age-related factors like comorbidities or medications add complexity to explain the sex specific genetic influence ^55^. The 64% genetic correlation between men and women also helps explain the differences in their genetic mechanisms.

The TWAS analysis revealed several important tissue-specific gene expression associations that may contribute to the biological mechanisms underlying gout. The significant associations of *SLC16A9* expression in adipose and arterial tissues suggest a potential link to adiposity, a known risk factor for gout. Increased body weight and adiposity are important contributors to gout risk, while weight loss has been shown to have a protective effect against its development ^56^. The TWAS results suggest that the genetic architecture of gout is shaped by a complex interplay of metabolic, neurological, and systemic regulatory mechanisms, with several genes showing tissue-specific influences that could be key to developing a more comprehensive understanding of gout pathogenesis.

The PheWAS analysis of novel gout-associated genetic variants revealed important connections between metabolic, immunological, and skeletal traits. The strong association of SNP rs2199936 and rs17300741 with uric acid levels confirms their pivotal role in urate metabolism, which is a key factor in gout development. SNPs such as rs149865899 and rs3041216 were linked to variations in white blood cell counts, suggesting that immune system function may interact with metabolic pathways to influence gout risk. These findings raise the possibility that inflammation and immune response could play a more significant role in gout than previously understood. The identification of rs1229984 was associated with alcohol consumption traits, providing insights into how genetic predispositions to alcohol metabolism influence the likelihood of developing gout. urate metabolism. This dual influence could offer new perspectives on how skeletal traits interact with the metabolic processes leading to gout, particularly in the context of bone health. At the gene level, the strong associations between *ABCG2* and *SLC2A9* with uric acid and serum urate levels reinforce the critical role of these genes in urate transport and metabolism. *SLC2A9*’s additional associations with purine metabolism further highlight that neurological dysfunctions and gout may be linked to specific biochemical abnormalities in purine metabolism ^57^. This PheWAS provides valuable insights into the broad phenotypic spectrum influenced by gout-related genetic variants.

Our MR findings suggest a potential causal association between gout and BMI. This observation aligns with existing literature that has associated higher BMI with a greater risk of developing gout ^58,58^. However, no significant causal relationships were observed between gout and heel bone mineral density or alcohol consumption. This result implies that while gout may impact metabolic and anthropometric factors like BMI, it may not directly affect bone health or alcohol consumption patterns. While observational studies have often linked alcohol intake with gout, our MR analysis suggests that this association may be confounded by other factors or may not represent a direct causal relationship. In summary, these findings highlight the utility of MR in disentangling potential causal relationships between gout and associated traits. Future studies could further investigate the biological mechanisms linking gout with BMI and explore additional phenotypes to fully understand the systemic impact of gout.

Among the identified loci, *ABCG2*, *SLC22A11*, *ADH1B*, and *GCKR* were classified as Tier-1 druggable genes. *GCKR* was found to interact with allopurinol and febuxostat, both approved for the treatment of gout, highlighting its direct relevance to gout pathogenesis and therapy. While no other identified genes were currently linked to approved gout medications, the associated drugs targeting genes such as *ABCG2*, *SLC22A11*, and *ADH1B*—involved in metabolic and inflammatory pathways—may hold potential for repurposing in gout treatment. These findings underscore the need for further investigation to explore the therapeutic applications of these genes in gout and related conditions.

Although the GWAS on gout in the UK Biobank has identified novel loci associated with the disease, several limitations must be acknowledged. One of the primary limitations relates to the disparity in the number of male and female participants. Our study included 7,907 male cases compared to 2,567 female cases. The smaller sample size for females reduces the statistical power to identify associations, meaning that some loci relevant to female gout may have been missed. Future studies with larger female cohorts are necessary to ensure more balanced and comprehensive findings across sexes. Another limitation is the reliance on self-reported data for the classification of gout cases and controls within the UK Biobank. Self-reported diagnoses may introduce biases such as recall inaccuracies or misclassification, particularly for gout, where the condition might be underreported or confused with other joint-related issues. Furthermore, the UK Biobank data lacks detailed clinical information, such as the severity or recurrence of gout episodes, which could provide a more nuanced understanding of the genetic factors influencing different forms of the disease. Additionally, the study’s findings are derived from a predominantly European ancestry cohort, which limits the generalizability of the results to other populations. Genetic variants identified in this study may not be applicable to individuals of non-European descent, thus reducing the external validity of the findings. Future studies should aim to include more diverse populations to better understand the genetic architecture of gout across different ethnicities. While our study adds valuable knowledge to the genetic basis of gout, the limitations discussed such as sample size disparities, reliance on self-reported data, limited population diversity, and lack of functional validation, highlight areas for improvement in future research.

## Conclusion

In summary, our primary GWAS identified 13 genetic loci associated with gout, including four novel ones. The sex-stratified GWAS further revealed 16 loci specific to males and 2 loci specific to females. These findings deepen our understanding of the genetic underpinnings of gout and underscore the importance of considering sex differences in genetic studies, which may help to refine risk assessments and therapeutic strategies for gout in the future.

## Data Availability and Acknowledgement

This study fully complies with the ethical standards and data protection regulations of the UK Biobank. The research was conducted using data from the UK Biobank under Application Number 89386. The summary statistics from the UK Biobank regarding gout will be made available upon publication. Additional data relevant to this study, not included in the article or supplementary materials, can be obtained from the authors upon reasonable request.

## Author information

## Authors’ Contributions

All authors contributed to the development of the study. YT conducted the GWAS analysis using the UK Biobank data and prepared the initial manuscript draft. TC, QP, SL and LY were responsible for data preparation. MH, TD and AA contributed critical feedback on the manuscript. WM coordinated the project, supervised research activities, and offered additional insights and revisions.

## Consent to Publish

All authors have agreed to the publication of this work.

## Ethical Approval

This research received approval from the Ethics Committee of the University of Nottingham Ningbo China.

## References

1 Choi HK, Mount DB, Reginato AM. Pathogenesis of Gout. Ann Intern Med 2005; 143: 499–516.

2 N D, Tr M, Lk S. Gout. Lancet (London, England) 2016; 388. doi:10.1016/S0140-6736(16)00346-9.

3 Emmerson BT. The Management of Gout. New England Journal of Medicine 1996; 334: 445–451.

4 Kuo C-F, Grainge MJ, Zhang W, Doherty M. Global epidemiology of gout: prevalence, incidence and risk factors. Nat Rev Rheumatol 2015; 11: 649–662.

5 Reginato AM, Mount DB, Yang I, Choi HK. The genetics of hyperuricaemia and gout. Nat Rev Rheumatol 2012; 8: 610–621.

6 Feig DI, Kang D-H, Johnson RJ. Uric Acid and Cardiovascular Risk. N Engl J Med 2008; 359: 1811–1821.

7 Ragab G, Elshahaly M, Bardin T. Gout: An old disease in new perspective – A review. Journal of Advanced Research 2017; 8: 495–511.

8 Kuo C-F, Grainge MJ, Mallen C, Zhang W, Doherty M. Rising burden of gout in the UK but continuing suboptimal management: a nationwide population study. 2015. doi:10.1136/annrheumdis-2013-204463.

9 Dehlin M, Jacobsson L, Roddy E. Global epidemiology of gout: prevalence, incidence, treatment patterns and risk factors. Nat Rev Rheumatol 2020; 16: 380–390.

10 Liu R et al. Prevalence of Hyperuricemia and Gout in Mainland China from 2000 to 2014: A Systematic Review and Meta-Analysis. BioMed Research International 2015; 2015: 762820.

11 Cadzow M, Merriman TR, Dalbeth N. Performance of gout definitions for genetic epidemiological studies: analysis of UK Biobank. Arthritis Research & Therapy 2017; 19: 181.

12 Kuo C-F et al. Familial aggregation of gout and relative genetic and environmental contributions: a nationwide population study in Taiwan. 2015. doi:10.1136/annrheumdis-2013-204067.

13 Dehghan A et al. Association of three genetic loci with uric acid concentration and risk of gout: a genome-wide association study. https://www.thelancet.com/journals/lancet/article/PIIS0140-6736(08)61343-4/abstract (accessed 11 Oct2024).

14 Sandoval-Plata G, Morgan K, Abhishek A. Variants in urate transporters, ADH1B, GCKR and MEPE genes associate with transition from asymptomatic hyperuricaemia to gout: results of the first gout versus asymptomatic hyperuricaemia GWAS in Caucasians using data from the UK Biobank. 2021. doi:10.1136/annrheumdis-2020-219796.

15 Major TJ et al. A genome-wide association analysis reveals new pathogenic pathways in gout. Nat Genet 2024. doi:10.1038/s41588-024-01921-5.

16 Kurki MI et al. FinnGen provides genetic insights from a well-phenotyped isolated population. Nature 2023; 613: 508–518.

17 Jiang L, Zheng Z, Fang H, Yang J. A generalized linear mixed model association tool for biobank-scale data. Nat Genet 2021; 53: 1616–1621.

18 Watanabe K, Taskesen E, van Bochoven A, Posthuma D. Functional mapping and annotation of genetic associations with FUMA. Nature Communications 2017; 8: 1826.

19 Bulik-Sullivan BK et al. LD Score regression distinguishes confounding from polygenicity in genome-wide association studies. Nature Genetics 2015; 47: 291–295.

20 Gusev A et al. Integrative approaches for large-scale transcriptome-wide association studies. Nat Genet 2016; 48: 245–252.

21 Watanabe K et al. A global overview of pleiotropy and genetic architecture in complex traits. Nature Genetics 2019; 51: 1339–1348.

22 Finan C et al. The druggable genome and support for target identification and validation in drug development. Science Translational Medicine 2017; 9: eaag1166.

23 Morris JA et al. An atlas of genetic influences on osteoporosis in humans and mice. Nat Genet 2019; 51: 258–266.

24 Clarke T-K et al. Genome-wide association study of alcohol consumption and genetic overlap with other health-related traits in UK Biobank (N=112 117). Mol Psychiatry 2017; 22: 1376–1384.

25 Sulem P et al. Identification of low-frequency variants associated with gout and serum uric acid levels. Nat Genet 2011; 43: 1127–1130.

26 Woodward OM. ABCG2: the molecular mechanisms of urate secretion and gout. American Journal of Physiology-Renal Physiology 2015; 309: F485–F488.

27 Stiburkova B, Pavelcova K, Pavlikova M, Ješina P, Pavelka K. The impact of dysfunctional variants of ABCG2 on hyperuricemia and gout in pediatric-onset patients. Arthritis Res Ther 2019; 21: 77.

28 Caulfield MJ et al. SLC2A9 Is a High-Capacity Urate Transporter in Humans. PLOS Medicine 2008; 5: e197.

29 Vitart V et al. SLC2A9 is a newly identified urate transporter influencing serum urate concentration, urate excretion and gout. Nat Genet 2008; 40: 437–442.

30 Ristic B, Sikder MOF, Bhutia YD, Ganapathy V. Pharmacologic inducers of the uric acid exporter ABCG2 as potential drugs for treatment of gouty arthritis. Asian Journal of Pharmaceutical Sciences 2020; 15: 173–180.

31 Itahana Y, Han R, Barbier S, Lei Z, Rozen S, Itahana K. The uric acid transporter SLC2A9 is a direct target gene of the tumor suppressor p53 contributing to antioxidant defense. Oncogene 2015; 34: 1799–1810.

32 Chung S, Kim G-H. Urate Transporters in the Kidney: What Clinicians Need to Know. Electrolytes & Blood Pressure : E & BP 2021; 19: 1.

33 Vadakedath S, Kandi V. Probable Potential Role of Urate Transporter Genes in the Development of Metabolic Disorders. Cureus 2018; 10: e2382.

34 Zhang Z, Ji G, Li M. Glucokinase regulatory protein: a balancing act between glucose and lipid metabolism in NAFLD. Front Endocrinol 2023; 14. doi:10.3389/fendo.2023.1247611.

35 Wright AF, Rudan I, Hastie ND, Campbell H. A ‘complexity’ of urate transporters. Kidney International 2010; 78: 446–452.

36 Edenberg HJ. The Genetics of Alcohol Metabolism: Role of Alcohol Dehydrogenase and Aldehyde Dehydrogenase Variants. Alcohol Research & Health 2007; 30: 5.

37 Havula E, Hietakangas V. Glucose sensing by ChREBP/MondoA–Mlx transcription factors. Seminars in Cell & Developmental Biology 2012; 23: 640–647.

38 Deng Y-N, Xia Z, Zhang P, Ejaz S, Liang S. Transcription Factor RREB1: from Target Genes towards Biological Functions. International Journal of Biological Sciences 2020; 16: 1463.

39 Piotrowska M, Spodzieja M, Kuncewicz K, Rodziewicz-Motowidło S, Orlikowska M. CD160 protein as a new therapeutic target in a battle against autoimmune, infectious and lifestyle diseases. Analysis of the structure, interactions and functions. European Journal of Medicinal Chemistry 2021; 224: 113694.

40 Cai G, Anumanthan A, Brown JA, Greenfield EA, Zhu B, Freeman GJ. CD160 inhibits activation of human CD4+ T cells through interaction with herpesvirus entry mediator. Nat Immunol 2008; 9: 176–185.

41 del Rio ML, Lucas CL, Buhler L, Rayat G, Rodriguez-Barbosa JI. HVEM/LIGHT/BTLA/CD160 cosignaling pathways as targets for immune regulation. Journal of Leukocyte Biology 2010; 87: 223–235.

42 Dalbeth N, Phipps-Green A, Frampton C, Neogi T, Taylor WJ, Merriman TR. Relationship between serum urate concentration and clinically evident incident gout: an individual participant data analysis. Annals of the Rheumatic Diseases 2018; 77: 1048–1052.

43 Housley WJ et al. Genetic variants associated with autoimmunity drive NFκB signaling and responses to inflammatory stimuli. Science Translational Medicine 2015; 7: 291ra93–291ra93.

44 Chang R et al. Upregulated expression of ubiquitin-conjugating enzyme E2Q1 (UBE2Q1) is associated with enhanced cell proliferation and poor prognosis in human hapatocellular carcinoma. J Mol Hist 2015; 46: 45–56.

45 Grzmil P et al. Embryo implantation failure and other reproductive defects in Ube2q1-deficient female mice. 2013. doi:10.1530/REP-12-0054.

46 Voutsadakis IA. Ubiquitin- and ubiquitin-like proteins-conjugating enzymes (E2s) in breast cancer. Mol Biol Rep 2013; 40: 2019–2034.

47 McCarty DJ. Crystal-Induced Inflammation of the Joints. Annual Review of Medicine 1970; 21: 357–366.

48 Miller JL, Koc H, Koc EC. Identification of phosphorylation sites in mammalian mitochondrial ribosomal protein DAP3. Protein Science 2008; 17: 251–260.

49 Kim H-R et al. Mammalian dap3 is an essential gene required for mitochondrial homeostasis in vivo and contributing to the extrinsic pathway for apoptosis. The FASEB Journal 2007; 21: 188–196.

50 Tseng C-C et al. Next-generation sequencing profiling of mitochondrial genomes in gout. Arthritis Res Ther 2018; 20: 137.

51 Kissil J. DAP genes: Structural and functional study of DAP-3 and implications of DAP-kinase in tumorigenesis.pdf. 1999.

52 Saxena K, Srikrishnan S, Celia-Terrassa T, Jolly MK. OVOL1/2: Drivers of Epithelial Differentiation in Development, Disease, and Reprogramming. Cells Tissues Organs 2020; 211: 183–192.

53 Sun P et al. OVOL1 Regulates Psoriasis-Like Skin Inflammation and Epidermal Hyperplasia. Journal of Investigative Dermatology 2021; 141: 1542–1552.

54 Liu H et al. Low expression of estrogen receptor β in renal tubular epithelial cells may cause hyperuricemia in premenopausal patients with systemic lupus erythematosus. Lupus 2021; 30: 560–567.

55 Dirken-Heukensfeldt KJ, Teunissen T, van de Lisdonk E, Lagro-Janssen A. “Clinical features of women with gout arthritis.” A systematic review. Clin Rheumatol 2010; 29: 575–582.

56 Merriman TR, Dalbeth N. The genetic basis of hyperuricaemia and gout. Joint Bone Spine 2011; 78: 35–40.

57 Curto R, Voit OE, Cascante M. Analysis of abnormalities in purine metabolism leading to gout and to neurological dysfunctions in man. Biochemical Journal 1998; 329: 477–487.

58 Raharjo S, Andiana O. Association of Body Mass Index with The Risk Of Gout Arthritis in Male and Female with Underweight, Normal Weight, Overweight, Obese. Jurnal Ilmiah Mandala Education 2022; 8. doi:10.58258/jime.v8i2.3035.

