## Supplementary figures and images for "Novel Genetic Variants Associated with Gout Identified Through a Genome-Wide Study in the UK Biobank (N = 150,542)"

### Supplementary Figure 1

#### Slide 1
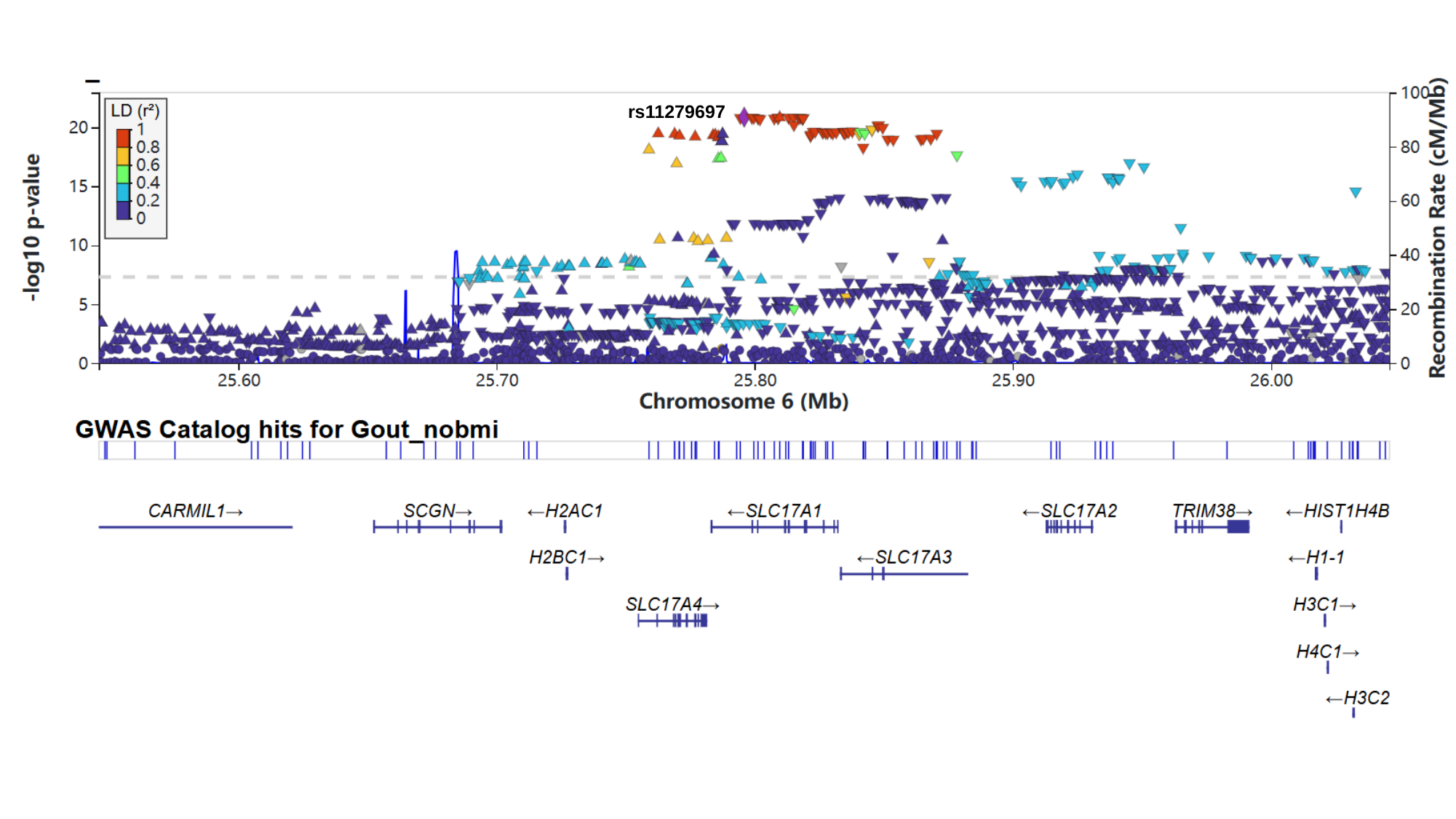

rs11279697

#### Slide 2
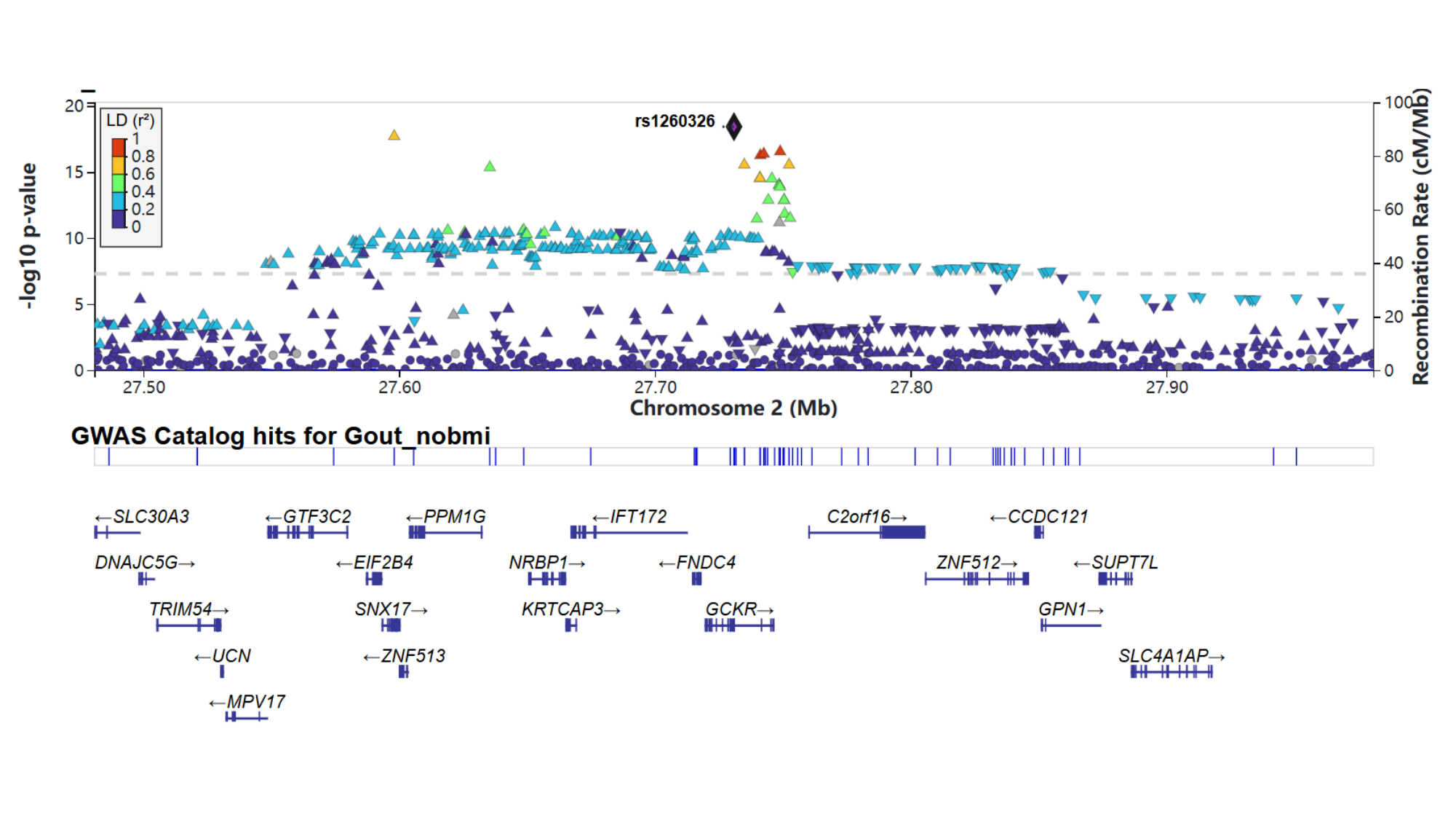

#### Slide 3
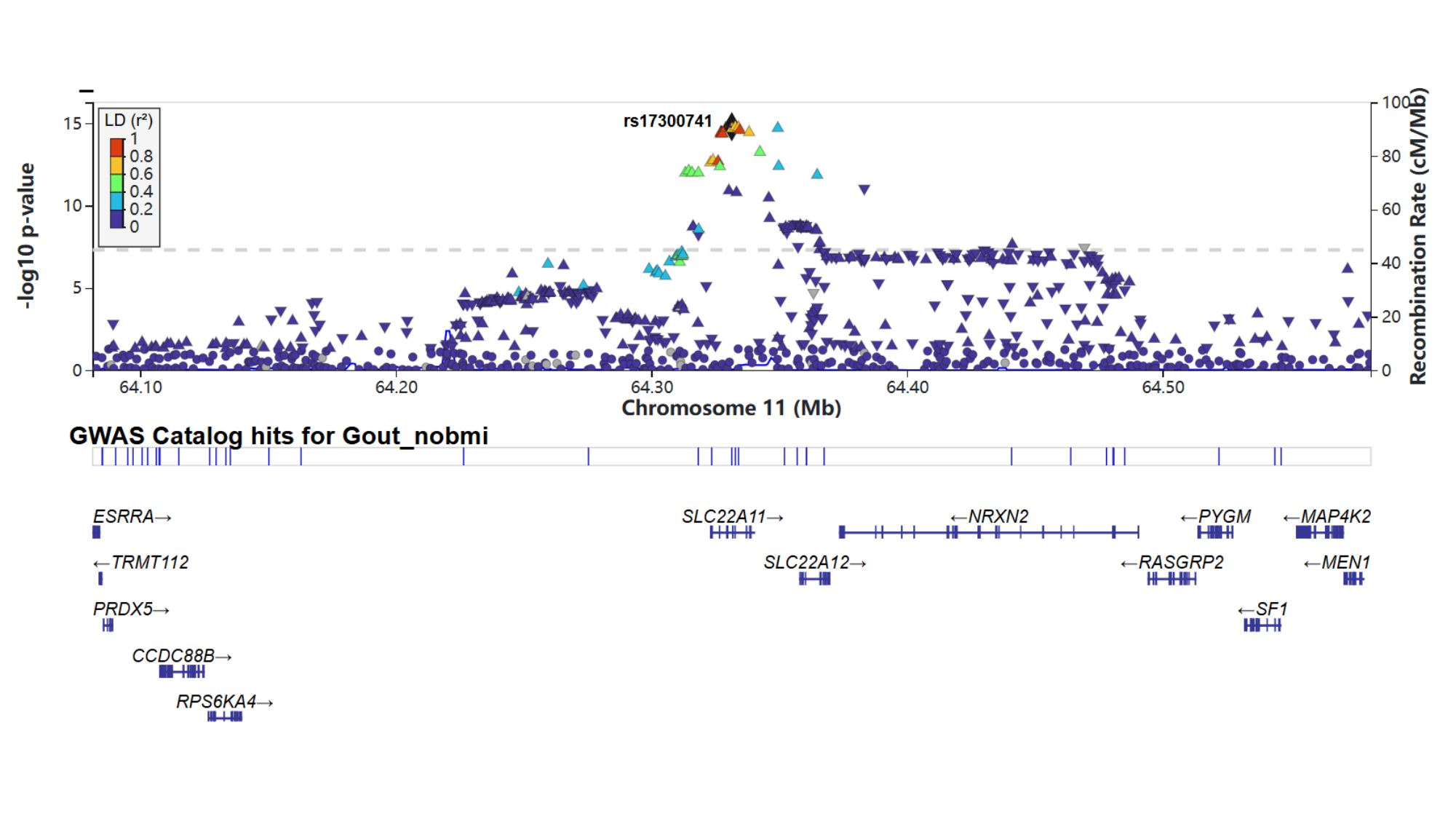

#### Slide 4
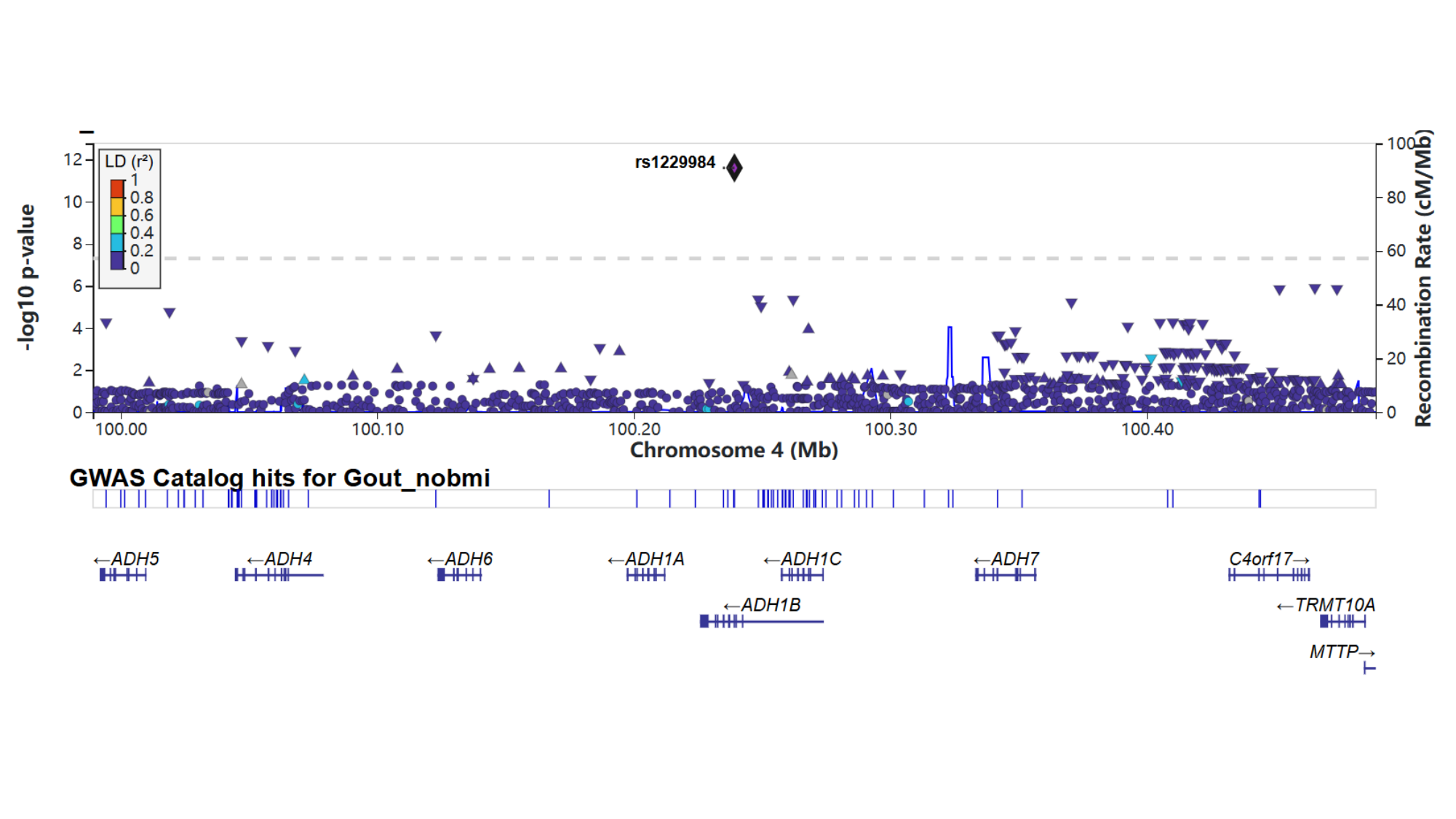

#### Slide 5
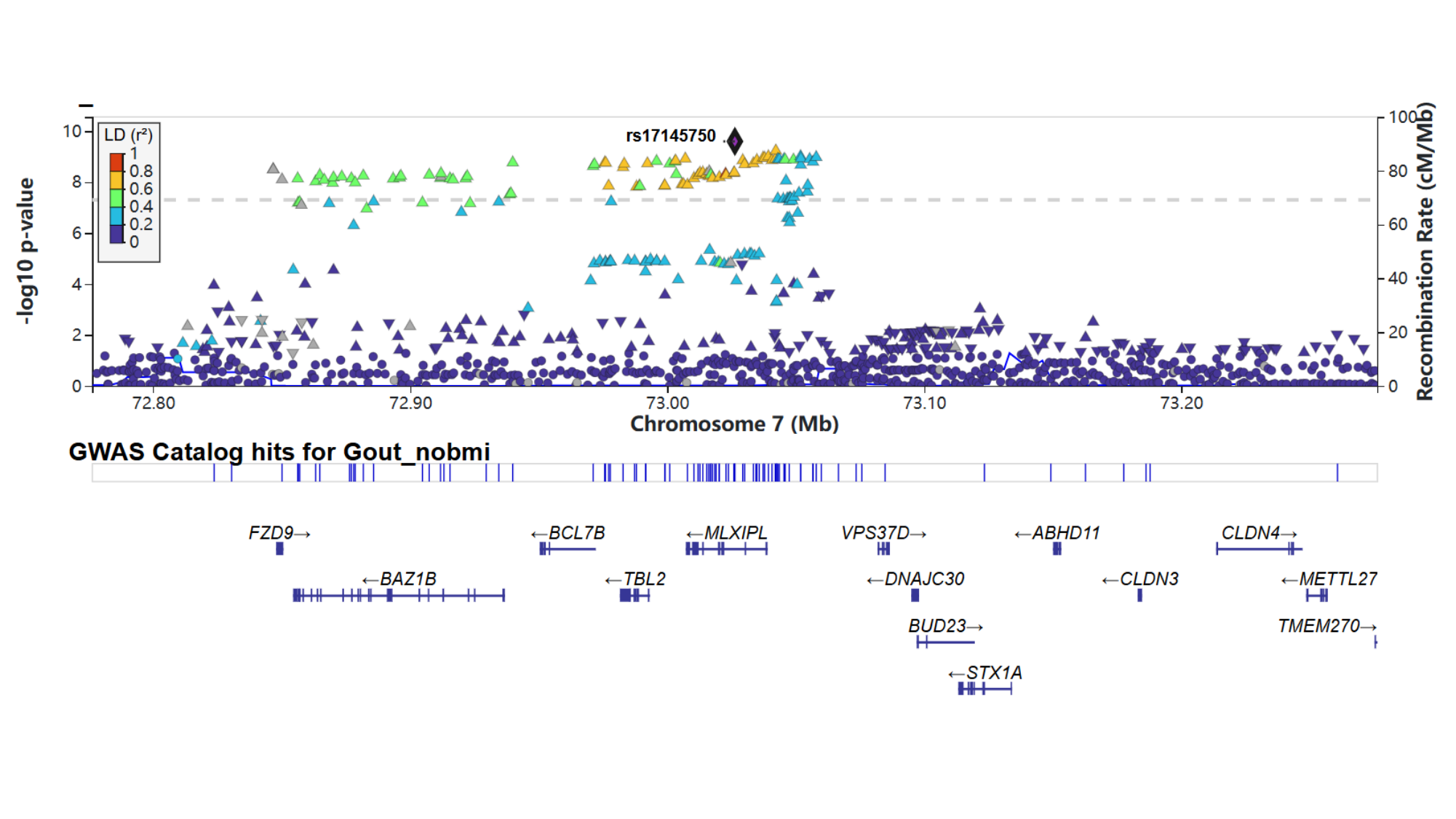

#### Slide 6
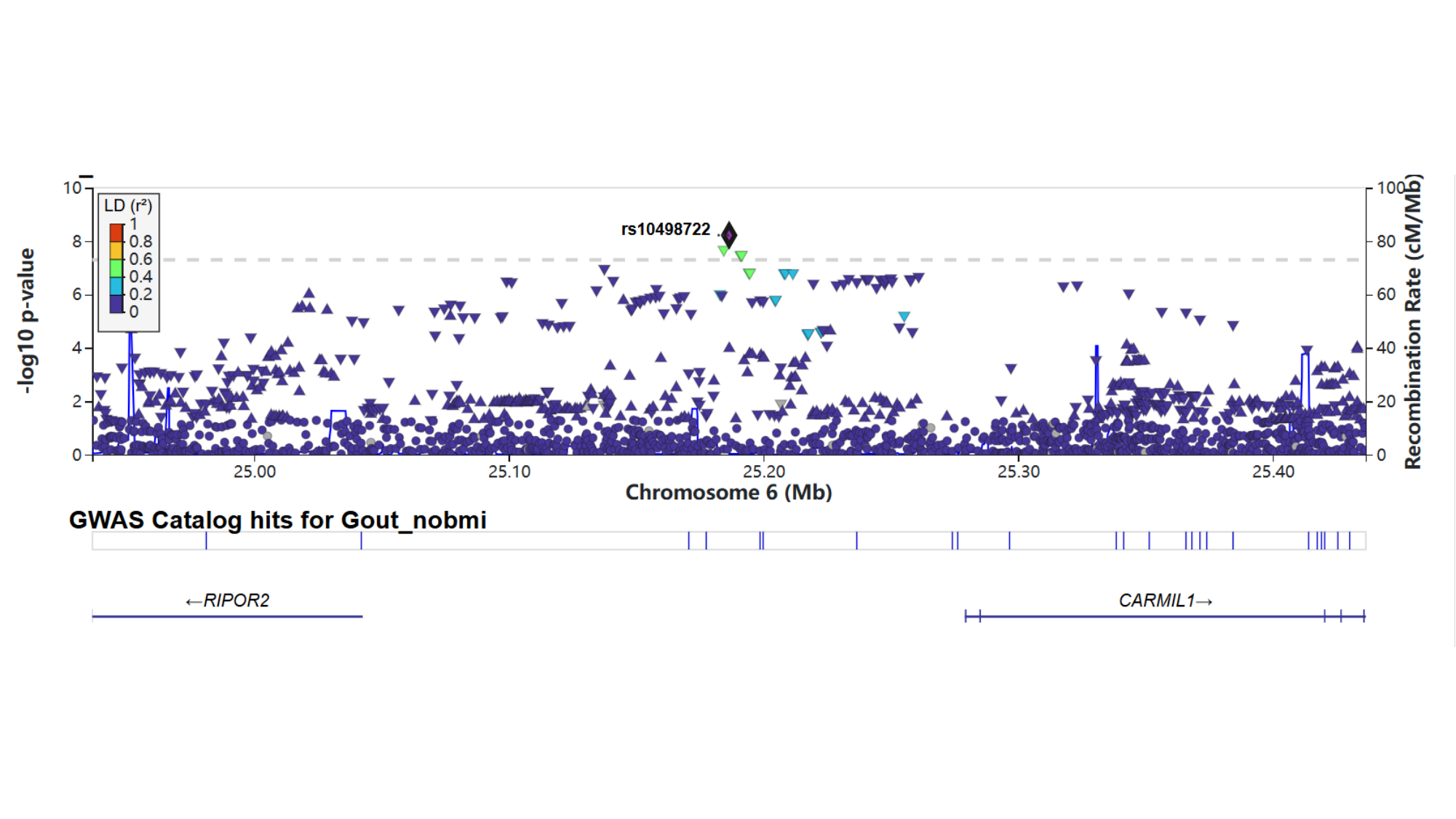

#### Slide 7
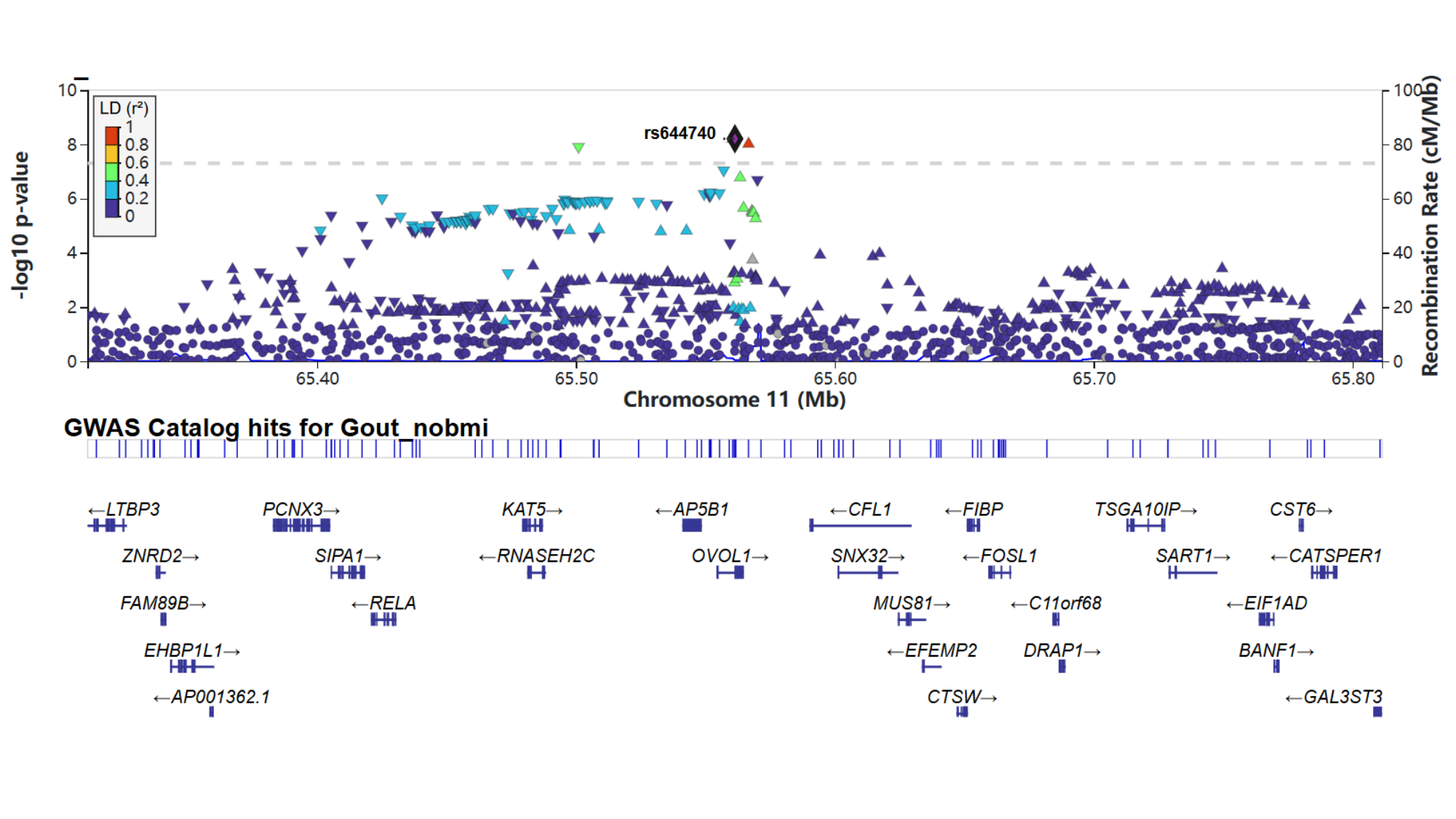

### Supplementary Figure 2

#### Slide 1
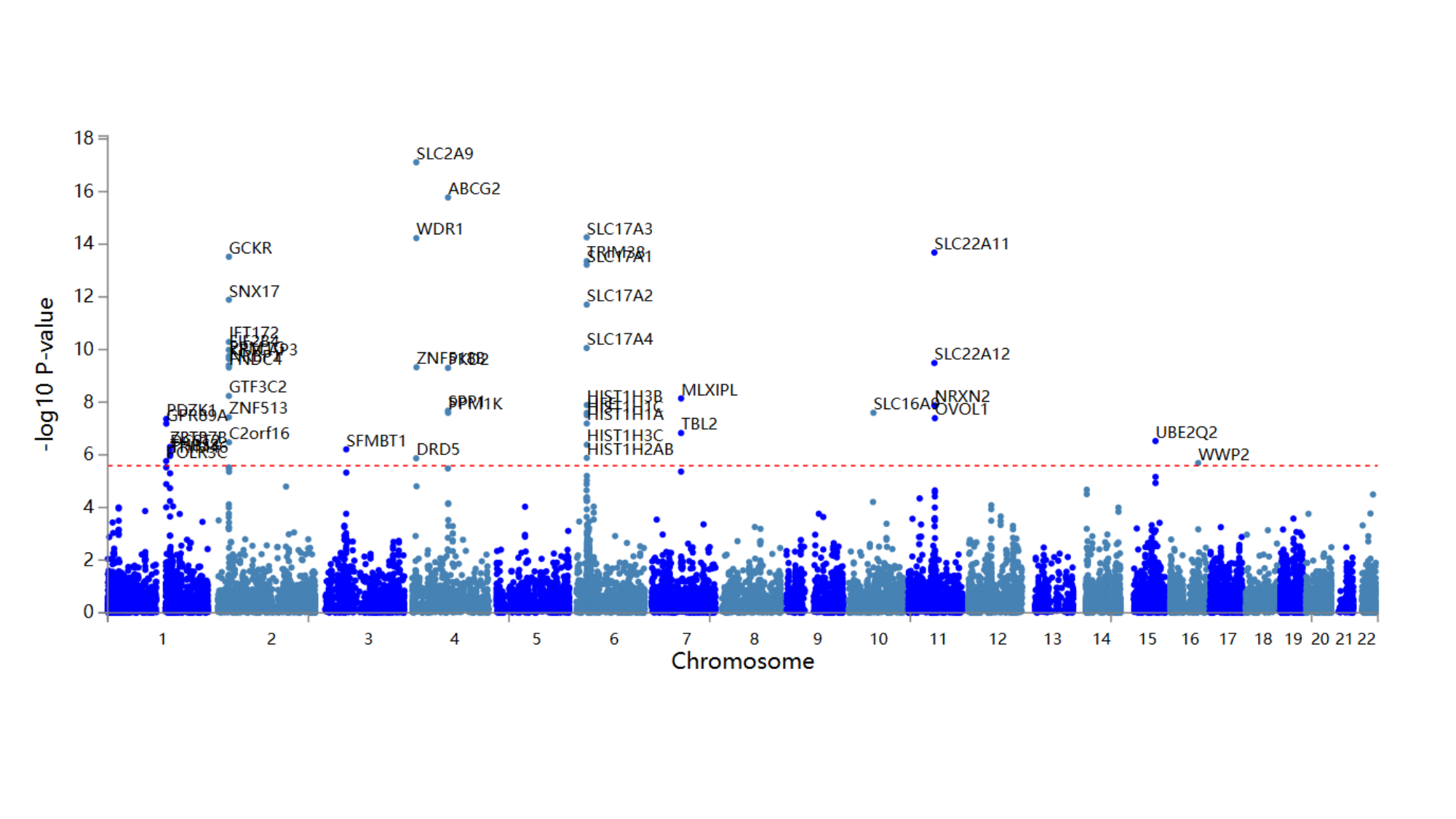

#

### Supplementary Figure 3

#### Slide 1
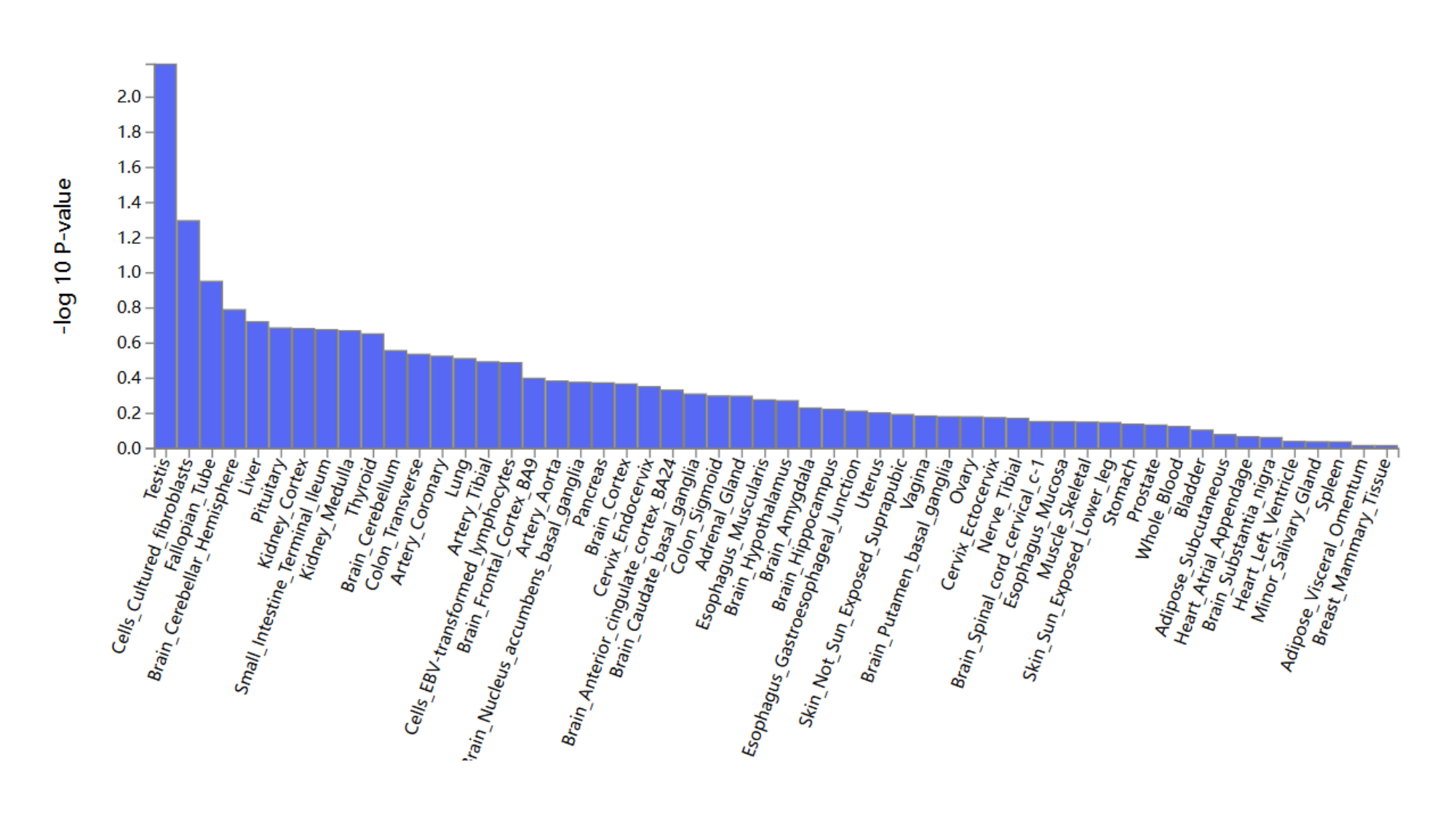

#

### Supplementary Figure 9

#### Slide 1
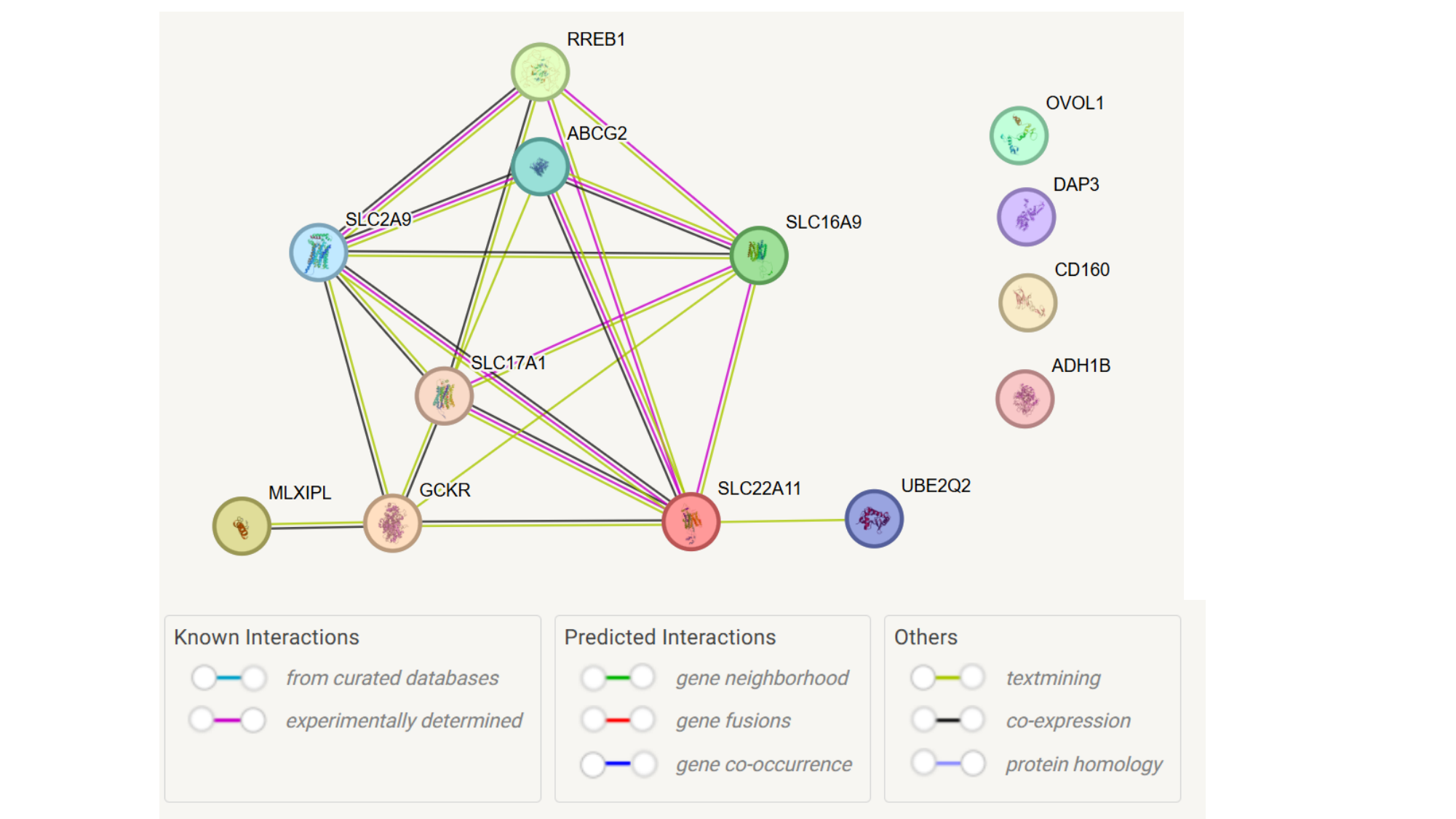
