## Supplementary Figure 4 for "Novel Genetic Variants Associated with Gout Identified Through a Genome-Wide Study in the UK Biobank (N = 150,542)"

### Slide 1
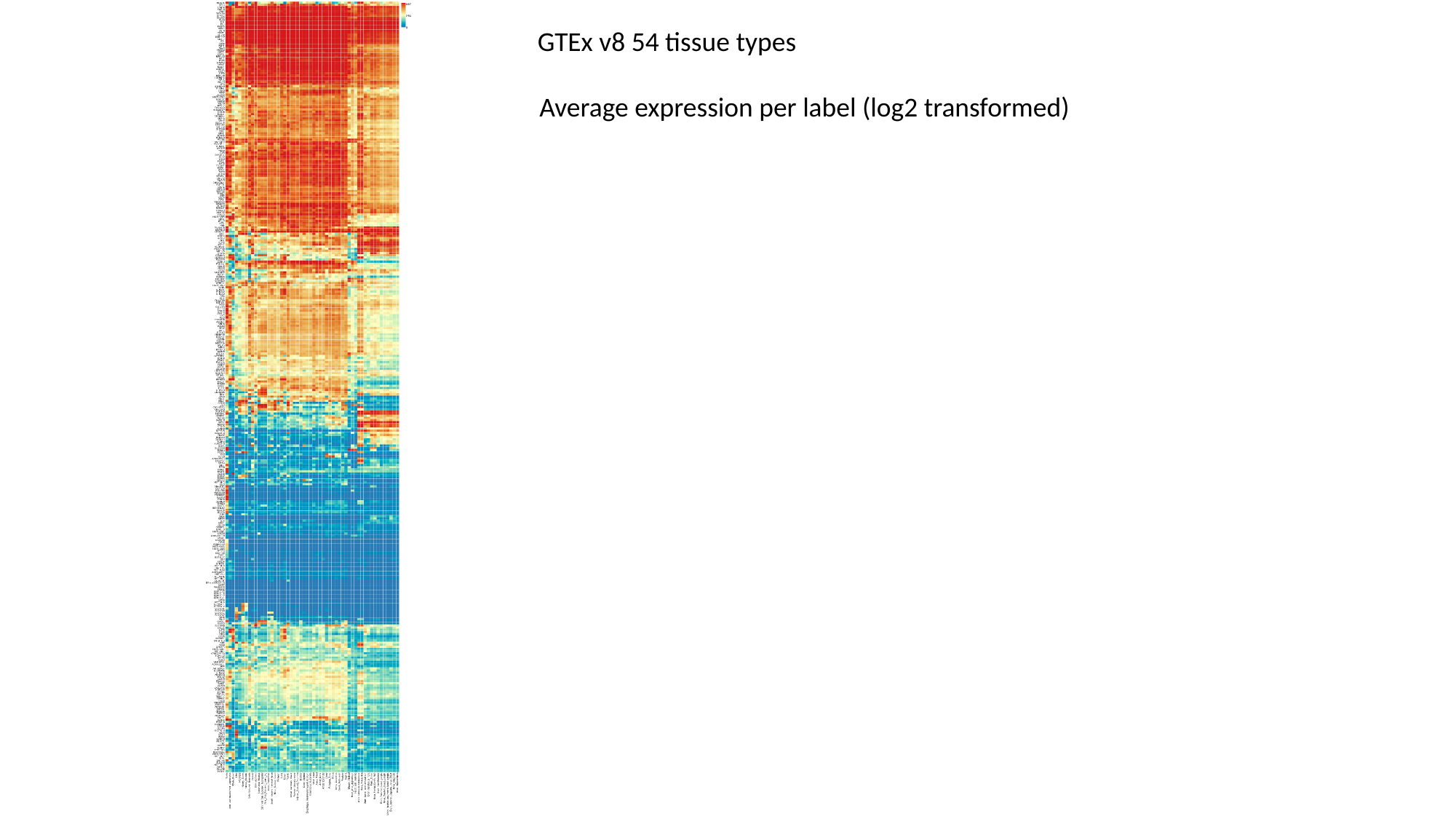

GTEx v8 54 tissue types
Average expression per label (log2 transformed)

### Slide 2
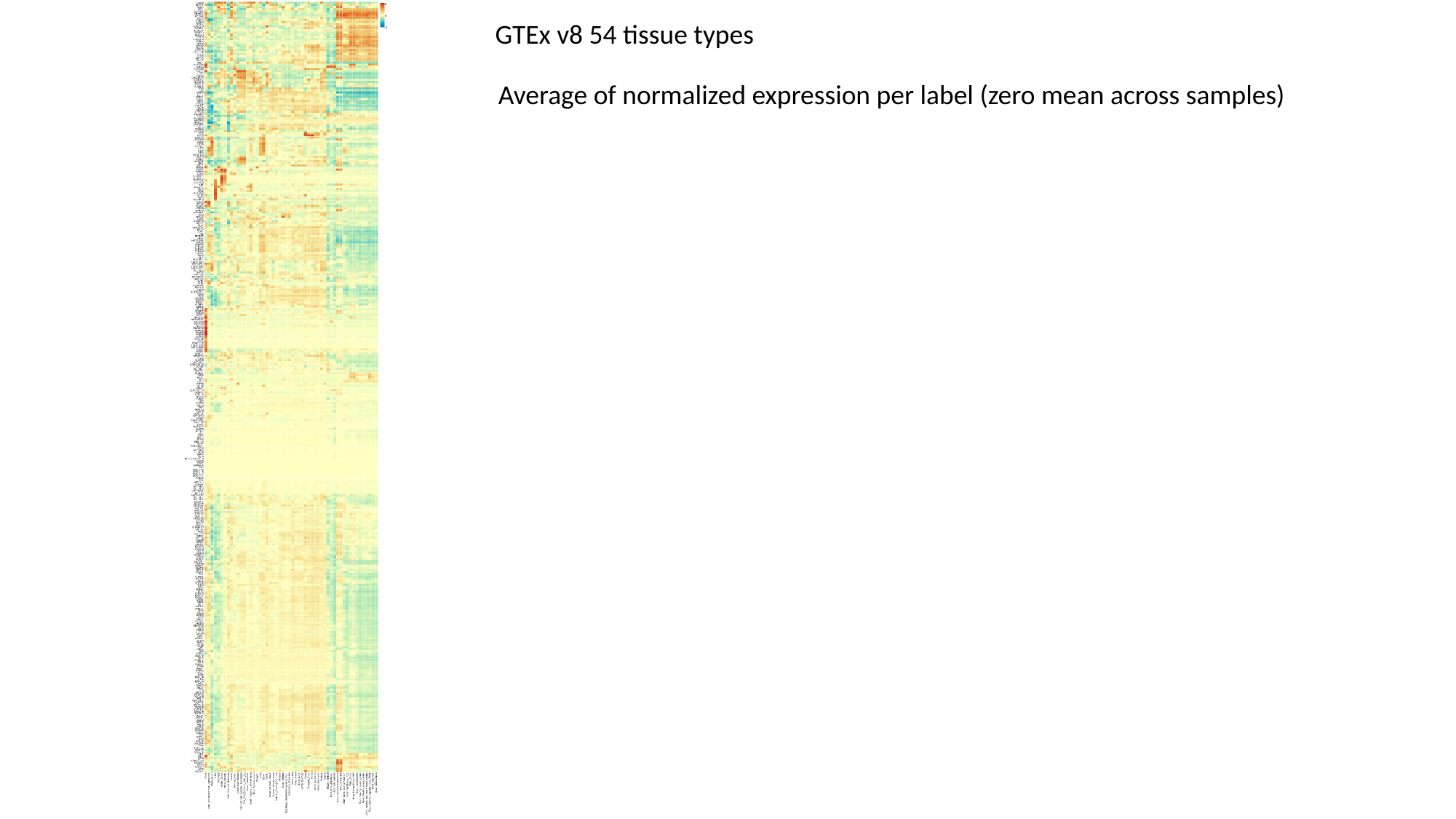

GTEx v8 54 tissue types
Average of normalized expression per label (zero mean across samples)

### Slide 3
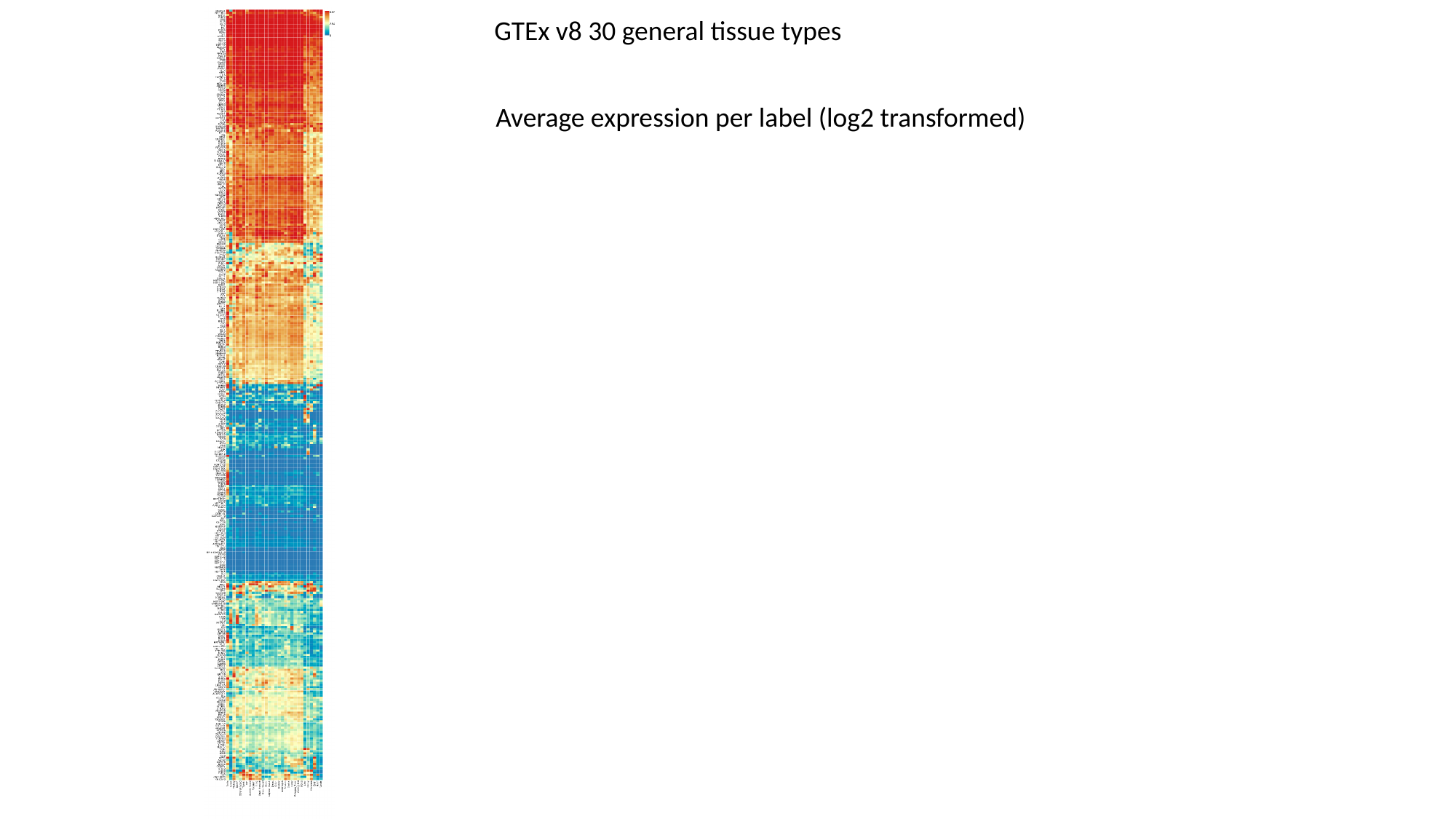

GTEx v8 30 general tissue types
Average expression per label (log2 transformed)

### Slide 4
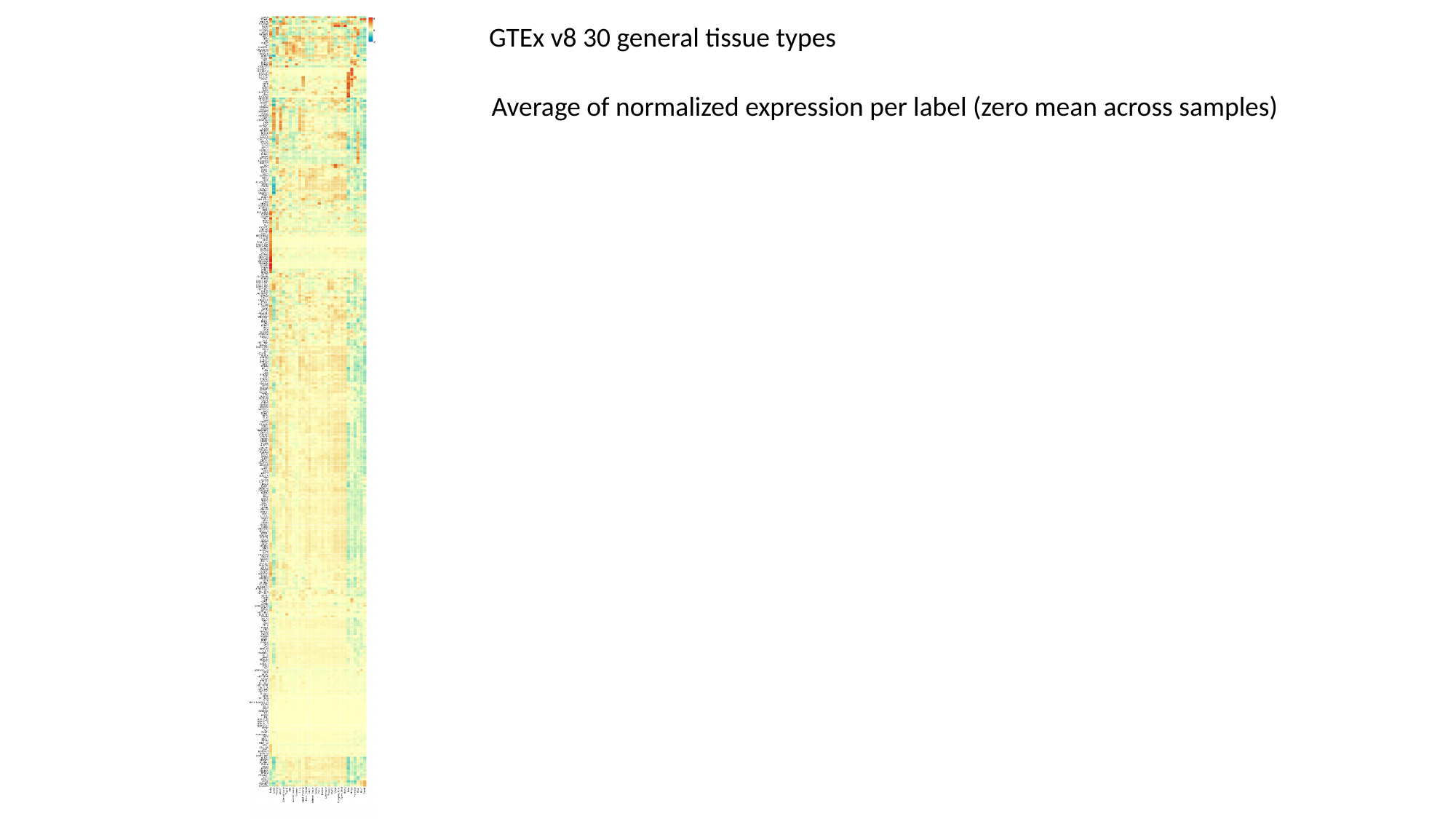

GTEx v8 30 general tissue types
Average of normalized expression per label (zero mean across samples)
