## Supplementary Figure 5 for "Novel Genetic Variants Associated with Gout Identified Through a Genome-Wide Study in the UK Biobank (N = 150,542)"

### Slide 1
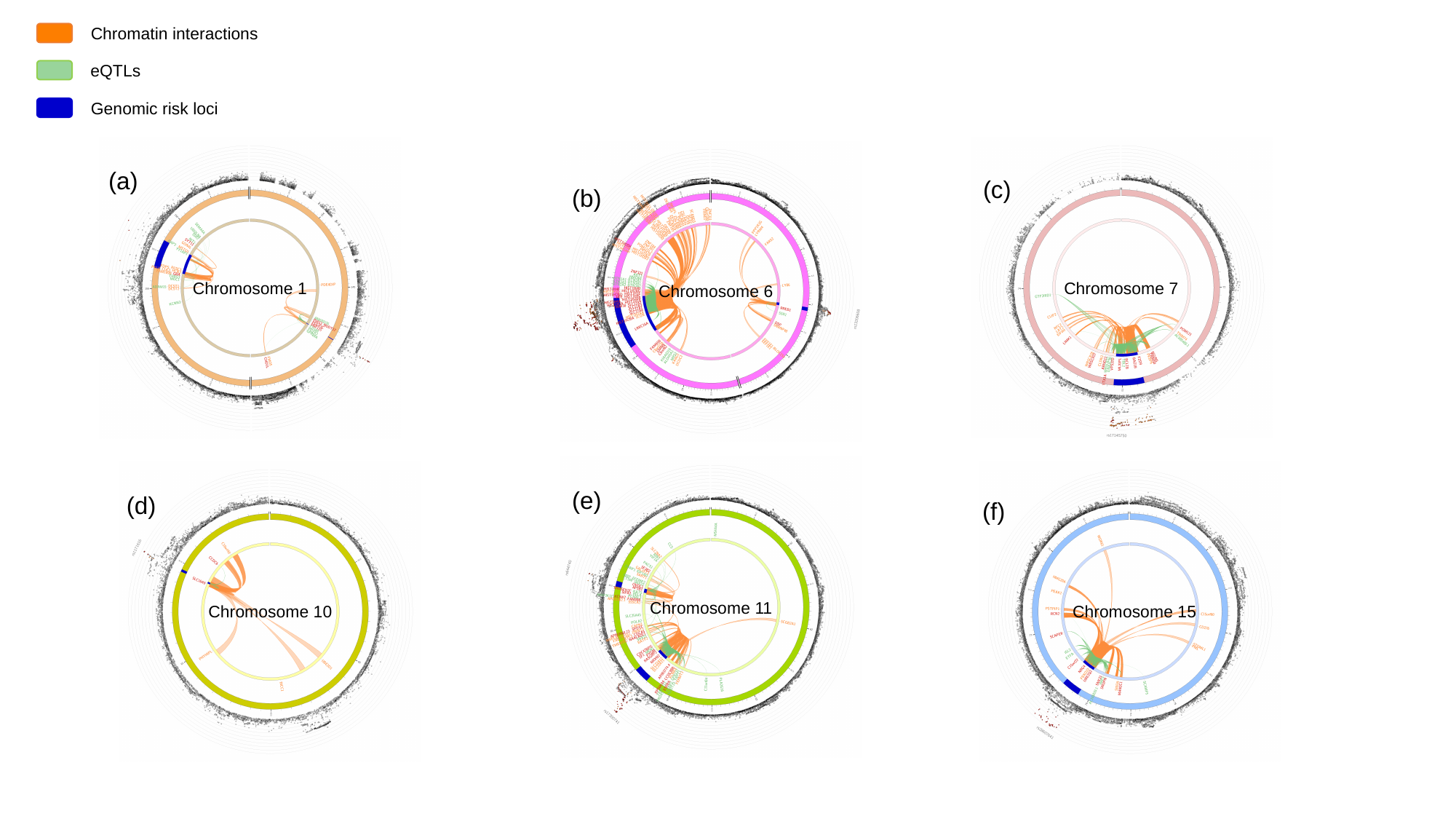

Chromatin interactions
eQTLs
Genomic risk loci
(c)
Chromosome 7
(a)
Chromosome 1
(b)
Chromosome 6
(e)
Chromosome 11
(d)
Chromosome 10
(f)
Chromosome 15
