## Supplementary Figure 6 for "Novel Genetic Variants Associated with Gout Identified Through a Genome-Wide Study in the UK Biobank (N = 150,542)"

### Slide 1
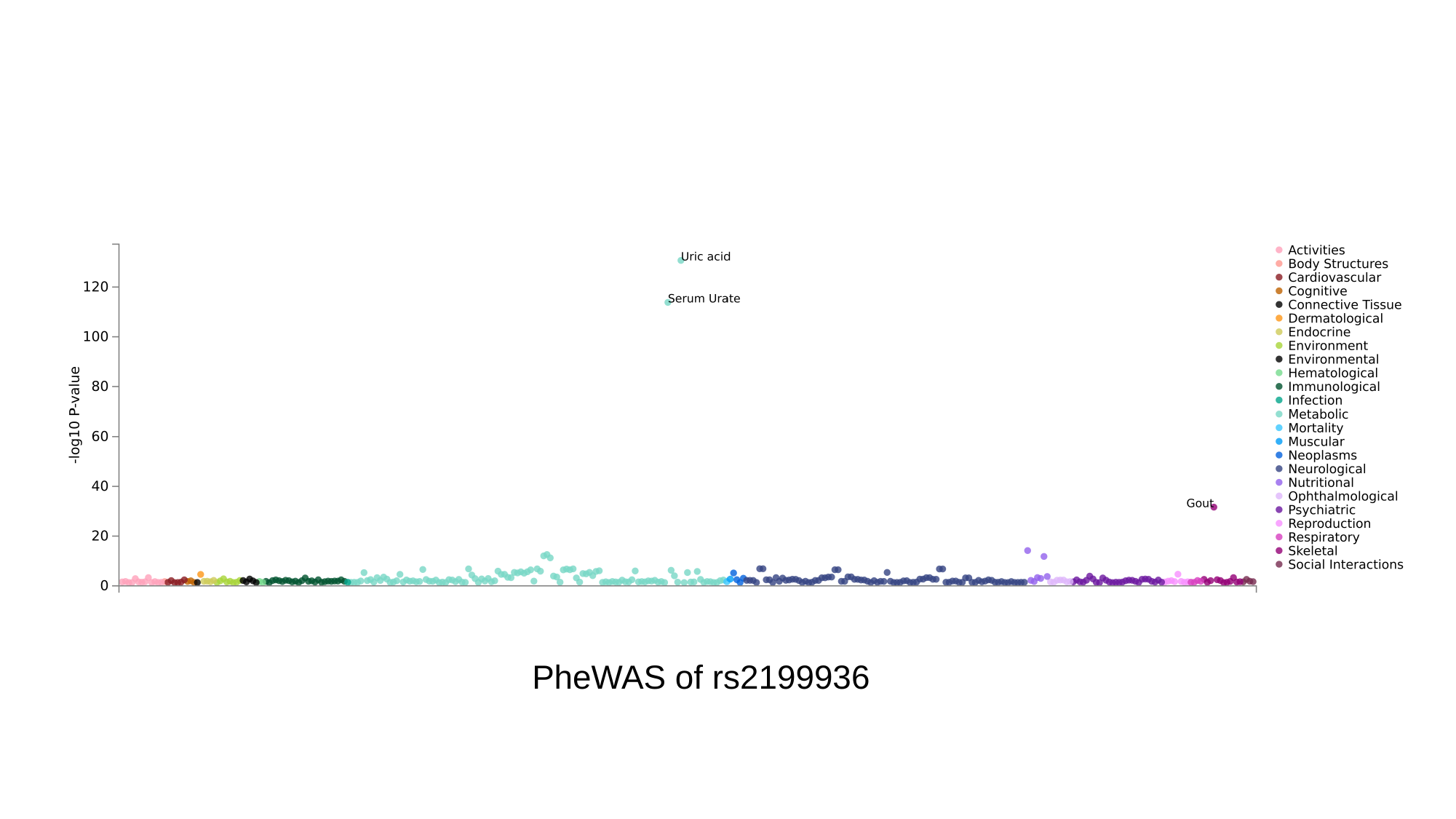

PheWAS of rs2199936

### Slide 2
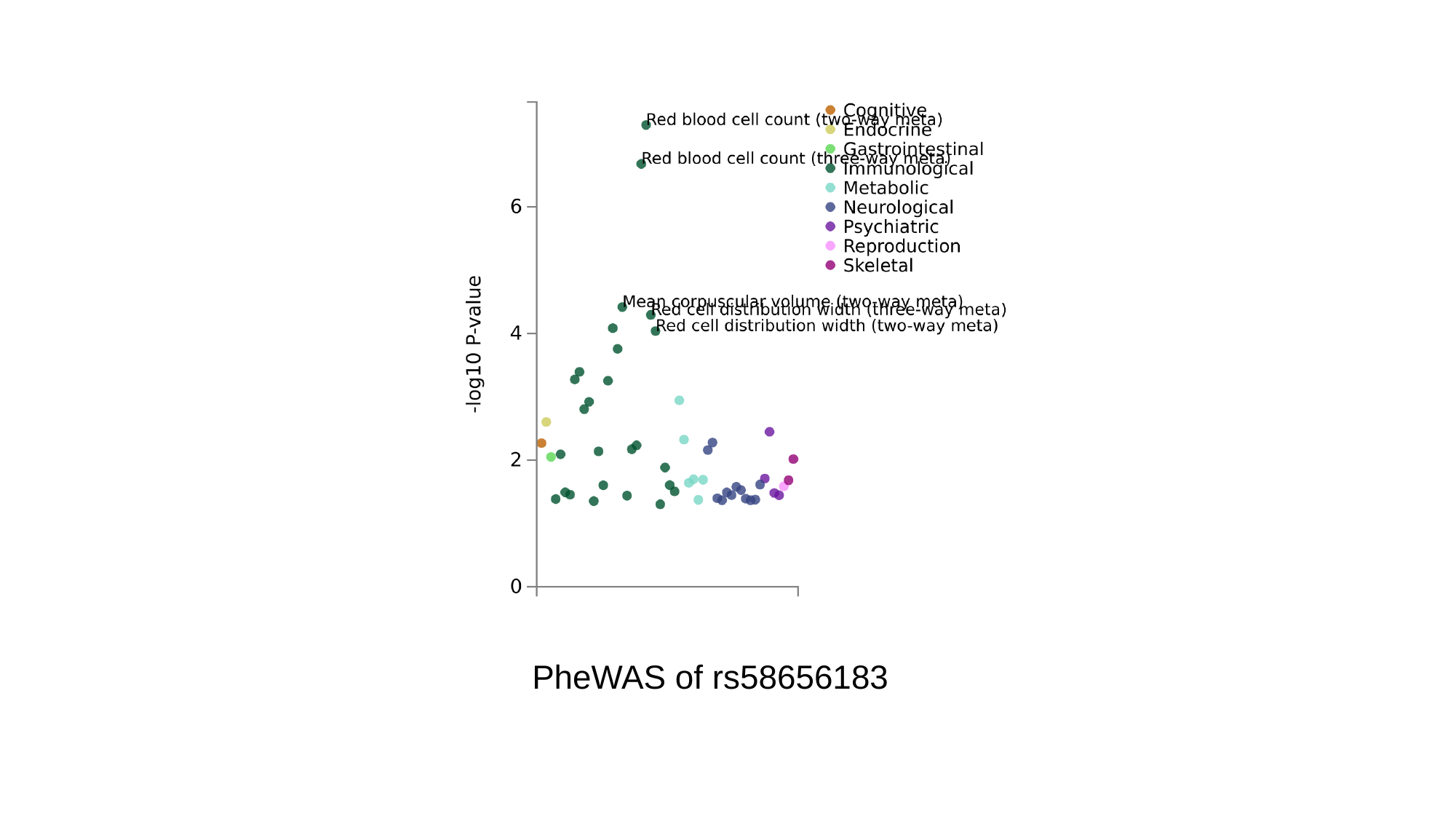

PheWAS of rs58656183

### Slide 3
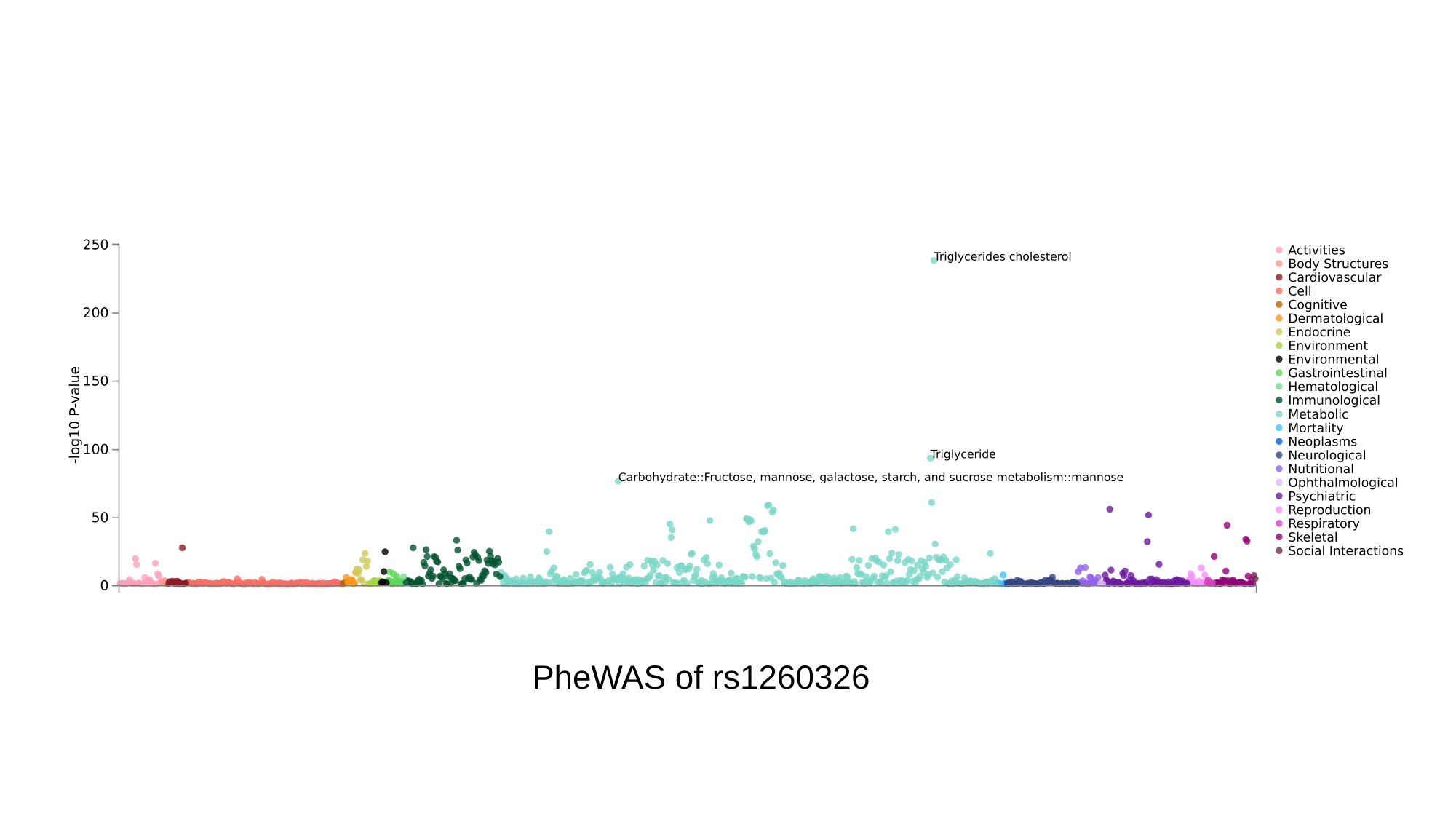

PheWAS of rs1260326

### Slide 4
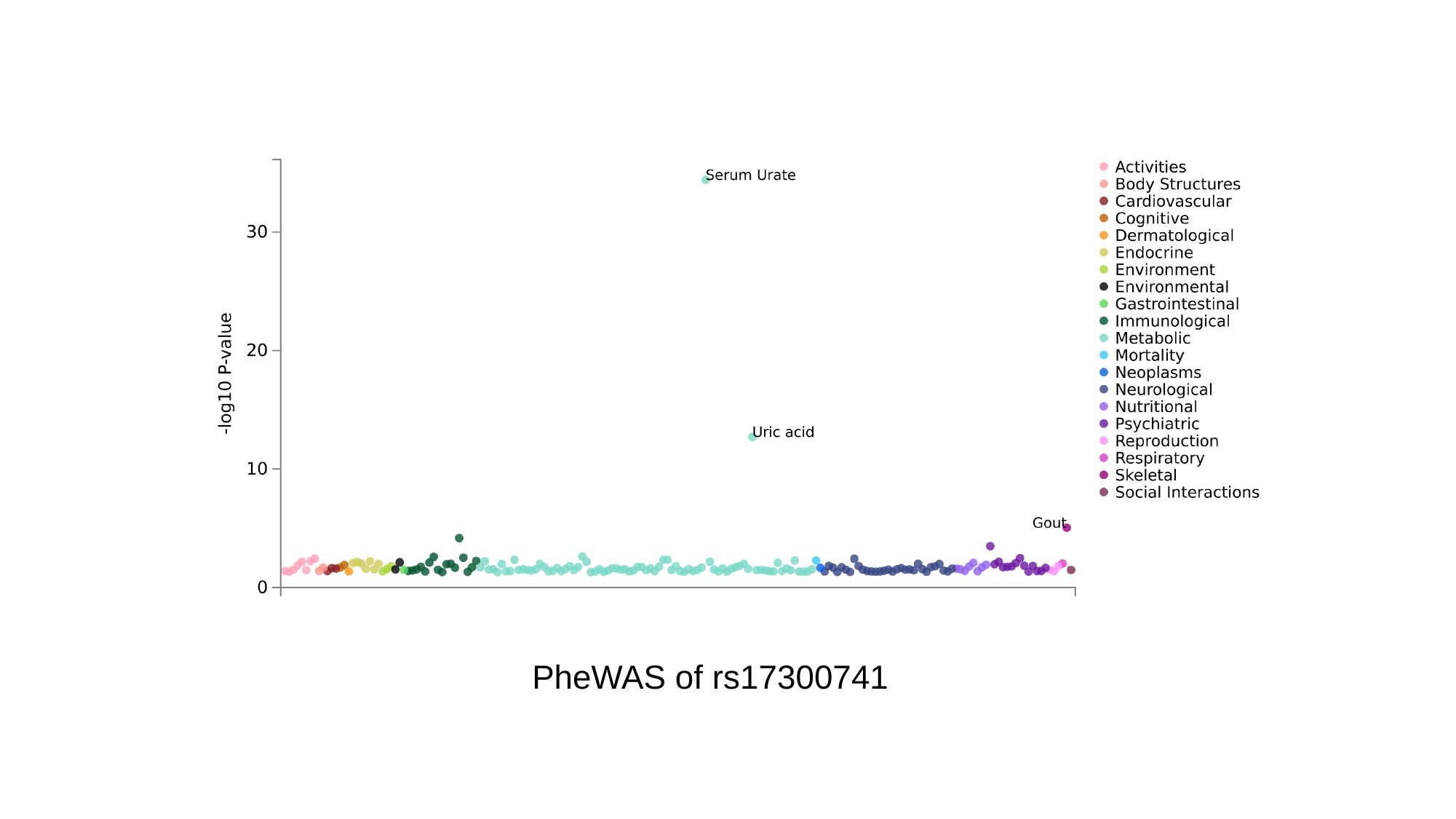

PheWAS of rs17300741

### Slide 5
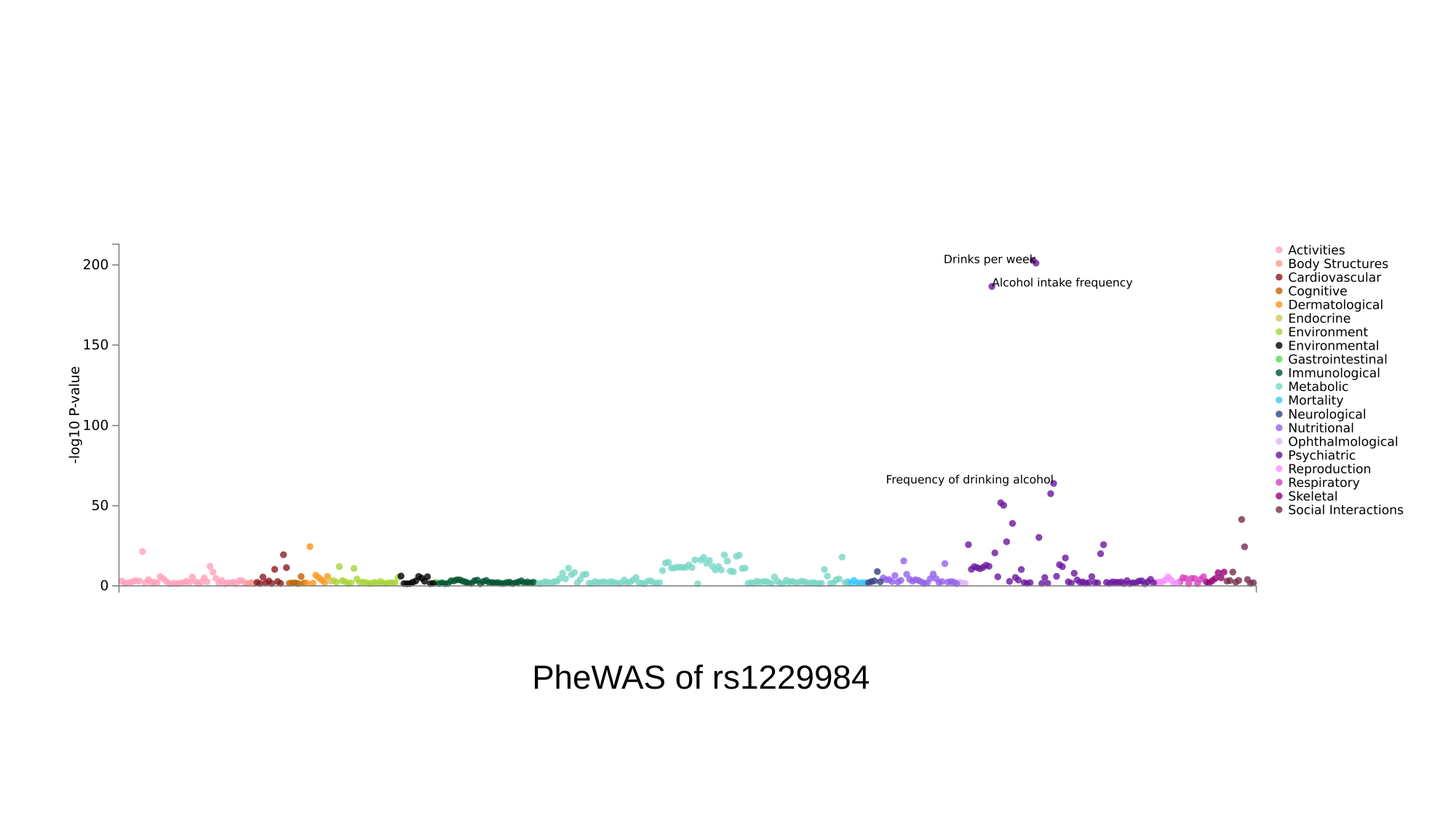

PheWAS of rs1229984

### Slide 6
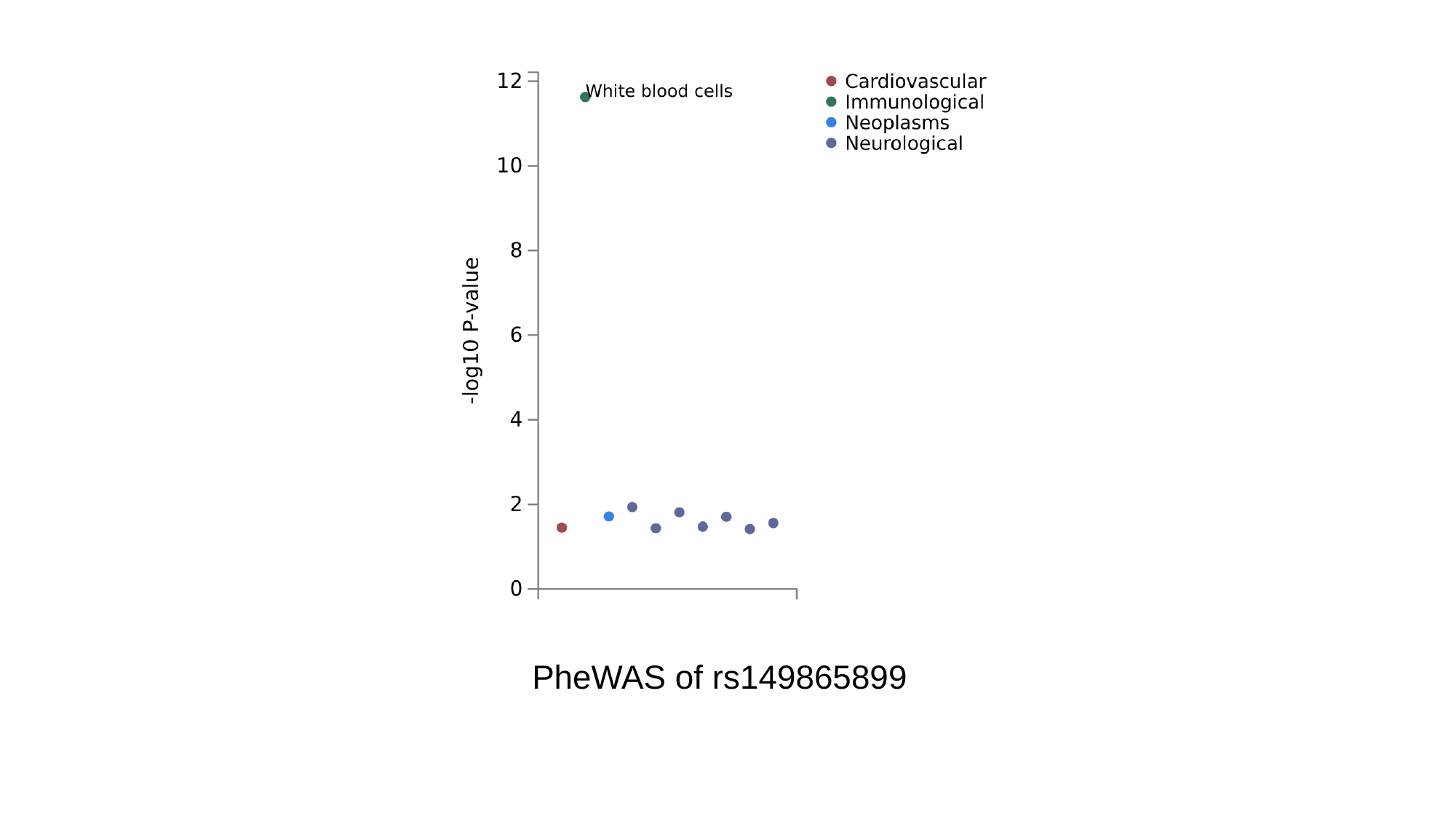

PheWAS of rs149865899

### Slide 7
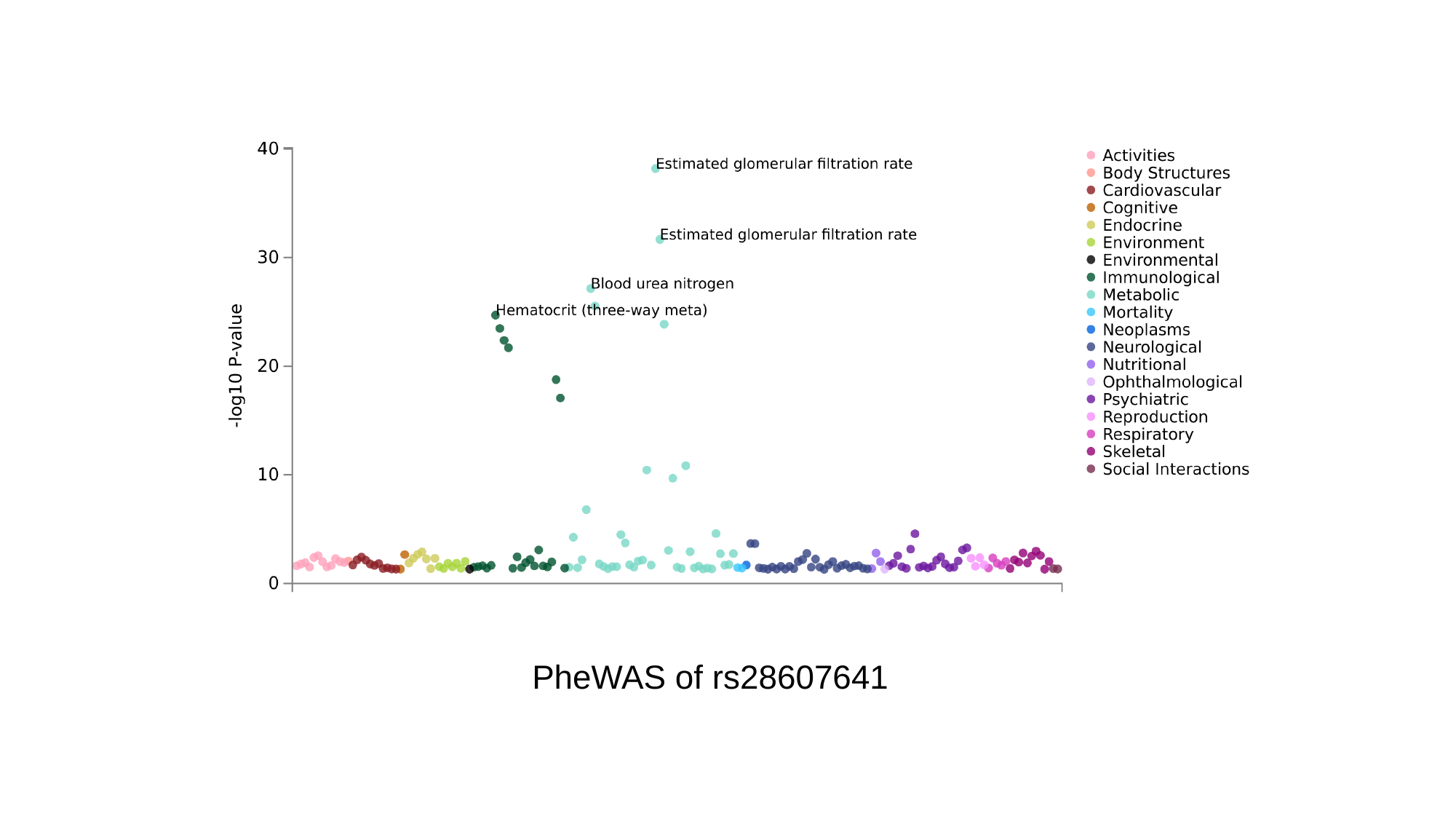

PheWAS of rs28607641

### Slide 8
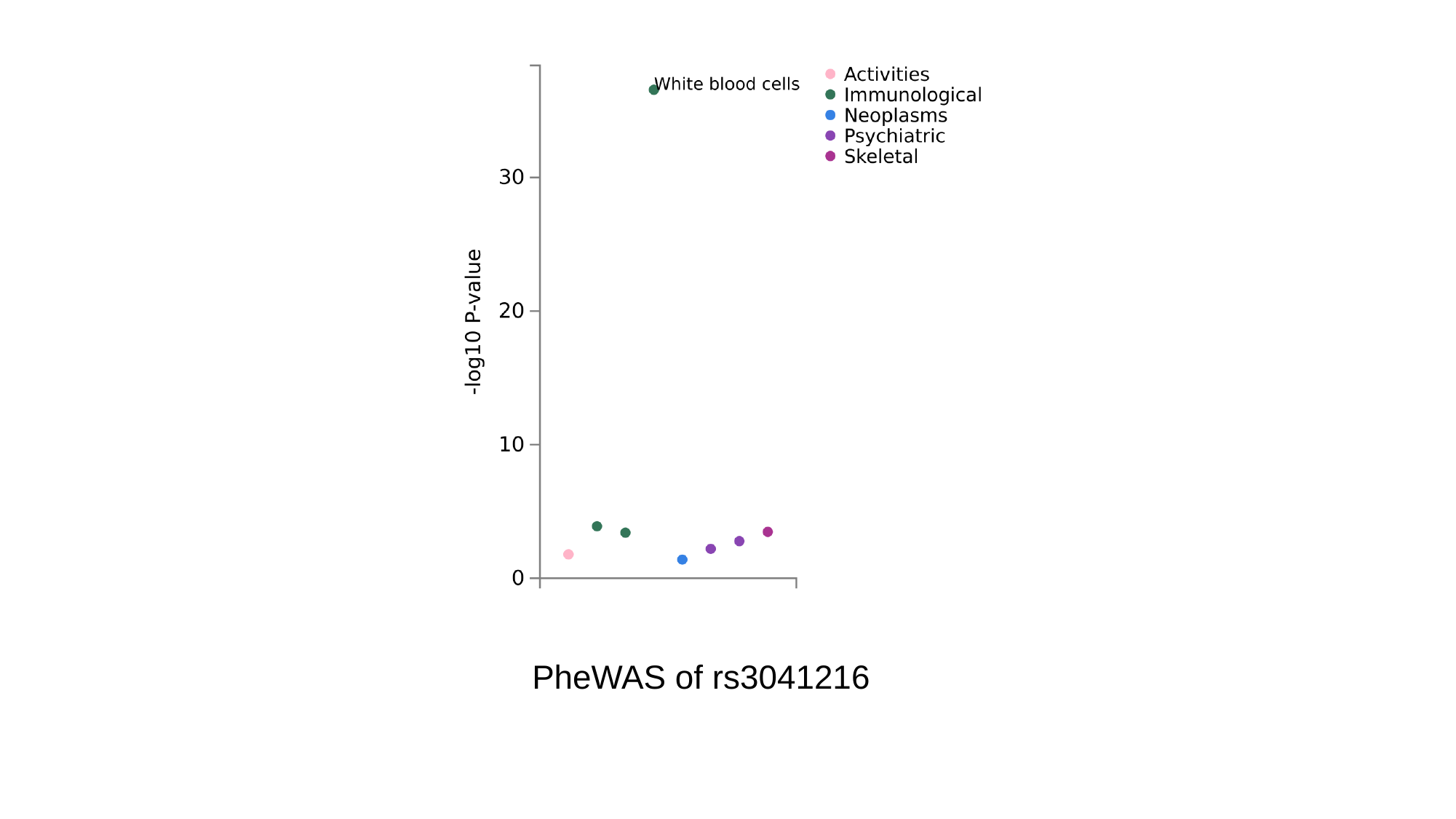

PheWAS of rs3041216

### Slide 9
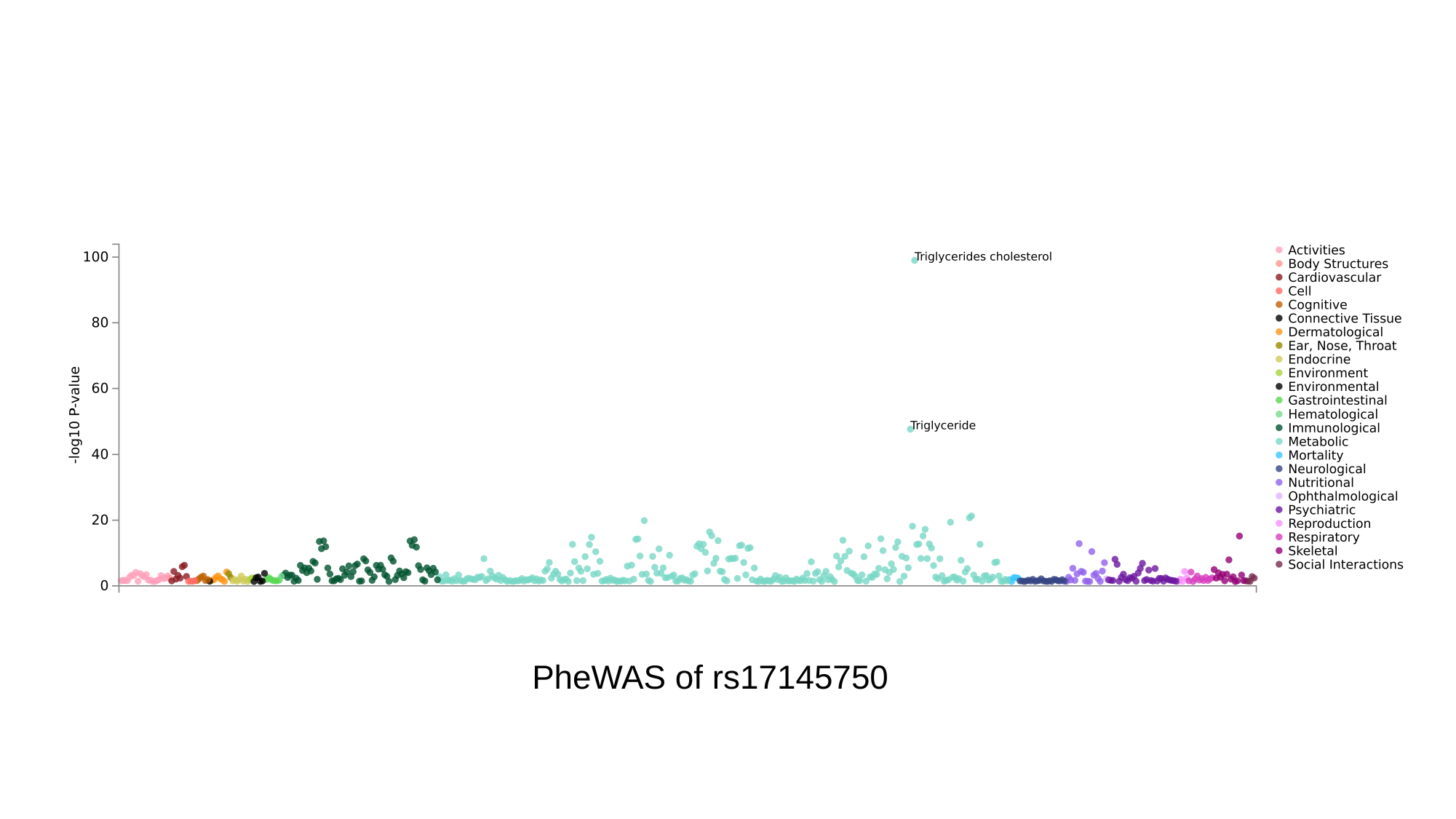

PheWAS of rs17145750

### Slide 10
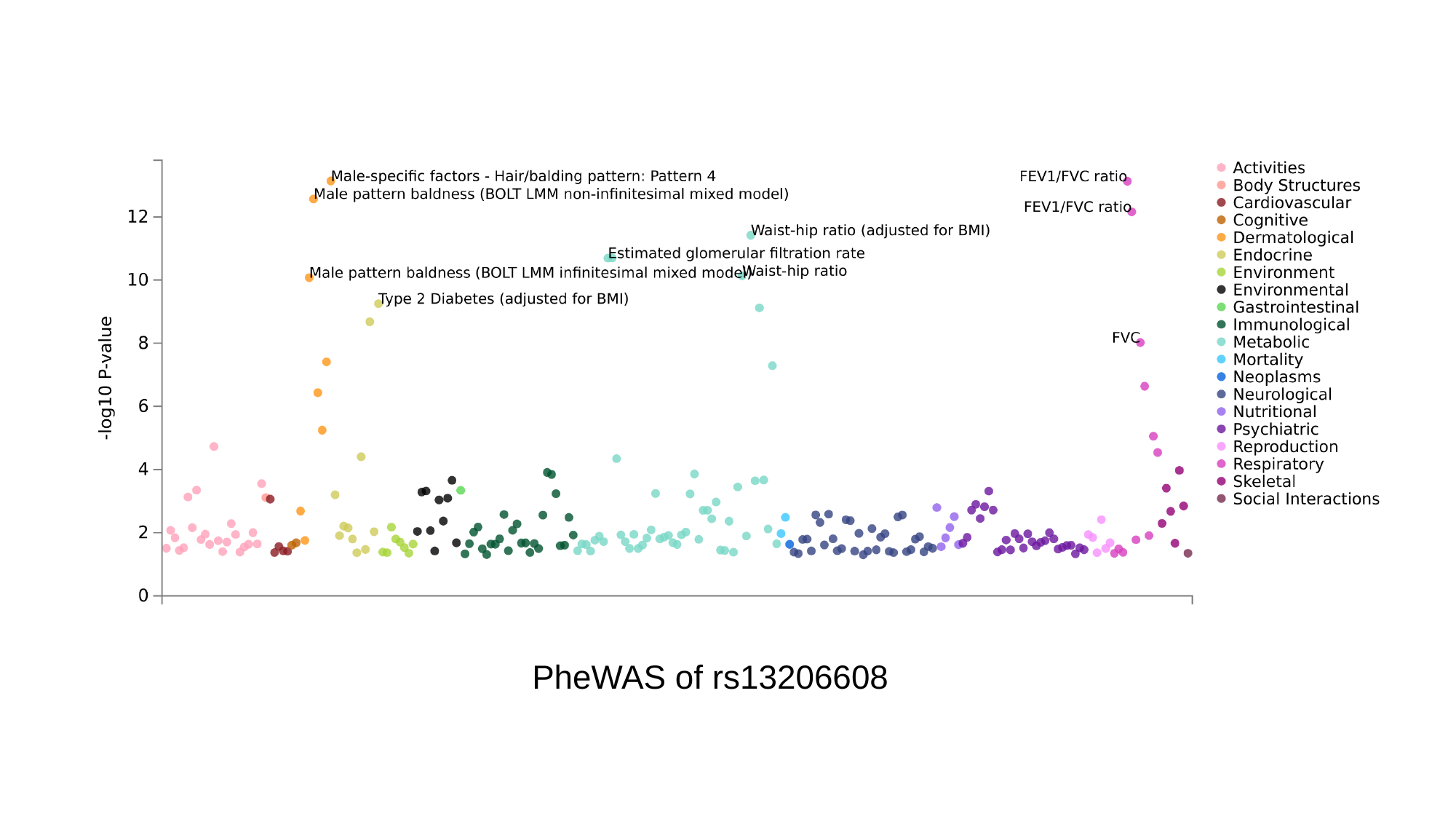

PheWAS of rs13206608

### Slide 11
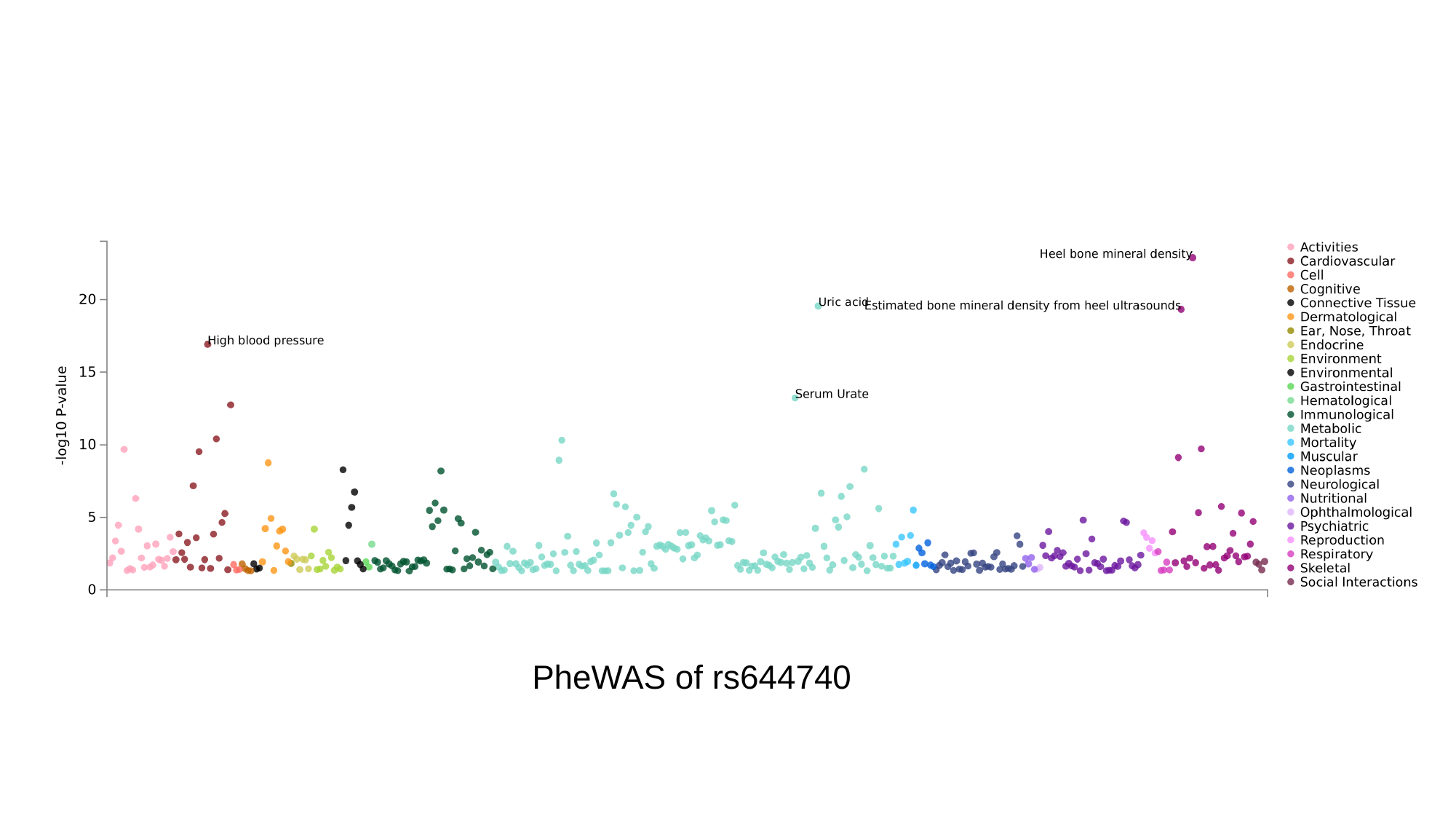

PheWAS of rs644740

### Slide 12
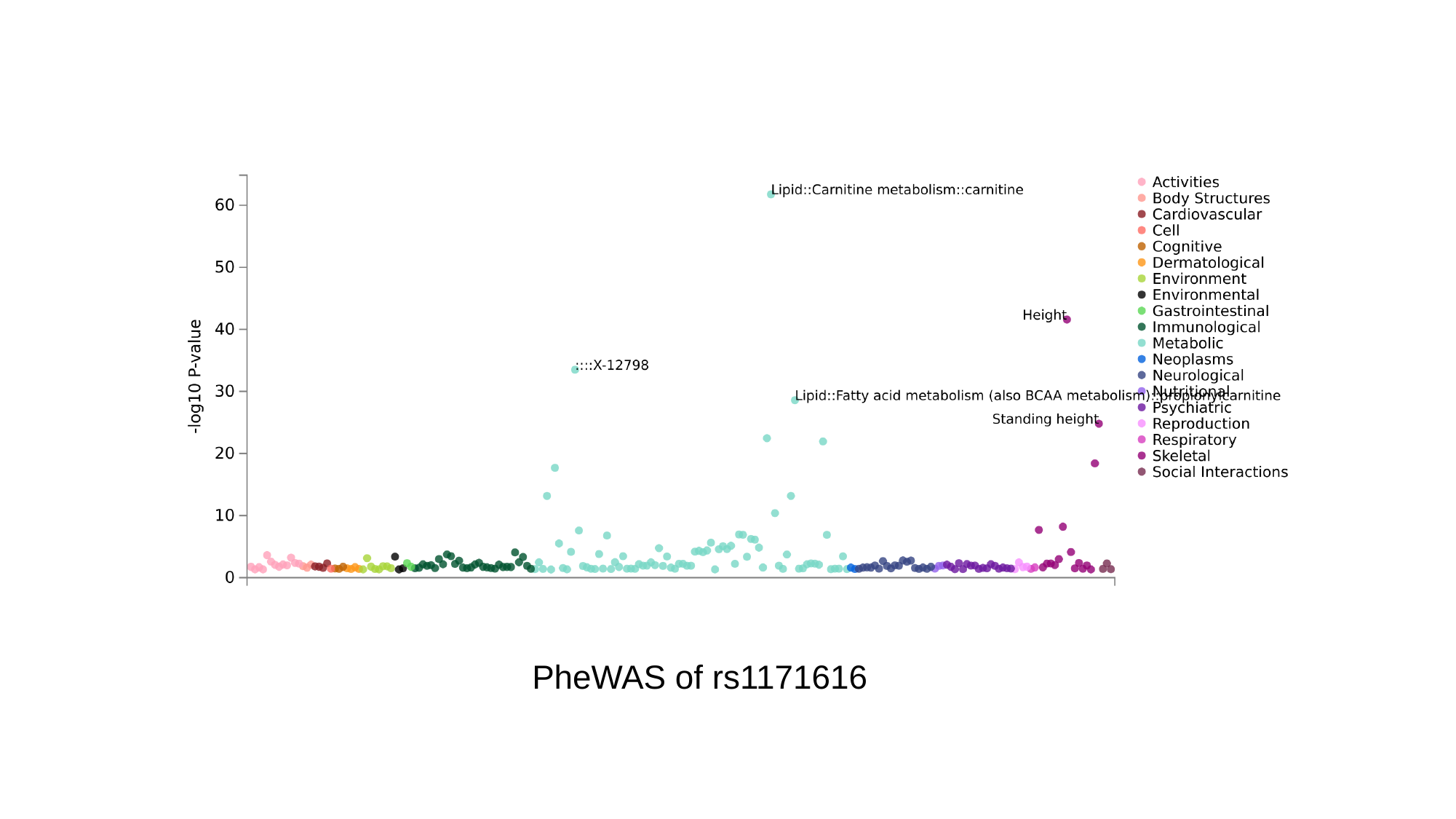

PheWAS of rs1171616
