## Supplementary Figure 7 for "Novel Genetic Variants Associated with Gout Identified Through a Genome-Wide Study in the UK Biobank (N = 150,542)"

### Slide 1
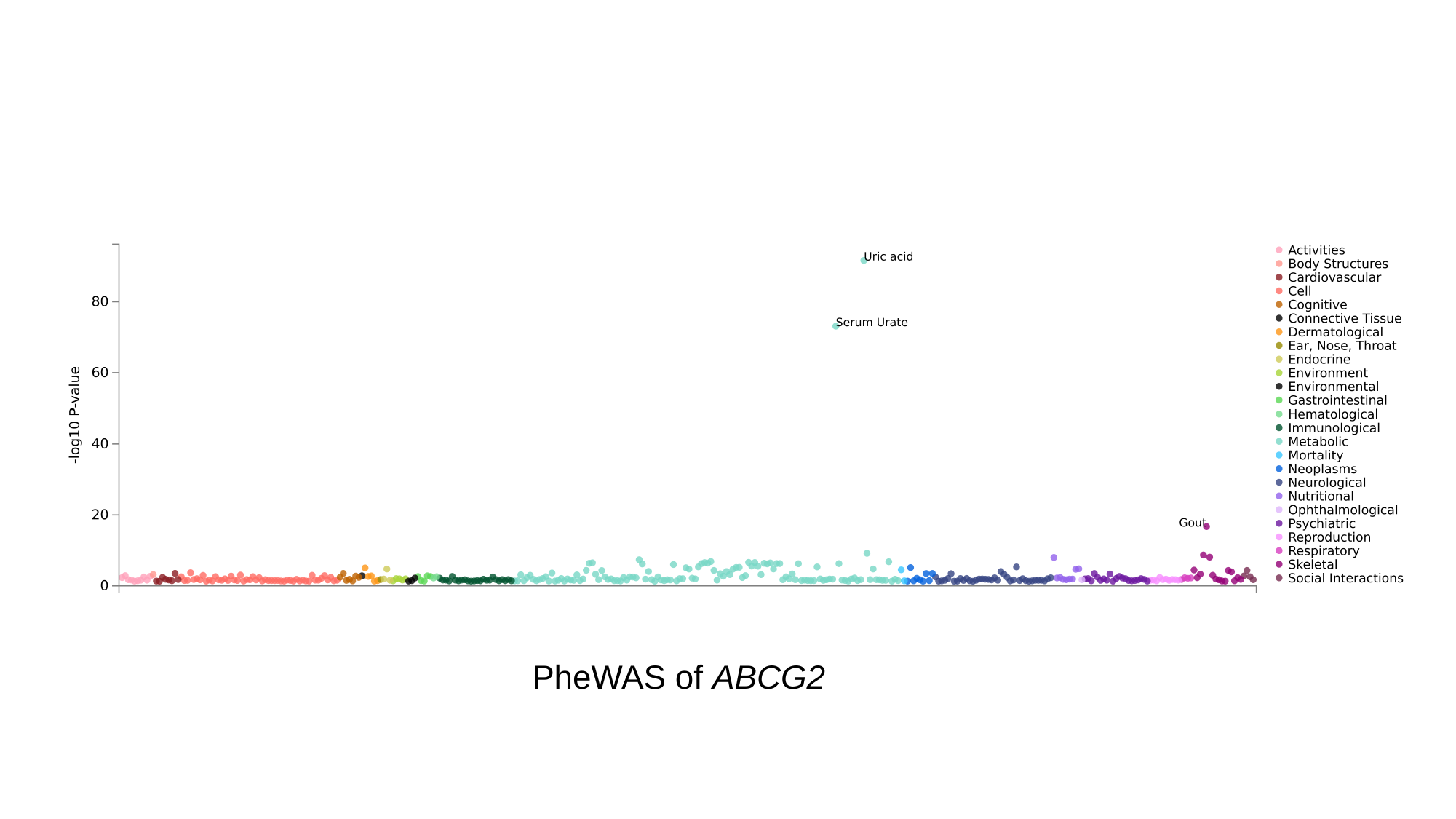

PheWAS of ABCG2

### Slide 2
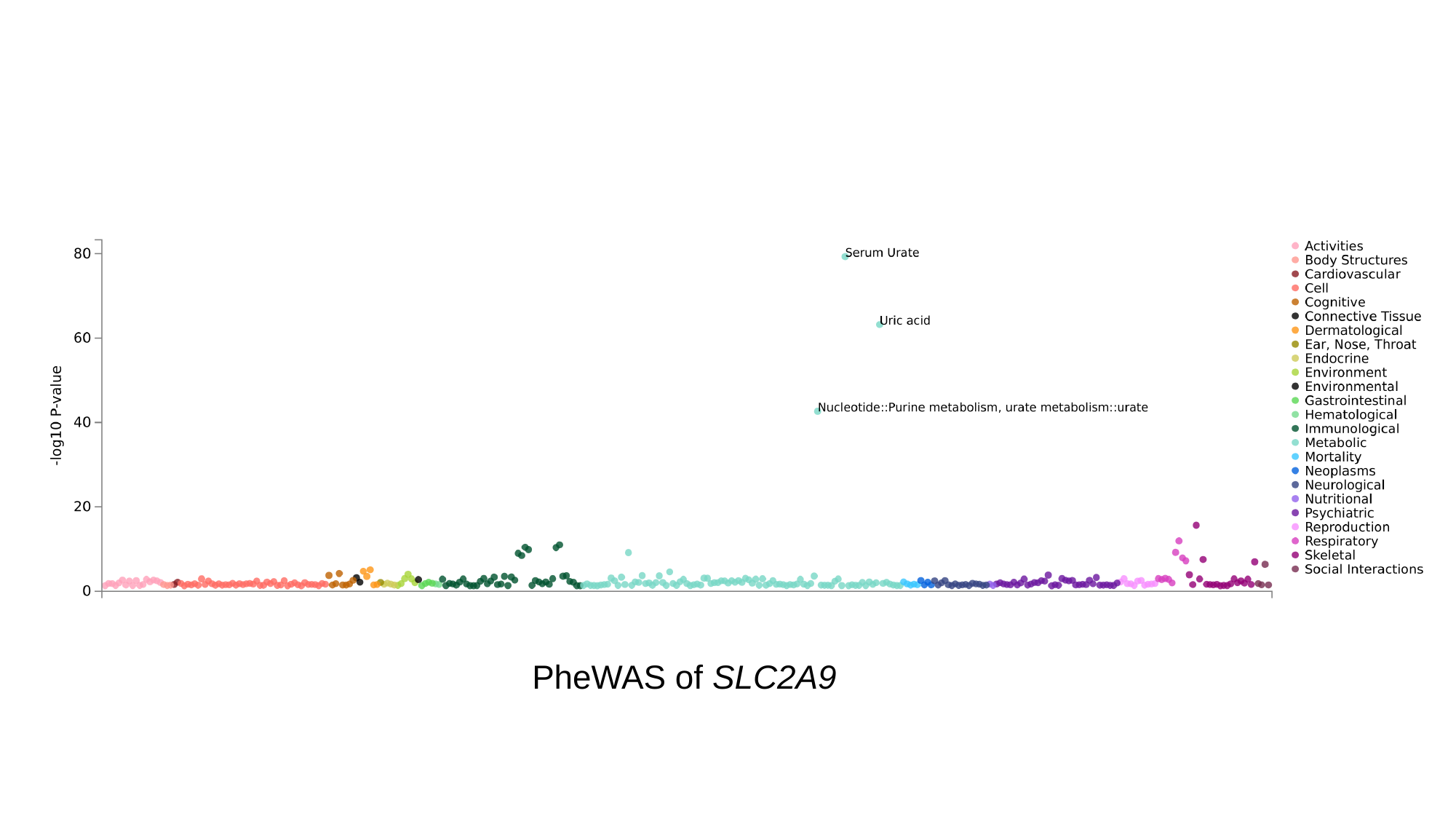

PheWAS of SLC2A9

### Slide 3
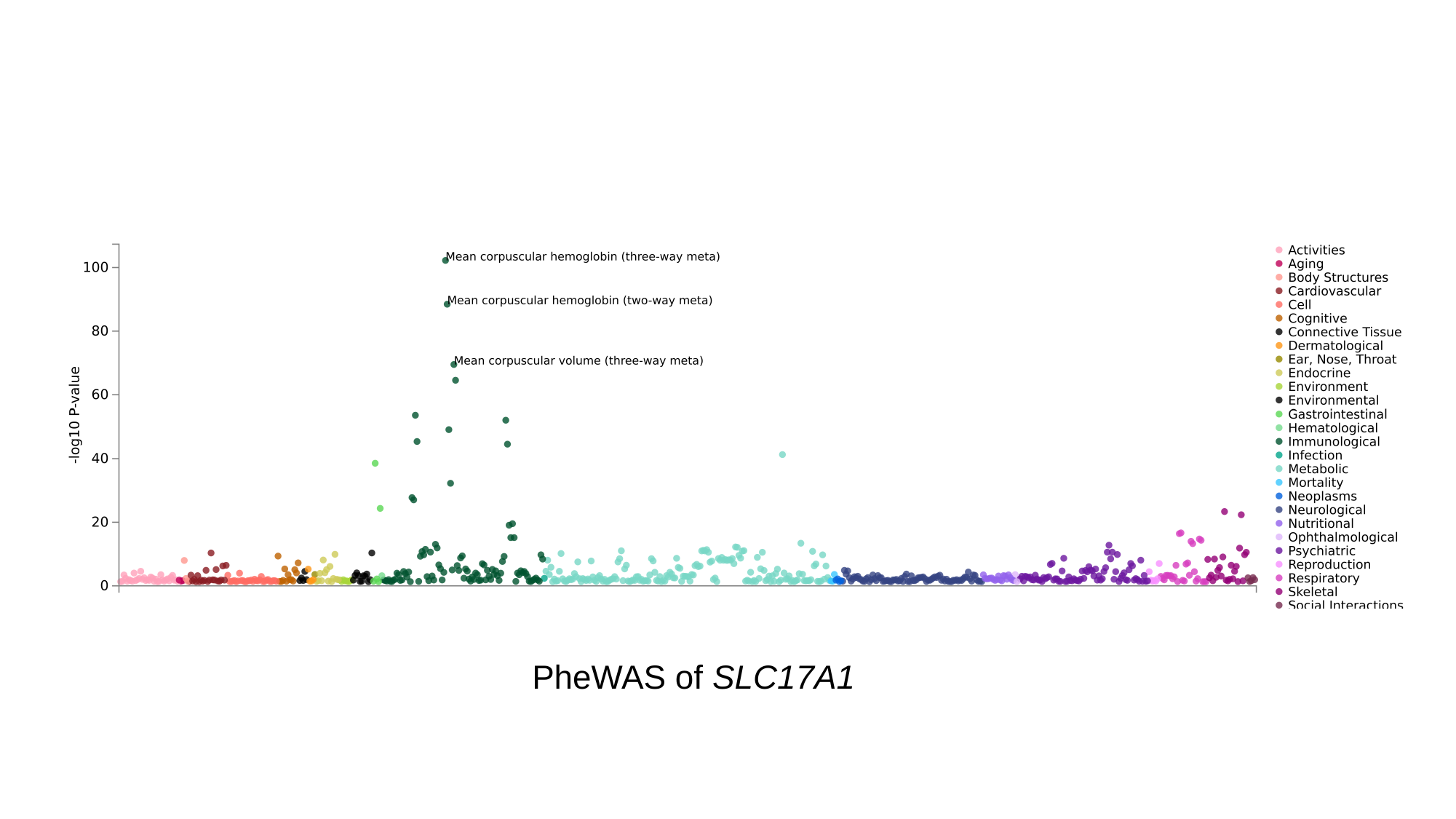

PheWAS of SLC17A1

### Slide 4
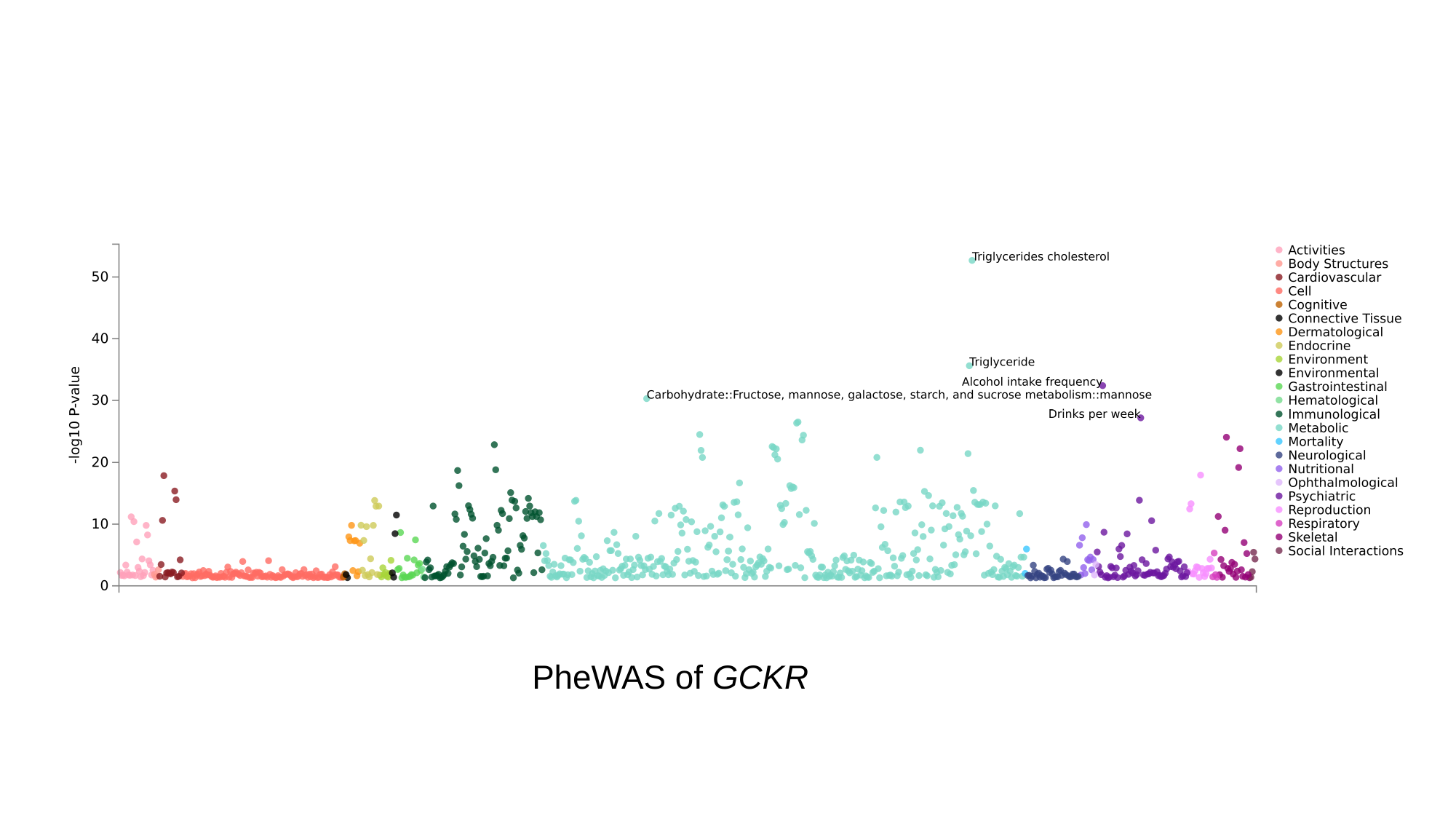

PheWAS of GCKR

### Slide 5
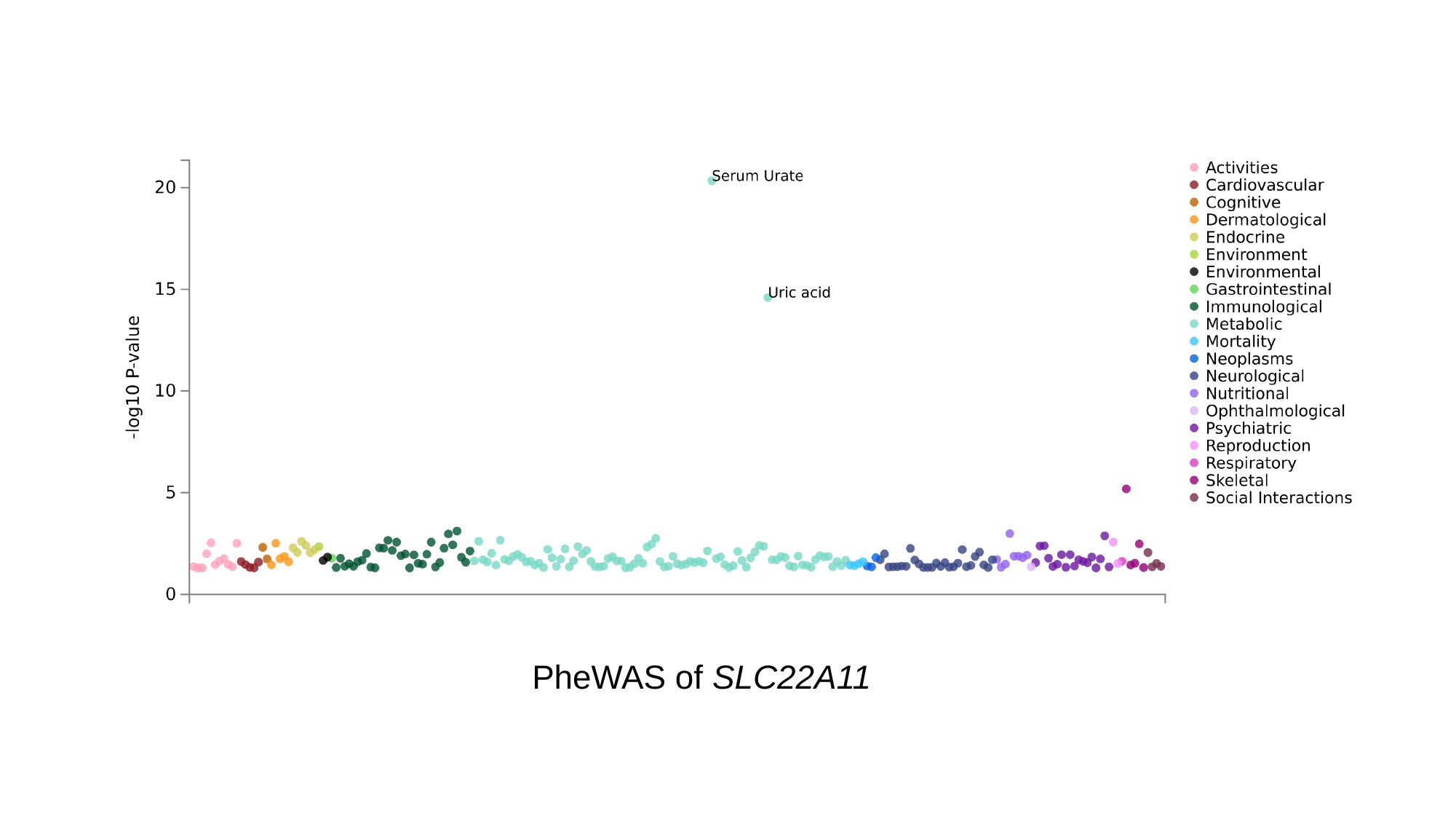

PheWAS of SLC22A11

### Slide 6
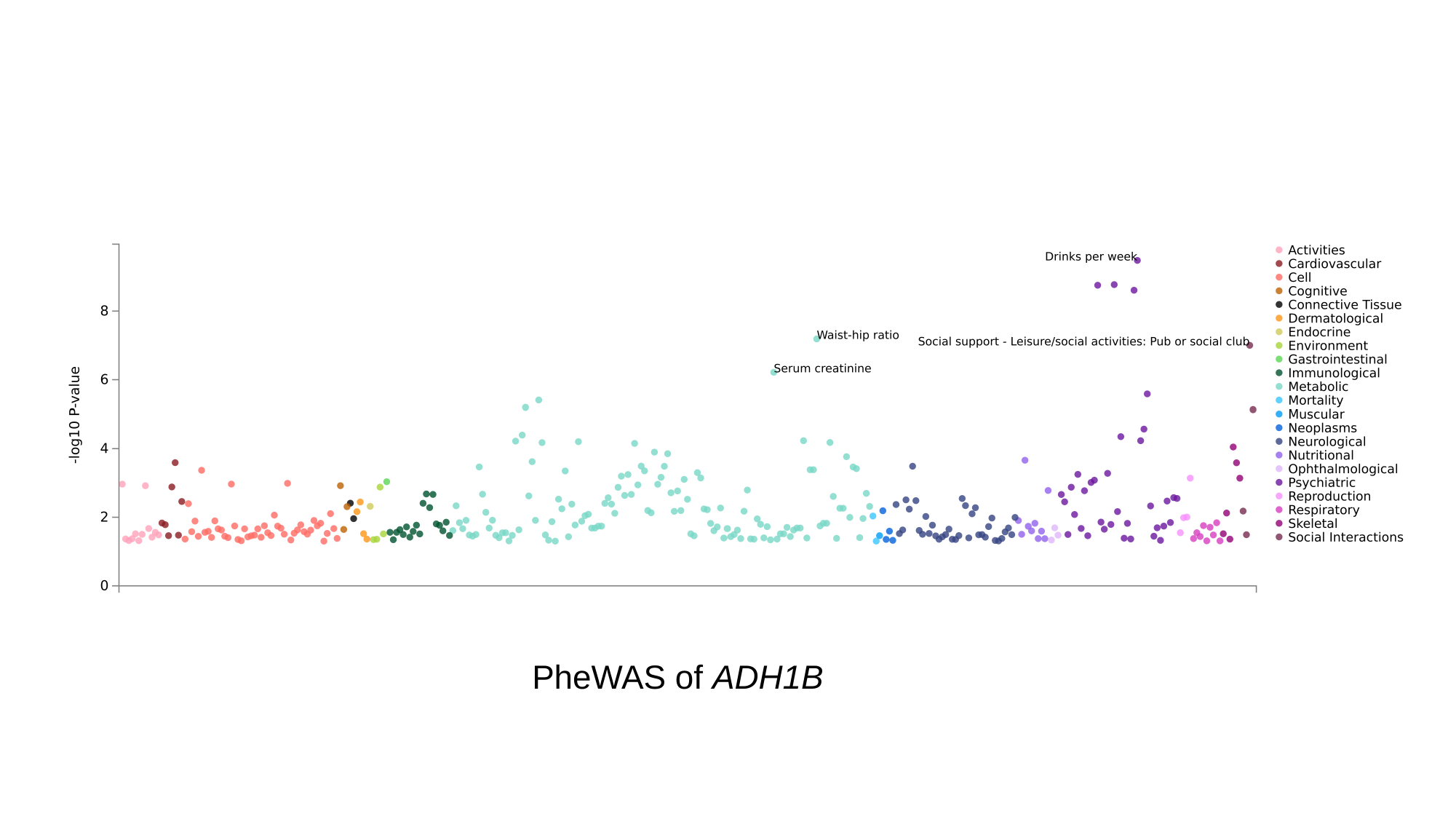

PheWAS of ADH1B

### Slide 7
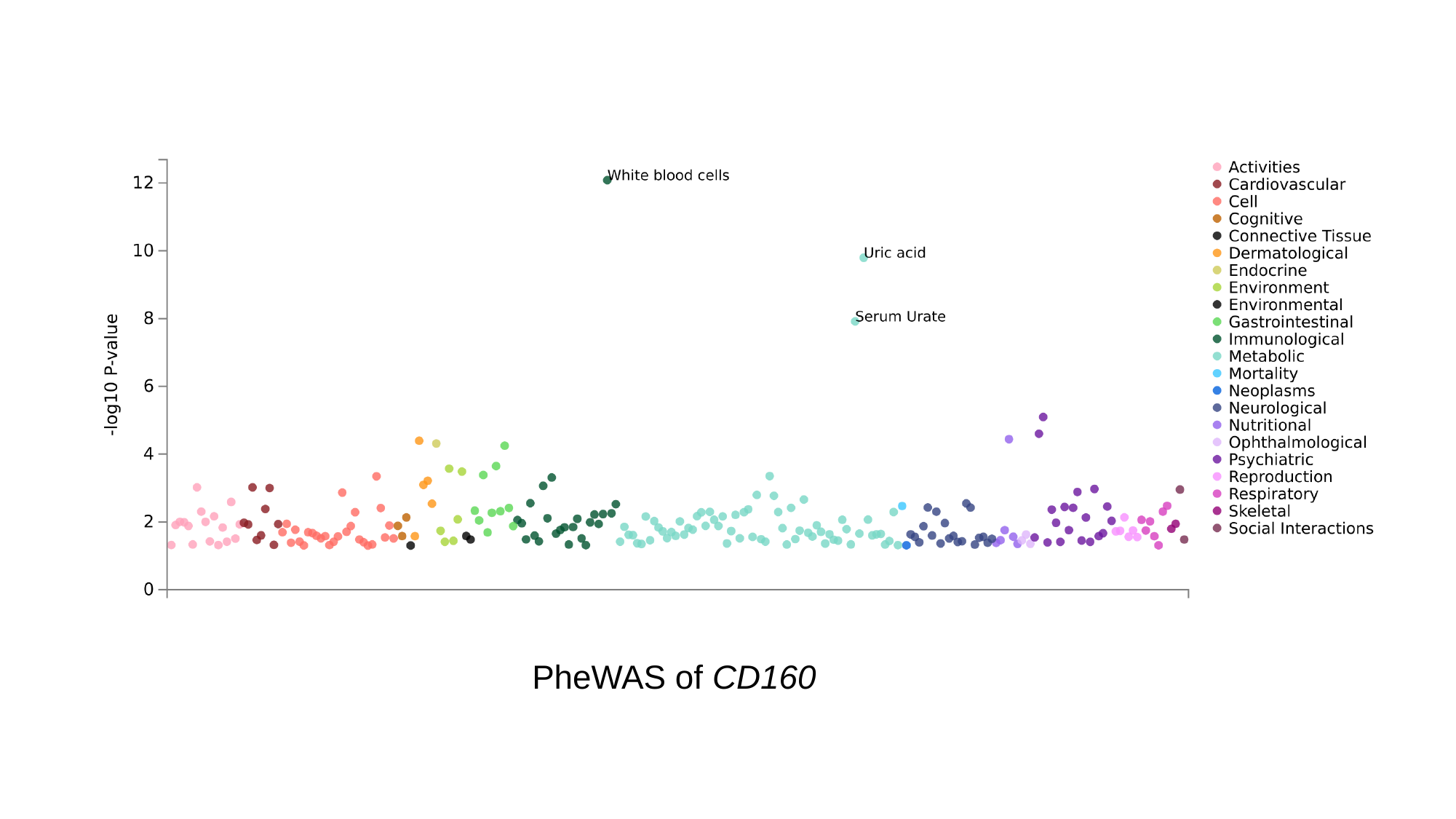

PheWAS of CD160

### Slide 8
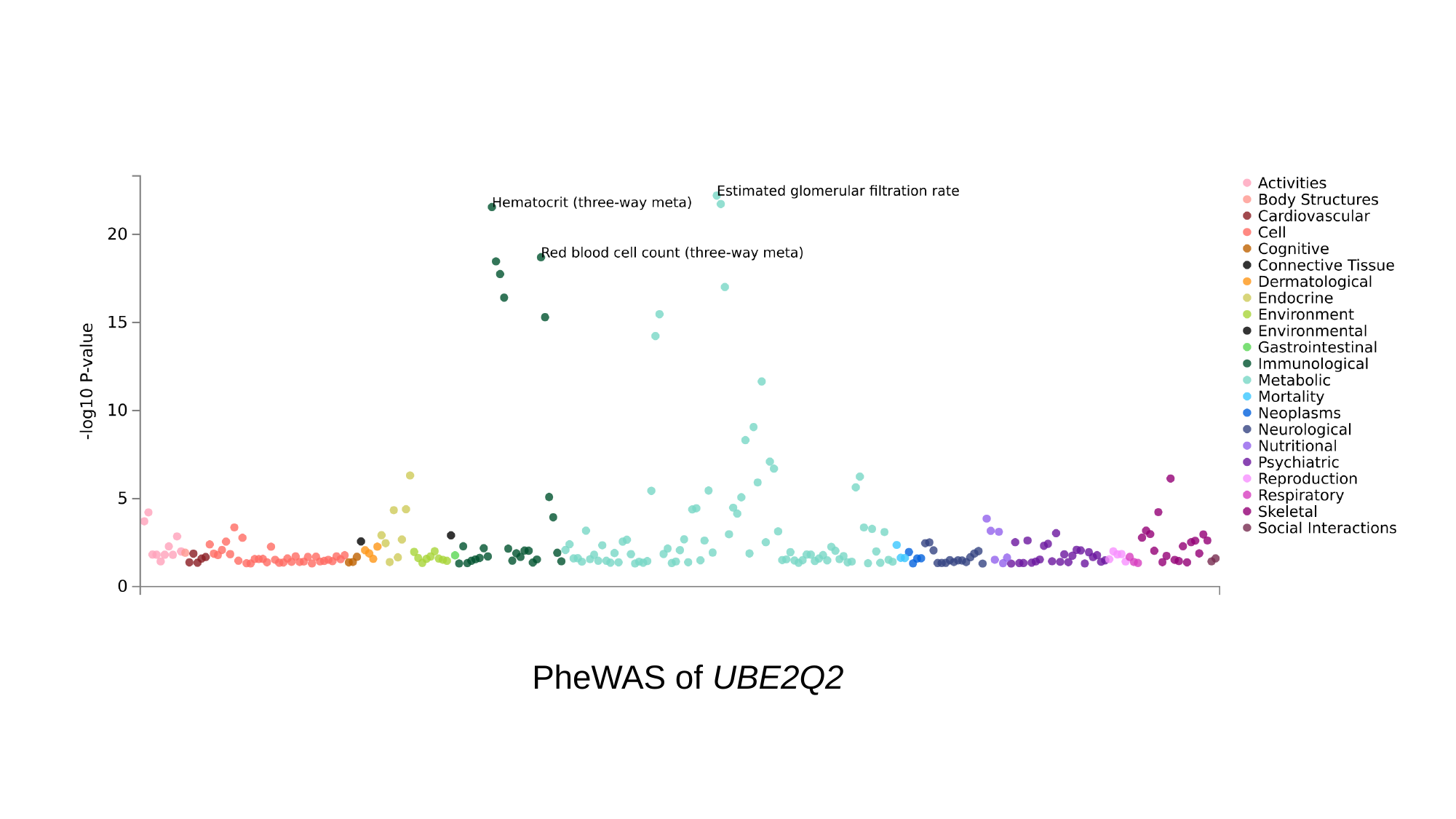

PheWAS of UBE2Q2

### Slide 9
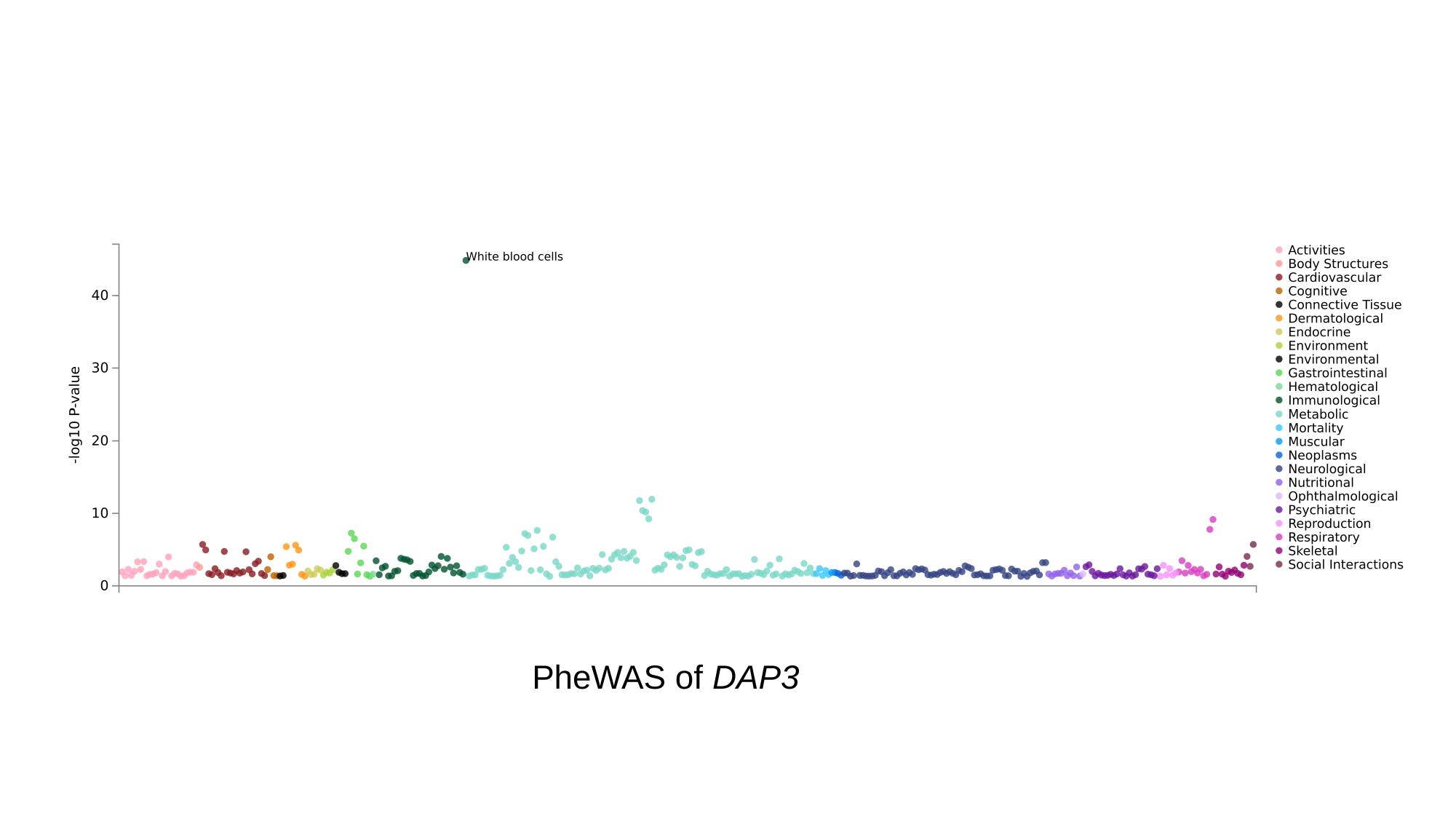

PheWAS of DAP3

### Slide 10
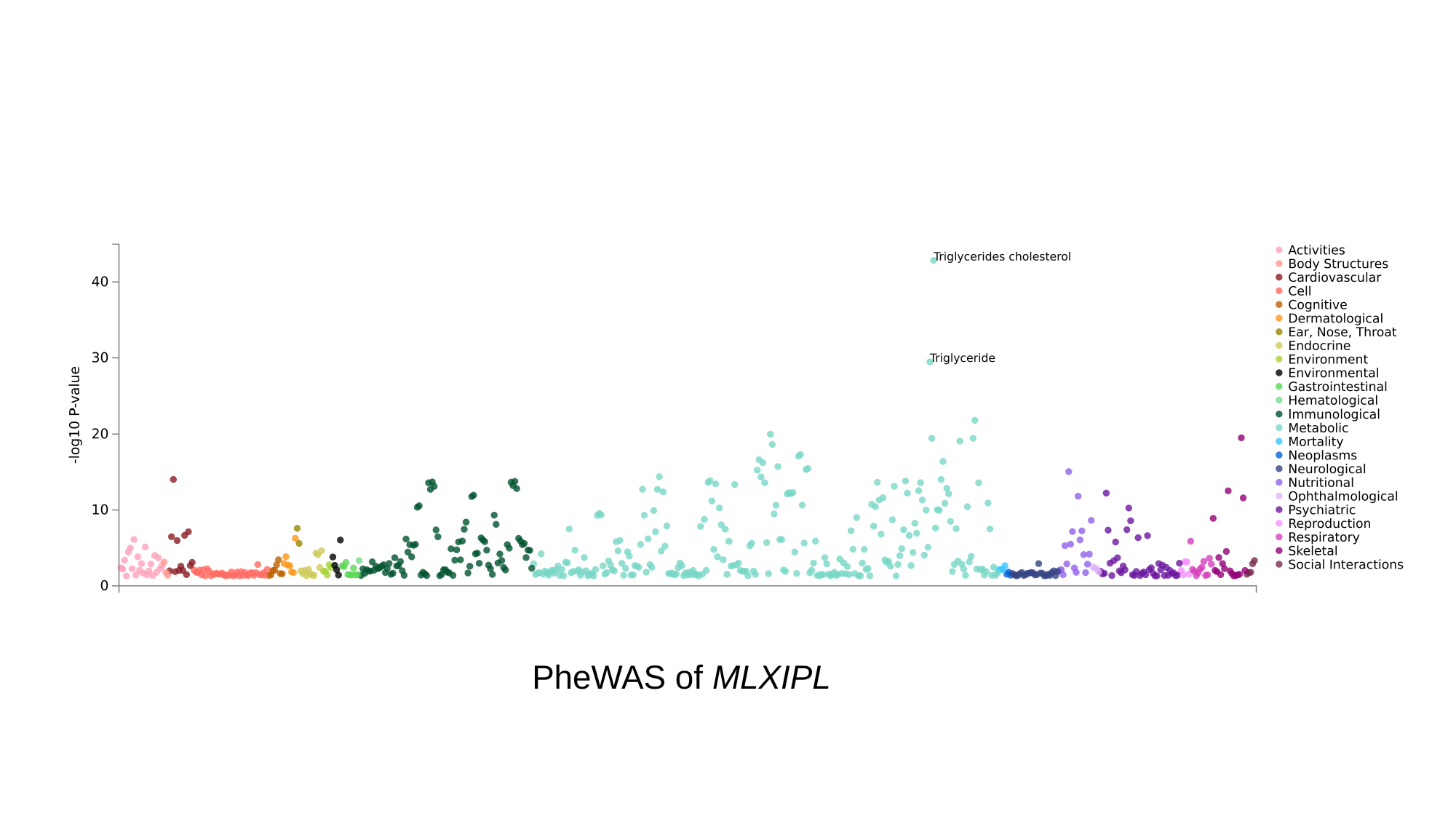

PheWAS of MLXIPL

### Slide 11
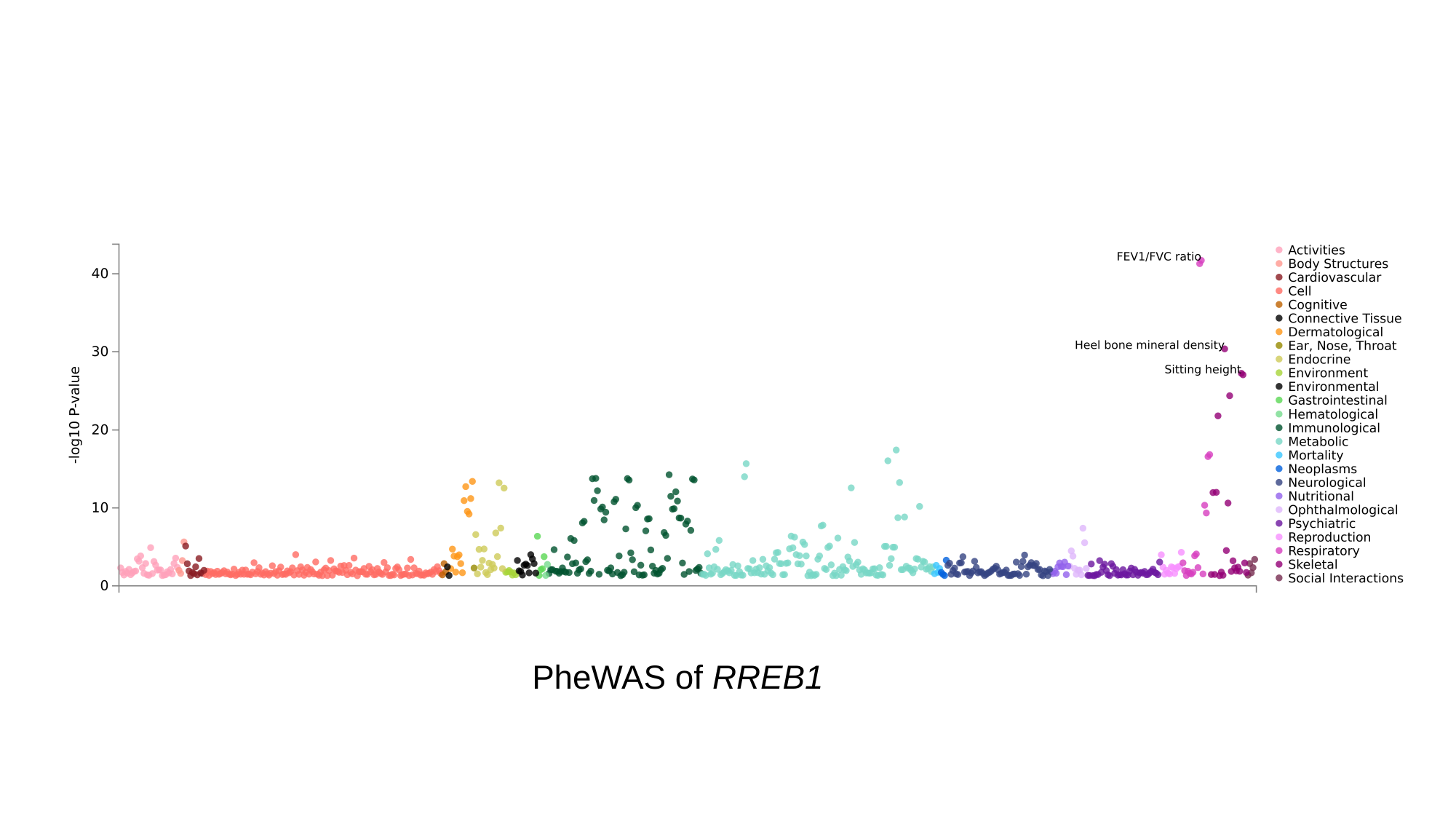

PheWAS of RREB1

### Slide 12
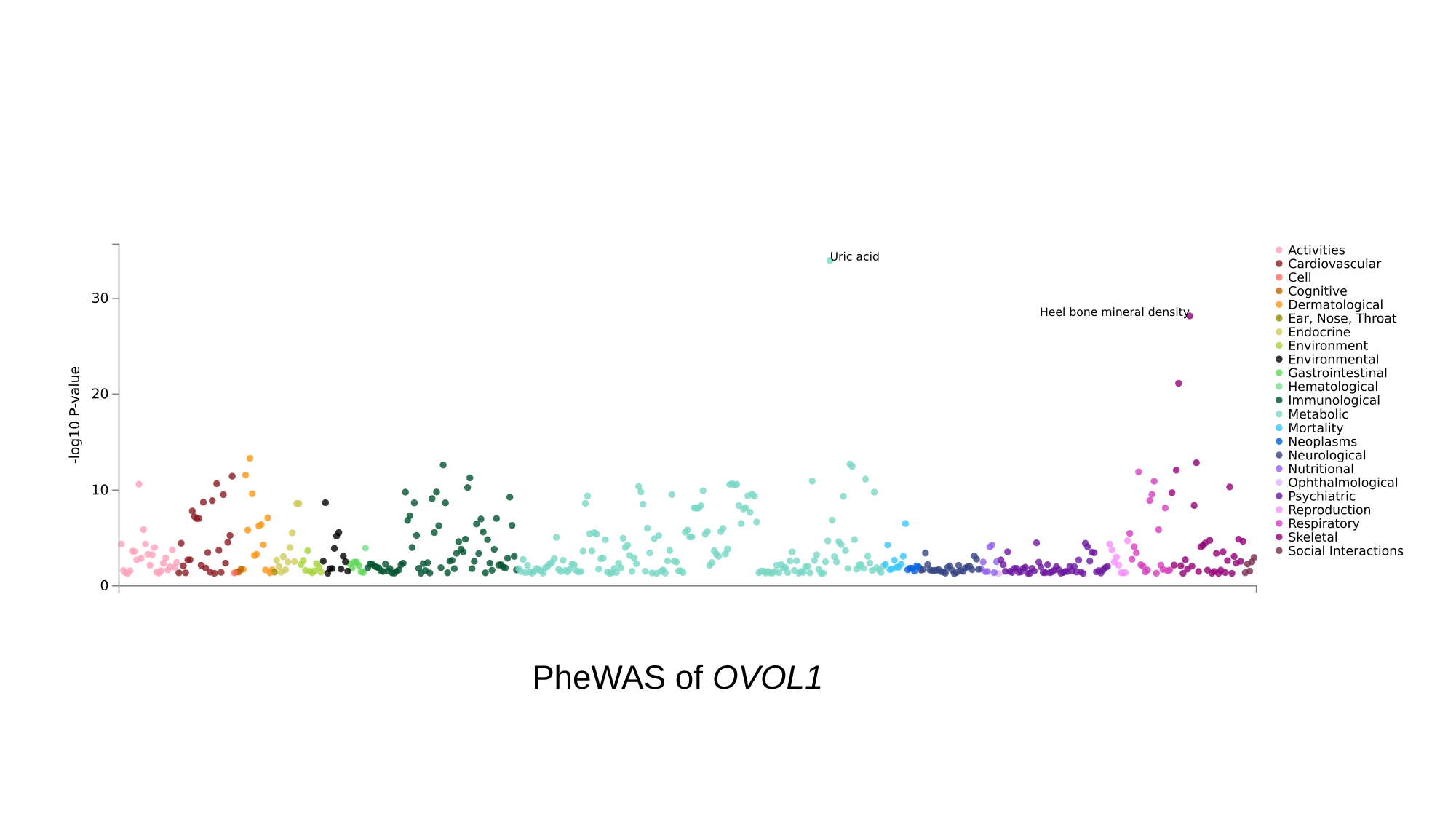

PheWAS of OVOL1

### Slide 13
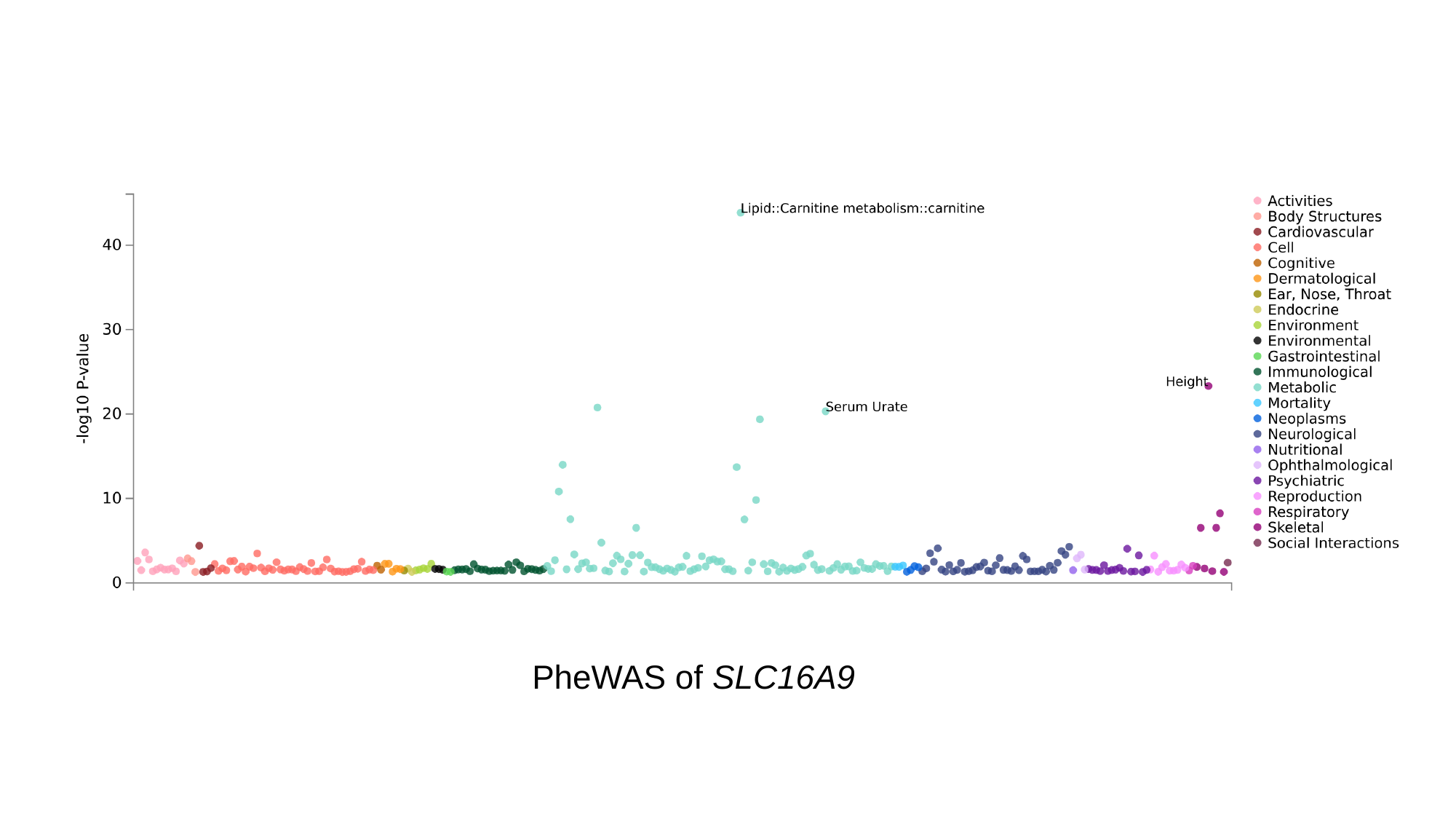

PheWAS of SLC16A9
