## Supplementary Figure 8 for "Novel Genetic Variants Associated with Gout Identified Through a Genome-Wide Study in the UK Biobank (N = 150,542)"

### Slide 1
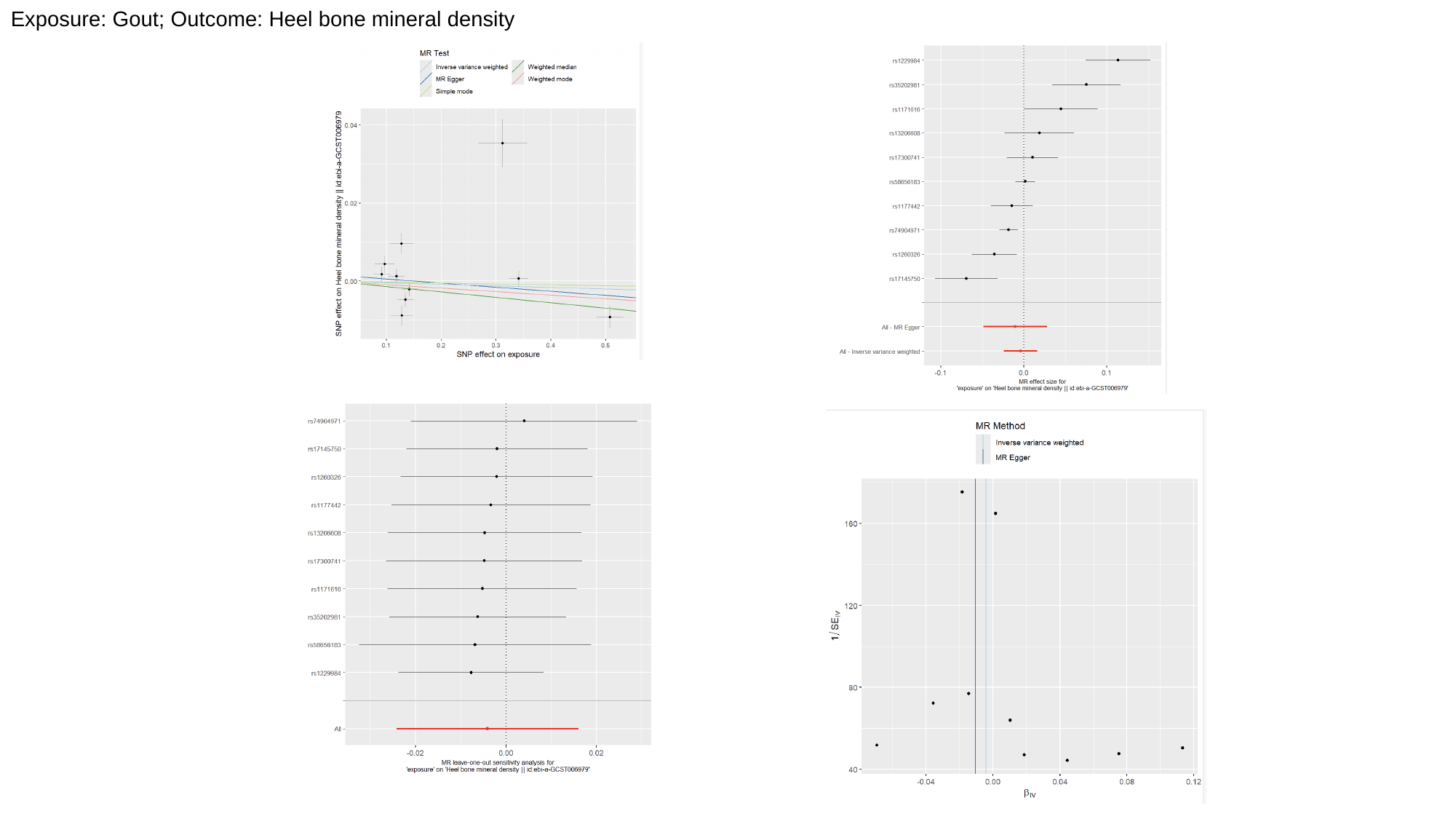

Exposure: Gout; Outcome: Heel bone mineral density

### Slide 2
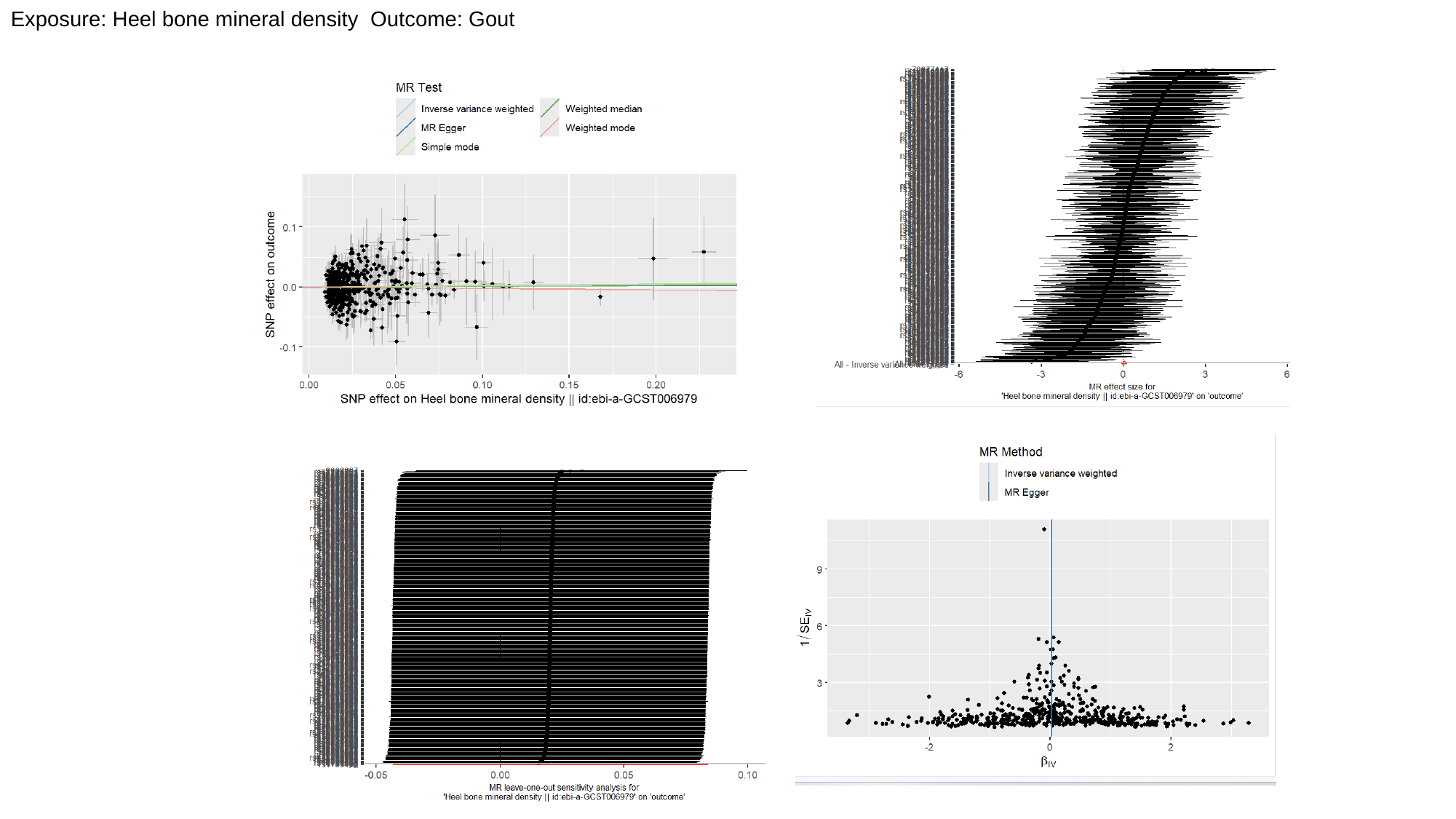

Exposure: Heel bone mineral density Outcome: Gout

### Slide 3
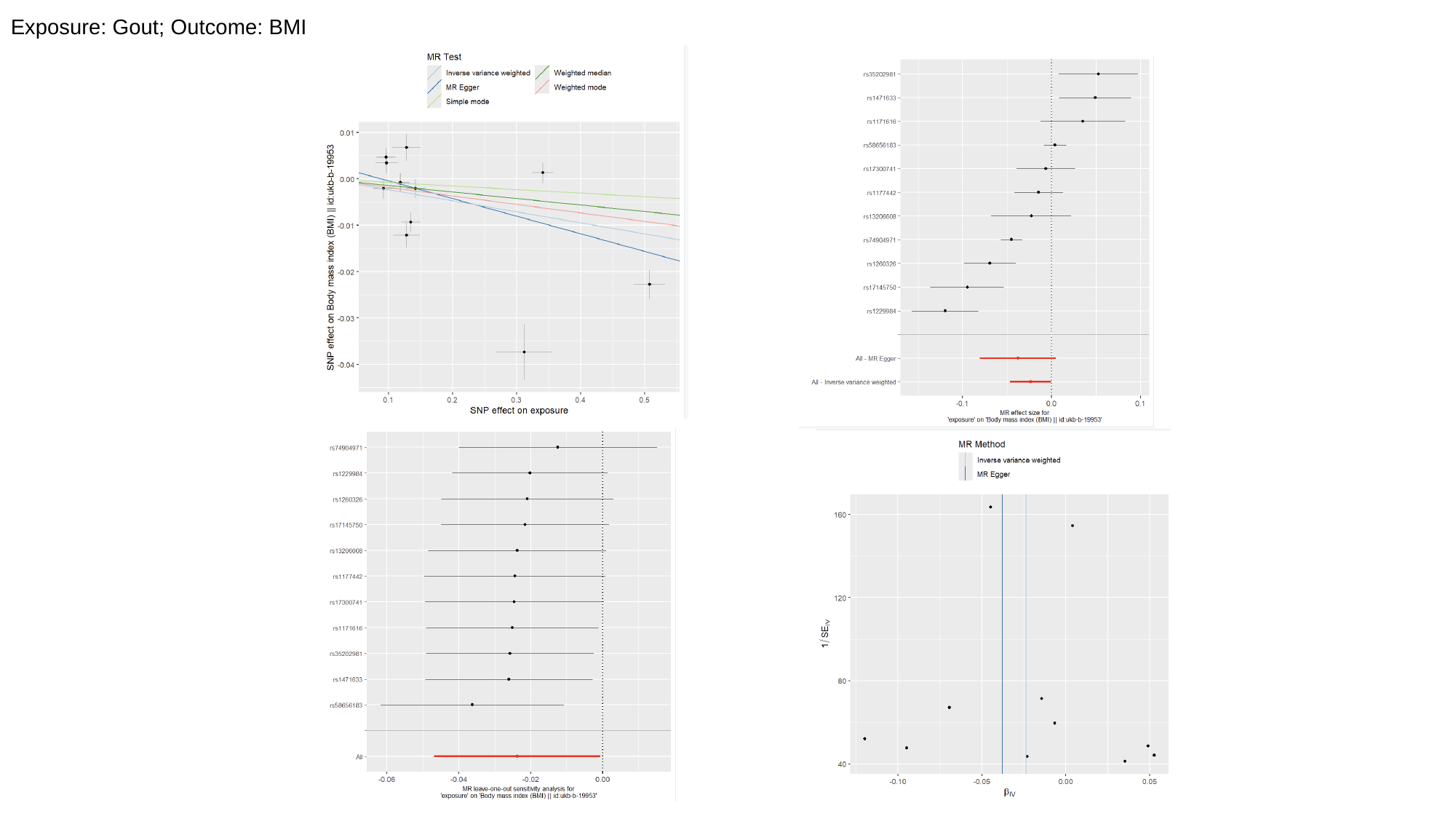

Exposure: Gout; Outcome: BMI

### Slide 4
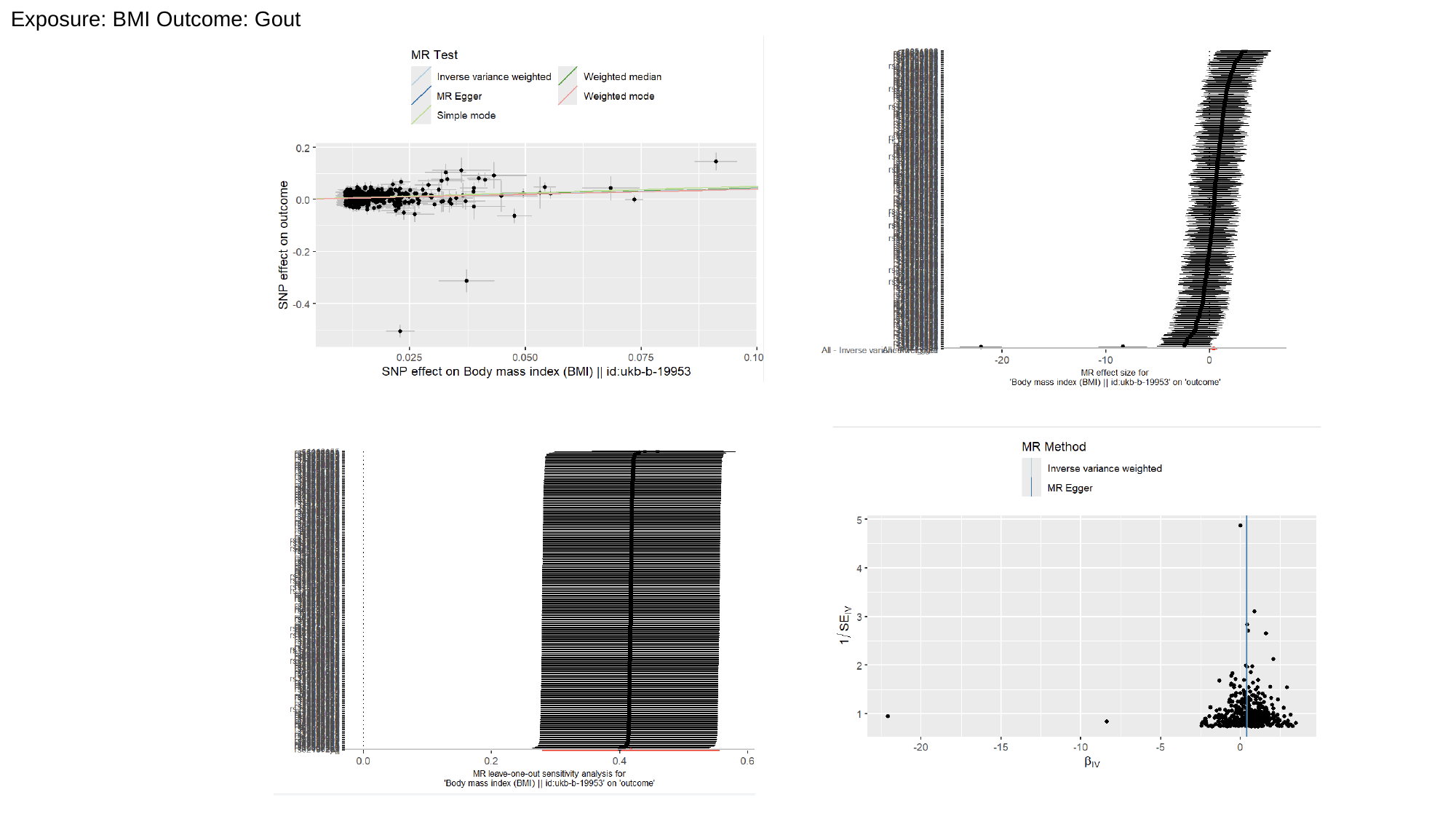

Exposure: BMI Outcome: Gout

### Slide 5
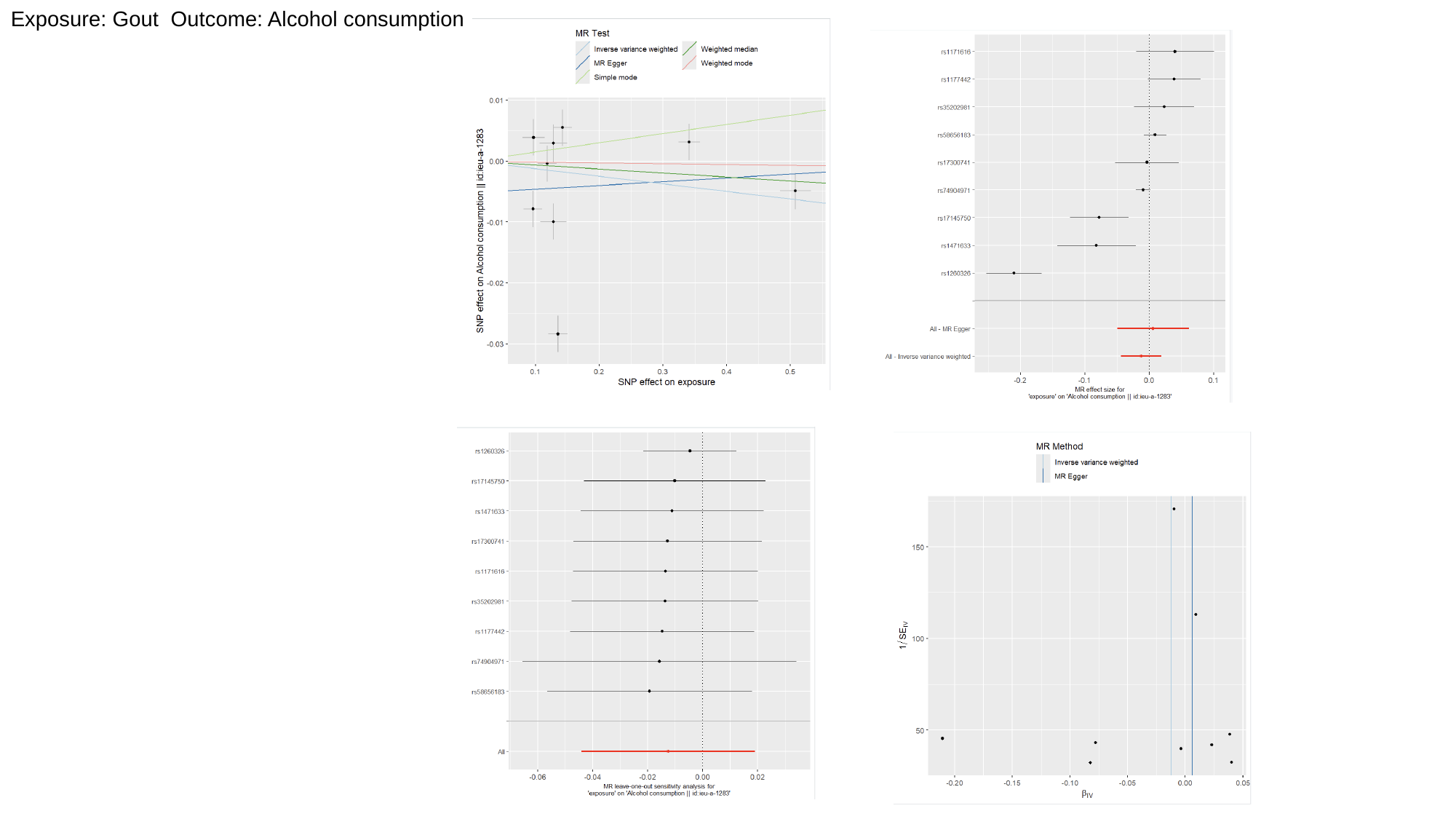

Exposure: Gout Outcome: Alcohol consumption

### Slide 6
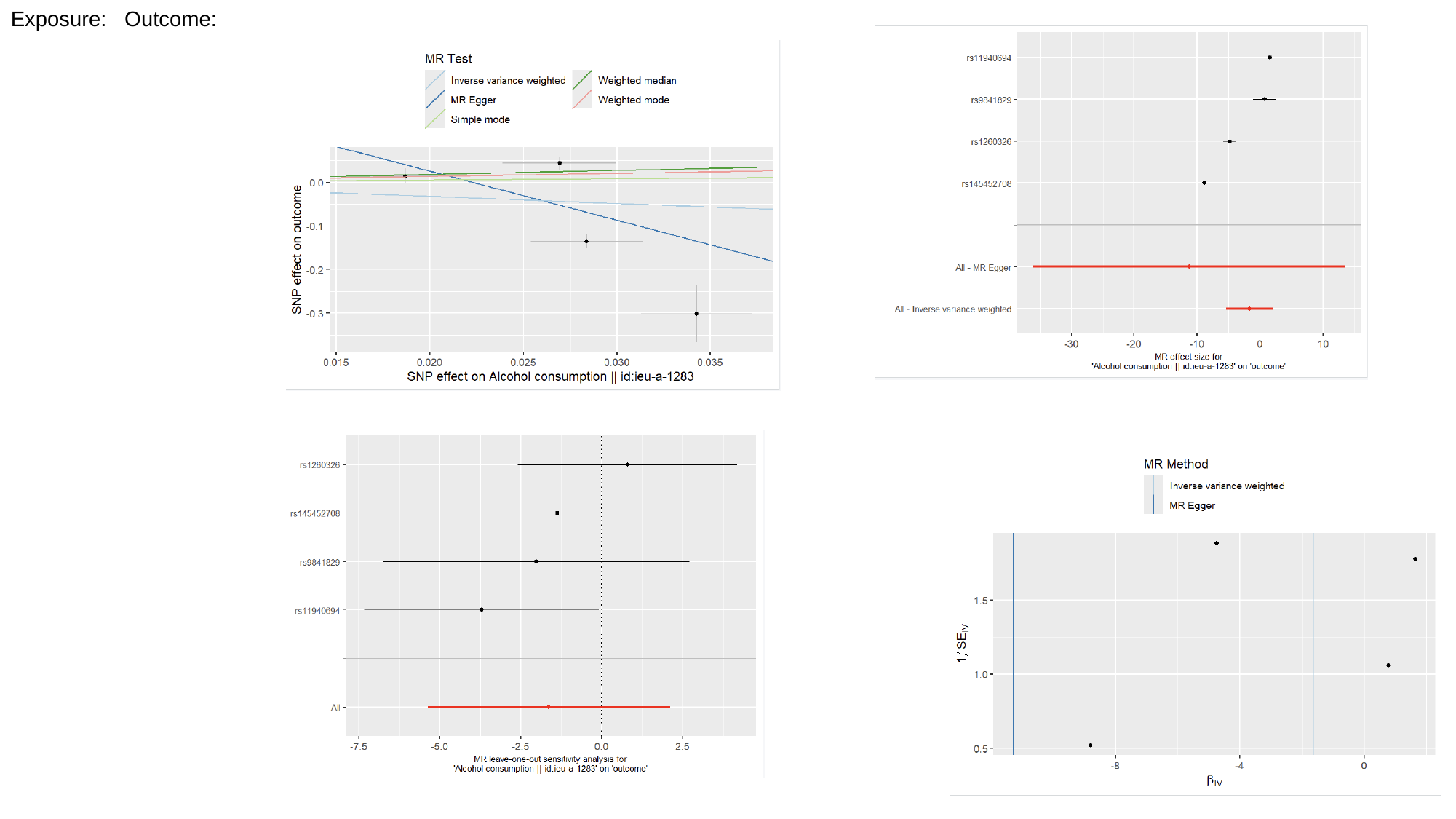

Exposure: Outcome:
