## Supplementary Files for "Novel Genetic Variants Associated with Gout Identified Through a Genome-Wide Study in the UK Biobank (N = 150,542)"

Supplementary Methods

In addition to the primary analyses, we conducted six supplementary GWAS to investigate the genetic architecture of serum urate levels across different subpopulations, stratified by gout status and sex. The first supplementary GWAS (GWAS 4) focused on 125,773 individuals without gout, aiming to uncover genetic determinants of serum urate levels in the general population free from the influence of gout-related pathophysiology. To explore sex-specific effects in this gout-free population, GWAS 5 was performed in 65,358 males, and GWAS 6 was conducted in 76,394 females. In parallel, we performed three additional GWAS analyses within gout-affected subpopulations to identify loci associated with serum urate levels in the context of gout. GWAS 7 included 9,439 individuals with gout, providing insights into the genetic architecture of serum urate levels specifically within this population. To further explore potential sex-specific differences among gout-affected individuals, GWAS 8 focused on 7,907 males with gout, while GWAS 9 analyzed 2,190 females with gout. These analyses aimed to uncover genetic variants influencing serum urate levels that may differ by sex in gout-affected cohorts. Manhattan plots illustrating the findings from these analyses are provided in the supplementary figure 1.

Supplementary Results

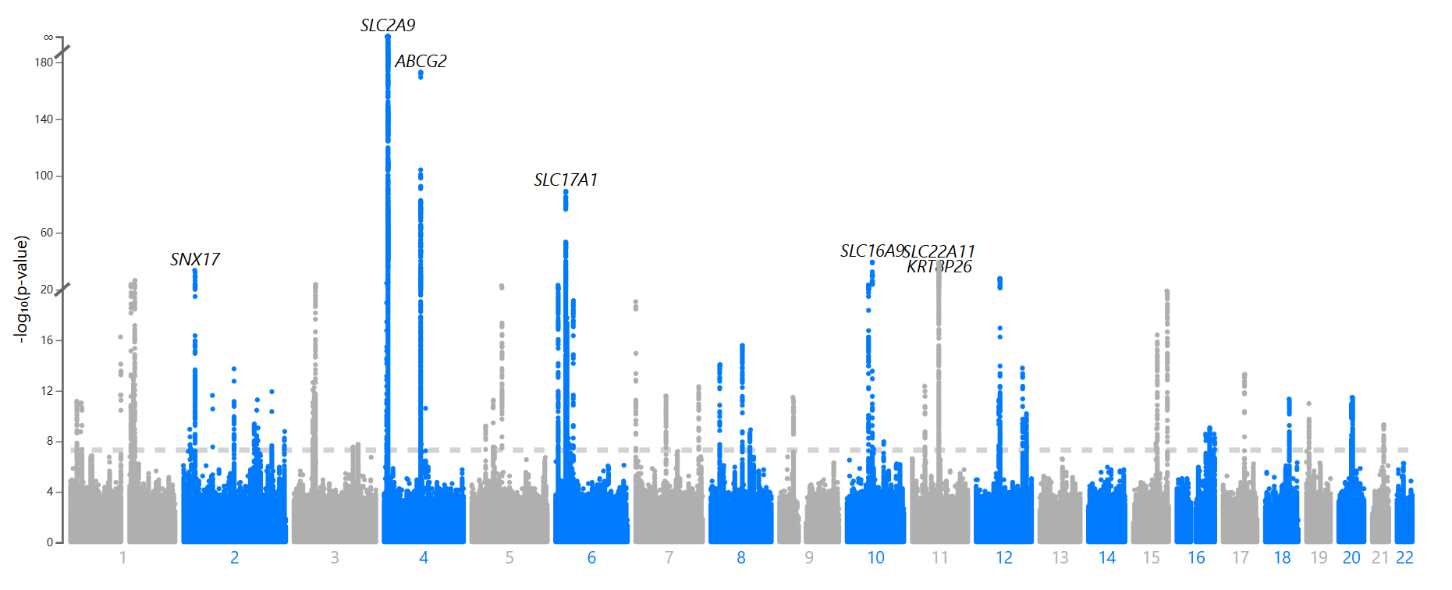
GWAS 4: Population without gout (n = 125,773)

| Locus Rank | rsID | chr | pos | *p* | | start | end | Nearest Gene |
| --- | --- | --- | --- | --- | --- | --- | --- | --- |
| 1 | rs141471965 | 4 | 89046202 | 1.00 x10^-173^ | | 87934226 | 90053135 | *ABCG2* |
| 2 | 6:25795971_ACACACC_A | 6 | 25795971 | 7.13 x10^-90^ | | 25343155 | 29607101 | *SLC17A1* |
| 3 | rs1171614 | 10 | 61469538 | 5.29 x10^-40^ | | 61388056 | 61524432 | *SLC16A9* |
| 4 | rs7929308 | 11 | 64329106 | 1.23 x10^-39^ | | 63869596 | 64584809 | *SLC22A11* |
| 5 | rs4665972 | 2 | 27598097 | 2.77 x10^-34^ | | 27498734 | 28206809 | *SNX17* |
| 6 | 11:65501060_TA_T | 11 | 65501060 | 2.58 x10^-29^ | | 64944069 | 65978453 | *KRT8P26* |
| 7 | rs4760254 | 12 | 57766392 | | 1.62 x10^-28^ | 57543572 | 57890864 | *R3HDM2* |
| 8 | rs2990223 | 1 | 155184975 | | 3.83 x10^-27^ | 155023634 | 155722506 | *GBAP1* |
| 9 | 1:145724599_CAT_C | 1 | 145724599 | | 2.44 x10^-24^ | 144894911 | 145831160 | *PDZK1* |
| 10 | 3:53038275_TC_T | 3 | 53038275 | | 2.88 x10^-24^ | 52310442 | 53174638 | *SFMBT1:RP11-894J14.5* |
| 11 | rs10994856 | 10 | 52645248 | | 8.52 x10^-24^ | 52467427 | 52690184 | *A1CF* |
| 12 | rs675209 | 6 | 7102084 | | 9.23 x10^-24^ | 6990674 | 7253262 | *RREB1* |
| 13 | rs17632159 | 5 | 72431482 | | 1.91 x10^-23^ | 72425077 | 72547189 | *RP11-232L2.2* |
| 14 | rs11634241 | 15 | 99259016 | | 1.30 x10^-20^ | 99248018 | 99320851 | *IGF1R* |
| 15 | rs9472137 | 6 | 43810469 | | 7.43 x10^-20^ | 43802563 | 43829941 | *RP11-344J7.2* |
| 16 | rs62435145 | 7 | 1286567 | | 9.16 x10^-20^ | 1269592 | 1293010 | *UNCX* |
| 17 | rs10851885 | 15 | 76304503 | | 3.95 x10^-17^ | 76064409 | 76409406 | *NRG4* |
| 18 | rs186147970 | 1 | 120484718 | | 5.64 x10^-17^ | 120484718 | 121436618 | *NOTCH2* |
| 19 | rs2943539 | 8 | 76479839 | | 2.63 x10^-16^ | 76442294 | 76774365 | *HNF4G* |
| 20 | rs11778129 | 8 | 23732226 | | 8.91 x10^-15^ | 23689919 | 23788395 | *STC1* |
| 21 | rs3184504 | 12 | 111884608 | | 1.63 x10^-14^ | 111685866 | 112986747 | *SH2B3* |
| 22 | rs17050272 | 2 | 121306440 | | 1.86 x10^-14^ | 121305771 | 121353006 | *AC073257.2* |
| 23 | 17:53367451_TG_T | 17 | 53367451 | | 5.08 x10^-14^ | 53356126 | 53381796 | *HLF* |
| 24 | rs11383974 | 1 | 150589374 | | 9.35 x10^-14^ | 150534368 | 151091290 | *ENSA* |
| 25 | rs62260779 | 3 | 48009467 | | 2.19 x10^-13^ | 46736187 | 52060065 | *MAP4* |
| 26 | rs55733296 | 11 | 30754837 | | 4.44 x10^-13^ | 30749090 | 30777790 | *DCDC1* |
| 27 | rs6464165 | 7 | 151413124 | | 4.84 x10^-13^ | 151396971 | 151415536 | *PRKAG2* |
| 28 | rs1047891 | 2 | 211540507 | | 1.20 x10^-12^ | 211540507 | 211652980 | *CPS1* |
| 29 | rs11693363 | 2 | 69821008 | | 2.38 x10^-12^ | 69746296 | 69941287 | *AAK1* |
| 30 | rs62466318 | 7 | 73042085 | | 2.61 x10^-12^ | 72809618 | 73058017 | *MLXIPL* |
| 31 | rs1969977 | 9 | 33120203 | | 3.33 x10^-12^ | 33118387 | 33180813 | *B4GALT1* |
| 32 | rs71194631 | 20 | 33257996 | | 3.36 x10^-12^ | 32514061 | 33536585 | *PIGU* |
| 33 | rs523288 | 18 | 57848369 | | 4.56 x10^-12^ | 57732418 | 57912226 | *RP11-795H16.2* |
| 34 | rs187355703 | 2 | 176993583 | | 5.27 x10^-12^ | 176765594 | 177073191 | *HOXD-AS2* |
| 35 | rs7298123 | 12 | 56860073 | | 5.60 x10^-12^ | 56860020 | 56958673 | *MIP* |
| 36 | rs255754 | 5 | 53320109 | | 5.84 x10^-12^ | 53271420 | 53327571 | *ARL15* |
| 37 | rs4233533 | 1 | 15829187 | | 7.13 x10^-12^ | 15801178 | 16039340 | *CASP9* |
| 38 | rs79598313 | 1 | 27284913 | | 9.23 x10^-12^ | 26933591 | 27368126 | *C1orf172* |
| 39 | rs112630404 | 19 | 7218635 | | 1.04 x10^-11^ | 7185576 | 7223961 | *INSR* |
| 40 | rs1229984 | 4 | 100239319 | | 2.48 x10^-11^ | 100239319 | 100239319 | *ADH1B* |
| 41 | rs11065358 | 12 | 121384495 | | 6.59 x10^-11^ | 121220375 | 121455873 | *HNF1A-AS1* |
| 42 | rs2075251 | 2 | 170011458 | | 4.40 x10^-10^ | 170003638 | 170047220 | *LRP2* |
| 43 | rs219787 | 21 | 37829508 | | 4.80 x10^-10^ | 37810961 | 37836477 | *AP000695.6:AP000695.4* |
| 44 | rs465697 | 5 | 34656290 | | 6.27 x10^-10^ | 34654441 | 34669237 | *RAI14* |
| 45 | rs11649114 | 16 | 79924857 | | 9.00 x10^-10^ | 79696939 | 79942776 | *RP11-345M22.3* |
| 46 | rs62005806 | 15 | 74883323 | | 1.04 x10^-9^ | 74629723 | 75027880 | *ARID3B* |
| 47 | rs555398472 | 12 | 58293905 | | 1.09 x10^-9^ | 58219173 | 58348609 | *RP11-620J15.2* |
| 48 | 2:15733023_TA_T | 2 | 15733023 | | 1.13 x10^-9^ | 15733023 | 15793014 | *DDX1* |
| 49 | rs201460177 | 8 | 95971303 | | 1.25 x10^-9^ | 95831273 | 96004024 | *NDUFAF6* |
| 50 | rs59104589 | 2 | 242237902 | | 1.69 x10^-9^ | 242237902 | 242403940 | *HDLBP* |
| 51 | rs74606487 | 16 | 89795305 | | 2.67 x10^-9^ | 89235401 | 90110798 | *ZNF276* |
| 52 | rs12149228 | 16 | 69765524 | | 2.69 x10^-9^ | 69548788 | 69818983 | *CTD-2033A16.3* |
| 53 | rs11202328 | 10 | 88845190 | | 1.07 x10^-8^ | 88845190 | 88966575 | *GLUD1* |
| 54 | rs1727923 | 3 | 153786303 | | 1.82 x10^-8^ | 153563827 | 154088411 | *ARHGEF26-AS1* |
| 55 | rs1744654 | 6 | 36396903 | | 2.00 x10^-8^ | 36391605 | 36524911 | *PXT1* |
| 56 | rs7299842 | 12 | 122492313 | | 3.02 x10^-8^ | 122483836 | 122630285 | *BCL7A* |
| 57 | rs40270 | 5 | 55804552 | | 3.02 x10^-8^ | 55794632 | 55816081 | *AC022431.2* |
| 58 | 3:141748306_GT_G | 3 | 141748306 | | 3.12 x10^-8^ | 141658316 | 141825598 | *TFDP2* |
| 59 | rs12618898 | 2 | 171890306 | | 3.43 x10^-8^ | 171762882 | 172039547 | *TLK1* |
| 60 | rs12249208 | 10 | 63730012 | | 4.14 x10^-8^ | 63697657 | 63756491 | *ARID5B* |
| 61 | rs71407906 | 2 | 18678677 | | 4.37 x10^-8^ | 18672346 | 18691613 | *RP11-111J6.2* |

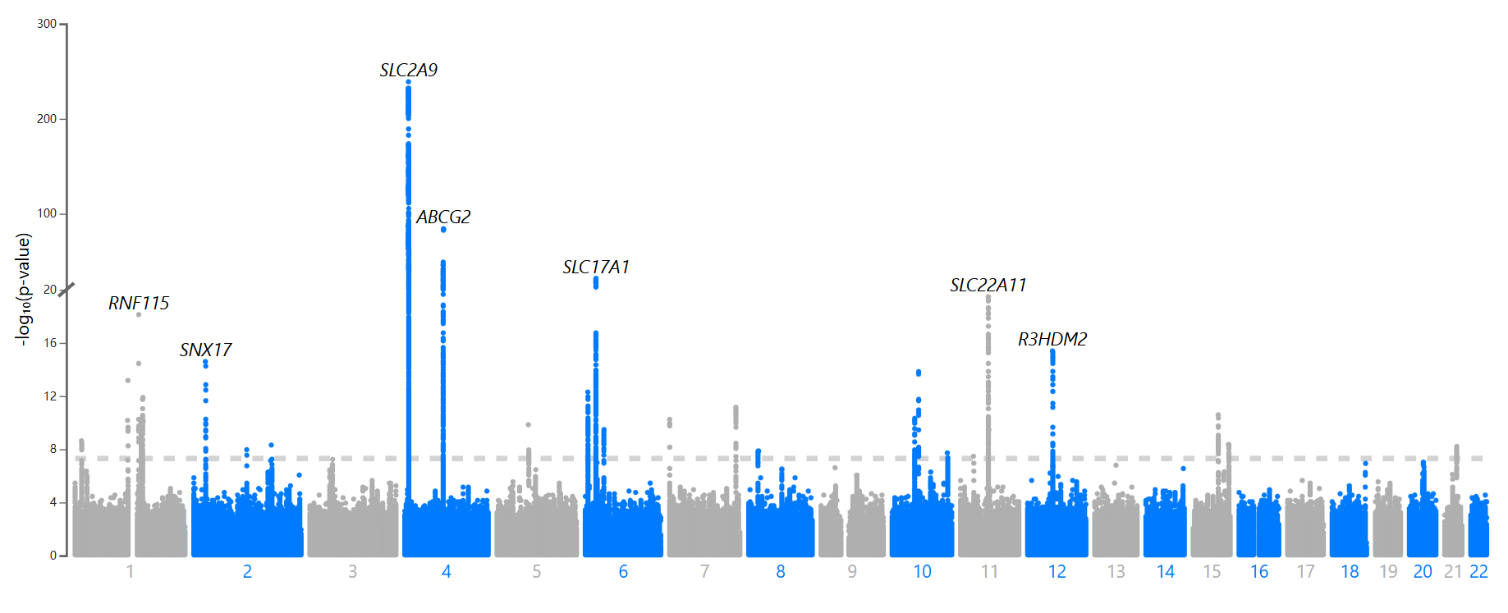
GWAS 5: Male cohorts without gout (n = 65,358)

| Locus Rank | rsID | chr | pos | *p* | start | end | Nearest Gene |
| --- | --- | --- | --- | --- | --- | --- | --- |
| 1 | rs16890979 | 4 | 9922167 | 1.11 x10^-239^ | 9035604 | 10916797 | *SLC2A9* |
| 2 | rs141471965 | 4 | 89046202 | 9.00 x10^-85^ | 88591554 | 89278779 | *ABCG2* |
| 3 | rs6909187 | 6 | 25785925 | 1.40 x10^-32^ | 25486626 | 26140100 | *SLC17A1* |
| 4 | rs7929308 | 11 | 64329106 | 3.37 x10^-20^ | 64248152 | 64584809 | *SLC22A11* |
| 5 | rs75246752 | 1 | 145630111 | 7.53 x10^-19^ | 144923070 | 145814880 | *RNF115* |
| 6 | rs4760254 | 12 | 57766392 | 4.05 x10^-16^ | 57634698 | 57851893 | *R3HDM2* |
| 7 | rs4665972 | 2 | 27598097 | 2.56 x10^-15^ | 27598097 | 27752871 | *SNX17* |
| 8 | rs1171616 | 10 | 61468589 | 1.47 x10^-14^ | 61397399 | 61469538 | *SLC16A9* |
| 9 | rs190675279 | 1 | 121152273 | 6.72 x10^-14^ | 120484718 | 121436618 | *AL592494.5* |
| 10 | 6:7120908_AT_A | 6 | 7120908 | 5.02 x10^-13^ | 7043760 | 7220379 | *RREB1* |
| 11 | rs2990223 | 1 | 155184975 | 1.29 x10^-12^ | 155095750 | 155722506 | *GBAP1* |
| 12 | rs17173238 | 7 | 151406220 | 6.98 x10^-12^ | 151396971 | 151415536 | *PRKAG2* |
| 13 | rs11636220 | 15 | 76239068 | 2.56 x10^-11^ | 76136194 | 76347375 | *NRG4* |
| 14 | rs11375604 | 10 | 52646867 | 4.77 x10^-11^ | 52478264 | 52648454 | *A1CF* |
| 15 | rs11761603 | 7 | 1286912 | 5.70 x10^-11^ | 1280952 | 1286912 | *UNCX* |
| 16 | rs750652170 | 1 | 150678001 | 8.48 x10^-11^ | 150536180 | 150958977 | *HORMAD1* |
| 17 | rs17632159 | 5 | 72431482 | 1.44 x10^-10^ | 72425077 | 72547189 | *RP11-232L2.2* |
| 18 | rs2396084 | 6 | 43804825 | 3.30 x10^-10^ | 43804103 | 43821897 | *RP11-344J7.2* |
| 19 | 11:65501060_TA_T | 11 | 65501060 | 3.46 x10^-10^ | 65401336 | 65570146 | *KRT8P26* |
| 20 | rs6679229 | 1 | 15914078 | 2.33 x10^-9^ | 15808198 | 15993603 | *DNAJC16:RP4-680D5.2* |
| 21 | rs2137683 | 15 | 99289638 | 4.50 x10^-9^ | 99249029 | 99296459 | *IGF1R* |
| 22 | rs187355703 | 2 | 176993583 | 4.87 x10^-9^ | 176984544 | 177073191 | *HOXD-AS2* |
| 23 | rs219787 | 21 | 37829508 | 6.16 x10^-9^ | 37810961 | 37836477 | *AP000695.6:AP000695.4* |
| 24 | rs17050272 | 2 | 121306440 | 1.08 x10^-8^ | 121306440 | 121310269 | *AC073257.2* |
| 25 | rs4872205 | 8 | 23785018 | 1.36 x10^-8^ | 23697492 | 23788395 | *STC1* |
| 26 | 10:126504461_CT_C | 10 | 126504461 | 1.89 x10^-8^ | 126396511 | 126551900 | *FAM175B* |

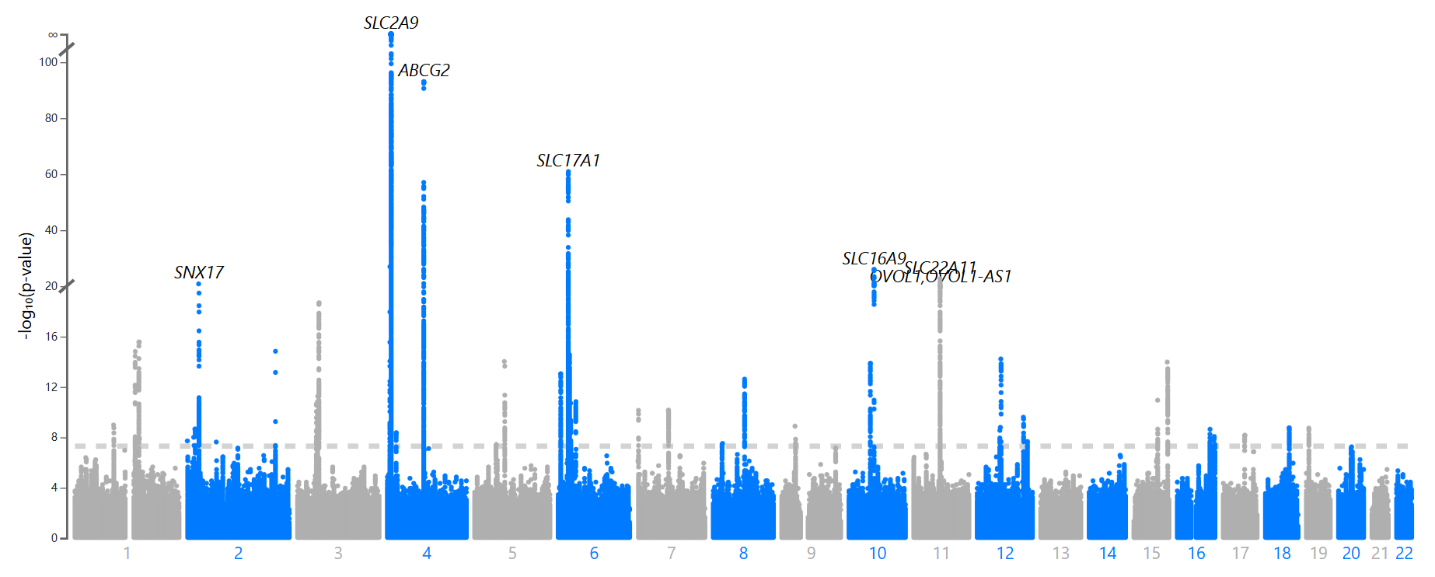
GWAS 6: Female cohorts without gout (n = 76,394)

| Locus Rank | rsID | chr | pos | *p* | start | end | Nearest Gene |
| --- | --- | --- | --- | --- | --- | --- | --- |
| 1 | rs138409370 | 4 | 89044312 | 9.98 x10^-94^ | 87934226 | 89747079 | *ABCG2* |
| 2 | 6:25795971_ACACACC_A | 6 | 25795971 | 1.15 x10^-61^ | 25343155 | 29607101 | *SLC17A1* |
| 3 | rs1171614 | 10 | 61469538 | 9.52 x10^-27^ | 61388056 | 61469538 | *SLC16A9* |
| 4 | rs2164495 | 11 | 64342474 | 2.37 x10^-23^ | 63873673 | 64584809 | *SLC22A11* |
| 5 | rs4665972 | 2 | 27598097 | 1.51 x10^-21^ | 27548038 | 27950837 | *SNX17* |
| 6 | rs5792371 | 11 | 65557116 | 1.26 x10^-20^ | 65372518 | 65580638 | *OVOL1:OVOL1-AS1* |
| 7 | 3:53038262_TTTC_T | 3 | 53038262 | 2.09 x10^-19^ | 52310442 | 53174638 | *SFMBT1:RP11-894J14.5* |
| 8 | rs2990223 | 1 | 155184975 | 2.71 x10^-16^ | 155073221 | 155722506 | *GBAP1* |
| 9 | rs1047891 | 2 | 211540507 | 1.48 x10^-15^ | 211540507 | 211652980 | *CPS1* |
| 10 | 1:145724599_CAT_C | 1 | 145724599 | 1.55 x10^-15^ | 145561116 | 145814880 | *PDZK1* |
| 11 | 12:57801037_GA_G | 12 | 57801037 | 6.00 x10^-15^ | 57645789 | 57844049 | *R3HDM2* |
| 12 | rs10942549 | 5 | 72426137 | 9.51 x10^-15^ | 72425458 | 72464994 | *TMEM171* |
| 13 | rs11634241 | 15 | 99259016 | 1.06 x10^-14^ | 99248018 | 99320851 | *IGF1R* |
| 14 | rs10994856 | 10 | 52645248 | 1.29 x10^-14^ | 52469952 | 52648454 | *A1CF* |
| 15 | rs143321297 | 6 | 7108549 | 9.21 x10^-14^ | 7023853 | 7220379 | *RREB1* |
| 16 | rs2941485 | 8 | 76479458 | 2.42 x10^-13^ | 76442294 | 76565110 | *HNF4G* |
| 17 | rs549664712 | 3 | 49749648 | 5.58 x10^-12^ | 46764220 | 52060065 | *RNF123* |
| 18 | rs374504639 | 15 | 76283012 | 1.13 x10^-11^ | 76136194 | 76347375 | *NRG4* |
| 19 | rs147954739 | 6 | 43817762 | 1.46 x10^-11^ | 43804103 | 43829941 | *RP11-344J7.2* |
| 20 | rs13232120 | 7 | 72983310 | 7.03 x10^-11^ | 72841823 | 73058017 | *TBL2* |
| 21 | rs62435145 | 7 | 1286567 | 7.30 x10^-11^ | 1280952 | 1293010 | *UNCX* |
| 22 | rs3184504 | 12 | 111884608 | 2.53 x10^-10^ | 111826477 | 112883476 | *SH2B3* |
| 23 | rs10782959 | 1 | 93811569 | 1.04 x10^-9^ | 93538949 | 93895483 | *RP4-717I23.3:DR1* |
| 24 | 9:33173589_GGT_G | 9 | 33173589 | 1.29 x10^-9^ | 33119241 | 33180362 | *RP11-326F20.5* |
| 25 | rs7298123 | 12 | 56860073 | 1.61 x10^-9^ | 56860020 | 56958673 | *MIP* |
| 26 | rs476828 | 18 | 57852587 | 1.81 x10^-9^ | 57732418 | 57912226 | *RP11-795H16.2* |
| 27 | rs74578307 | 2 | 18680691 | 2.18 x10^-9^ | 18672346 | 18691613 | *RP11-111J6.2* |
| 28 | rs8113542 | 19 | 7202759 | 2.24 x10^-9^ | 7185576 | 7213500 | *INSR* |
| 29 | rs4575545 | 16 | 79755446 | 2.27 x10^-9^ | 79696939 | 79756197 | *RP11-345M22.1:RP11-345M22.2* |
| 30 | rs420777 | 4 | 22805792 | 4.26 x10^-9^ | 22794601 | 22830939 | *GBA3* |
| 31 | rs3829578 | 17 | 53381510 | 6.89 x10^-9^ | 53364788 | 53386826 | *HLF* |
| 32 | rs74606487 | 16 | 89795305 | 8.49 x10^-9^ | 89535888 | 90110798 | *ZNF276* |
| 33 | 2:15733023_TA_T | 2 | 15733023 | 9.86 x10^-9^ | 15733023 | 15793014 | *DDX1* |
| 34 | rs7563362 | 2 | 620297 | 1.92 x10^-8^ | 600575 | 653874 | *AC093326.3* |
| 35 | rs7979473 | 12 | 121420260 | 2.15 x10^-8^ | 121380544 | 121455873 | *HNF1A* |
| 36 | rs11693363 | 2 | 69821008 | 2.36 x10^-8^ | 69813458 | 69941287 | *AAK1* |
| 37 | rs9314273 | 8 | 23735559 | 3.06 x10^-8^ | 23697492 | 23788395 | *STC1* |
| 38 | rs1664798 | 5 | 53285757 | 3.63 x10^-8^ | 53271420 | 53287099 | *ARL15* |

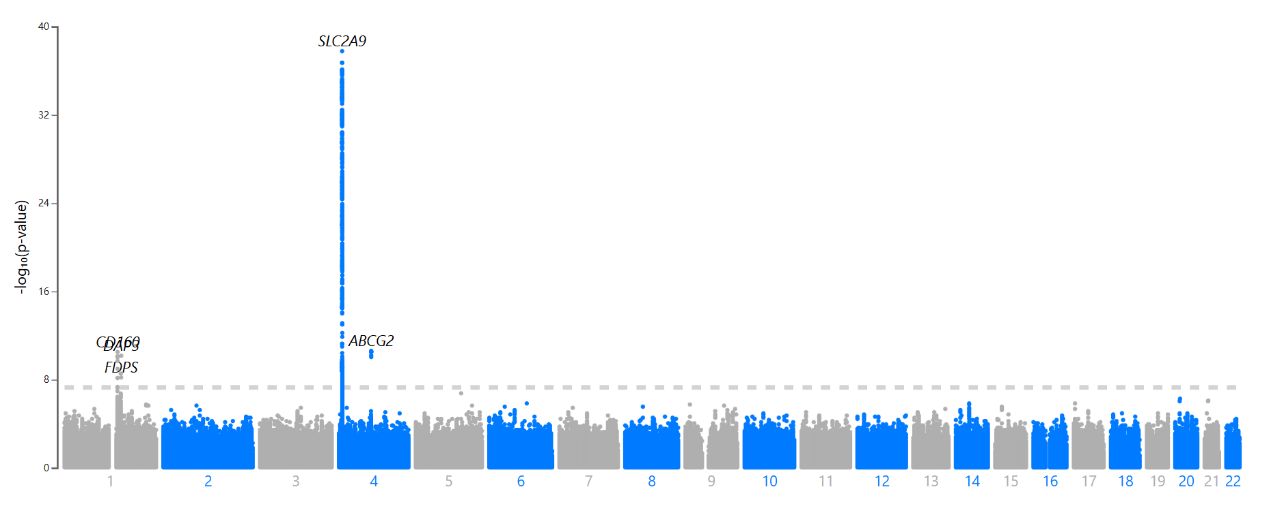
GWAS 7: Population with gout (n =9,439)

| Locus Rank | rsID | chr | pos | *p* | start | end | Nearest Gene |
| --- | --- | --- | --- | --- | --- | --- | --- |
| 1 | rs13129697 | 4 | 9926967 | 1.69 x10^-38^ | 9915325 | 10528226 | *SLC2A9* |
| 2 | rs74904971 | 4 | 89050026 | 2.43 x10^-11^ | 89039082 | 89054667 | *ABCG2* |
| 3 | rs149865899 | 1 | 145719488 | 3.10 x10^-11^ | 145719488 | 145814880 | *CD160* |
| 4 | rs3041216 | 1 | 155659698 | 6.74 x10^-11^ | 155277204 | 155659698 | *DAP3* |

GWAS 8: Male cohorts with gout (n = 7,907)

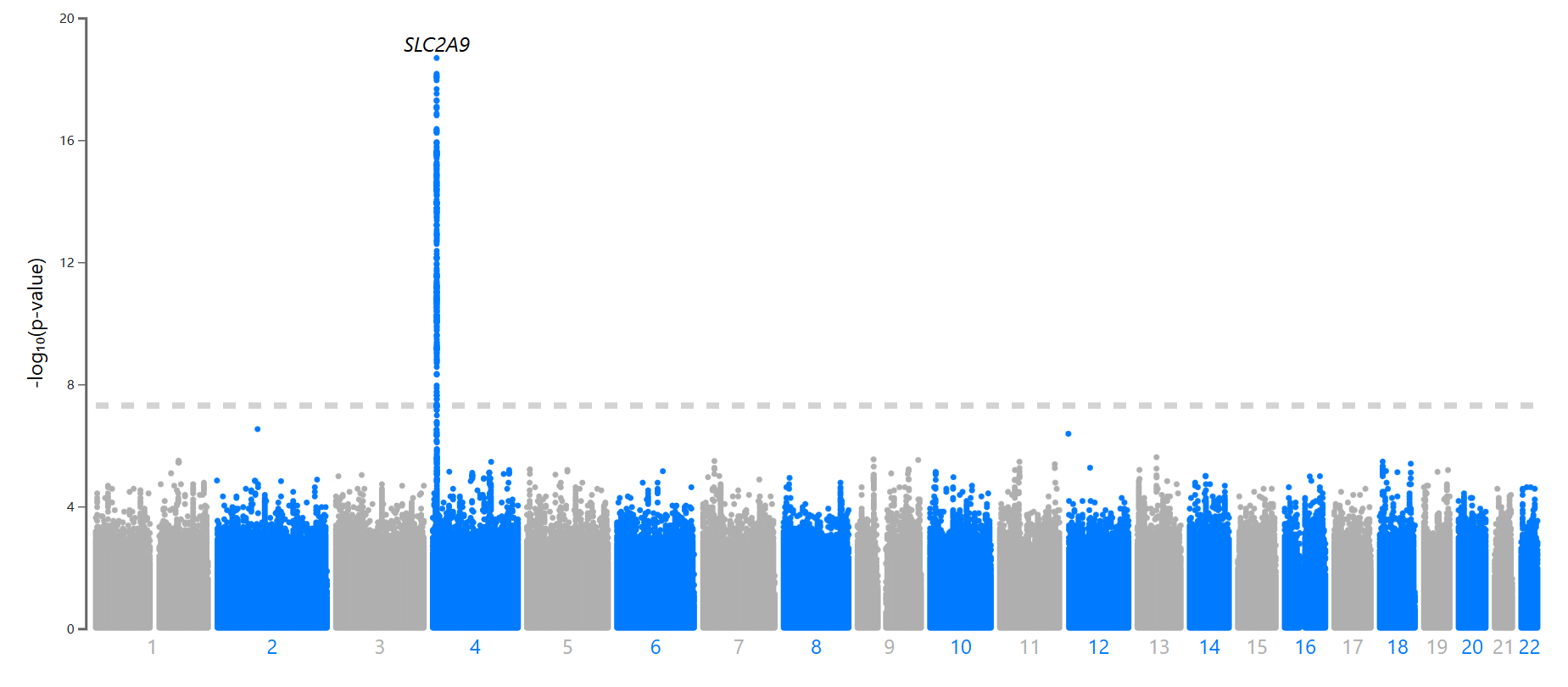

| Locus Rank | rsID | chr | pos | *p* | start | end | Nearest Gene |
| --- | --- | --- | --- | --- | --- | --- | --- |
| 1 | rs13129697 | 4 | 9926967 | 1.69 x10^-38^ | 9915325 | 10528226 | *SLC2A9* |

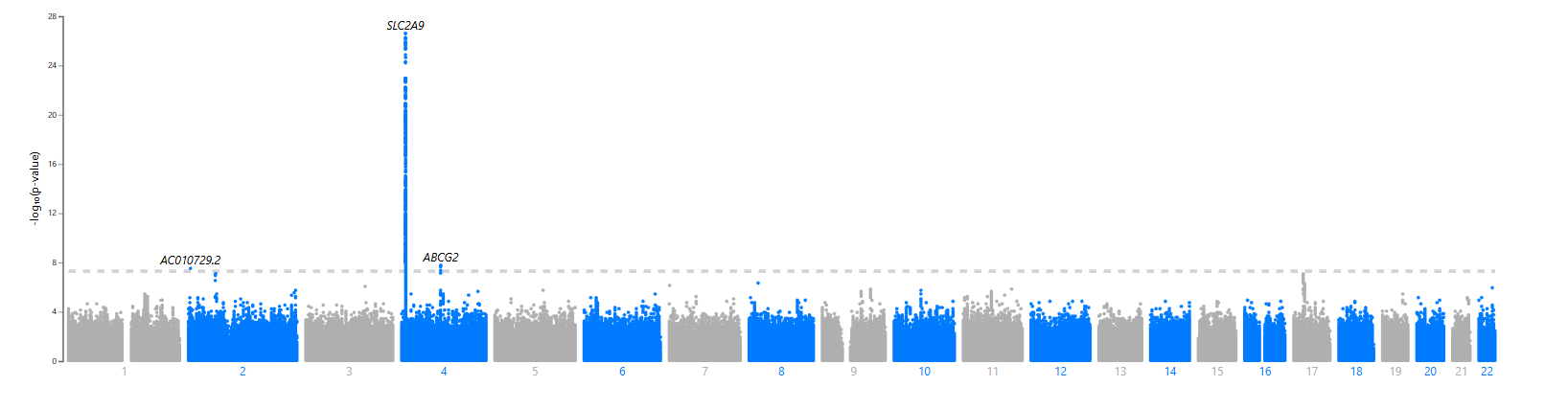
GWAS 9: Female cohorts with gout (n = 2,190)

| Locus Rank | rsID | chr | pos | *p* | start | end | Nearest Gene |
| --- | --- | --- | --- | --- | --- | --- | --- |
| 1 | rs62294282 | 4 | 9927894 | 2.49 x10^-27^ | 9915325 | 10416360 | *SLC2A9* |
| 2 | rs2231142 | 4 | 89052323 | 1.70 x10^-8^ | 89039082 | 89054667 | *ABCG2* |
| 3 | rs79261179 | 2 | 5860129 | 2.96 x10^-8^ | 5835227 | 5865222 | *AC010729.2* |
